# Measuring appropriateness of antibiotic prescribing in high-income countries: a rapid systematic review of indicators

**DOI:** 10.1101/2025.10.08.25337205

**Authors:** Rachel Berry, Lucy Catteau, Martine G. Caris, Markus G.J. de Boer, Yousra Kherabi, Eugenia Magrini, Filippo Medioli, Rita Murri, Bee Y. Ng, Nathan Peiffer-Smadja, Diane Ashiru-Oredope, ESCMID Study Group for Antimicrobial Stewardship (ESGAP)

**Author notes:** Corresponding Author; Diane Ashiru-Oredope, UK Health Security Agency, 10 South Colonnade, Canary Wharf, London, E14 4PU.

## Abstract

**Background:** Antimicrobials are essential for treating and preventing bacterial infections, yet inappropriate use drives the development of antimicrobial resistance (AMR), posing a major global health challenge. Antimicrobial stewardship (AMS) aims to optimise antibiotic use, making the definition and measurement of “appropriate” prescribing critical.

**Objectives:** The primary objective was to identify indicators used to measure appropriateness of antibiotic prescribing in high-income countries. The secondary objective was to describe levels of inappropriate prescribing in high-income countries.

**Methods:** A rapid systematic review (registration no: CRD42024628584) was conducted using Embase, Medline and Cochrane databases. Eligible studies reported indicators for measuring appropriateness of antibiotic prescribing in high-income countries. Each article was independently reviewed for inclusion, extracted, and assessed for risk of bias by one reviewer, with dual screening of 10%. Indicators were grouped as general or proxy indicators, further synthesised thematically, and described narratively. Quantitative data on inappropriateness for general indicators were standardised and summarised.

**Results:** This rapid review identified 103 unique indicators from 165 studies: 58 general indicators from 128 studies and 45 proxy indicators from 38 studies. The most frequent general indicator was compliance with guidelines (90/128, 70%). The most common proxy indicator was rate of prescribing by indication (22/38, 58%).

Indicators were applied across diverse settings, populations, and types of infection. Among studies describing general indicators, 103/128 gave quantifiable outcomes for inappropriateness of prescribing, ranging from 2% to 88%.

**Conclusions:** The review highlights the wide variety of indicators used to assess appropriateness of antibiotic use in high-income countries, reflecting the complexity of the concept. Most indicators focused on whether the correct antibiotic, dose, frequency, and duration was prescribed, in-line with clinical guidance and tailored to the clinical context - core principles of AMS. These findings provide a valuable resource for those aiming to monitor and improve antibiotic prescribing practices.

## Background

The use of antibiotics is essential for the treatment and prevention of bacterial infections and can be lifesaving. However, the growing threat of antimicrobial resistance (AMR) poses a serious global health challenge. In 2021 alone, an estimated 4.7 million deaths were associated with AMR, and projections suggest that this number could rise to 8.2 million globally by 2050 if no effective action is taken(1).

One of the key drivers of AMR is the inappropriate use of antibiotics. Across different healthcare settings, antibiotics are prescribed when not indicated, or inappropriate agents, doses, or durations are chosen(2). Antimicrobial stewardship (AMS) programs aim to address this issue by promoting the rational use of antimicrobials, thereby preserving their effectiveness and improving patient outcomes(3).

An important step in supporting AMS efforts is the ability to define and measure what constitutes “appropriate” or “inappropriate” antibiotic prescribing. This requires valid and reliable indicators that can assess the quality of antibiotic use at various levels of the healthcare system(4). Such indicators may be direct measures of appropriateness or proxy indicators; these estimate appropriateness based on prescribing patterns, adherence to guidelines, or other criteria(5).

Several initiatives and studies have attempted to develop or apply such indicators(6–9). However, no standardised set of indicators has been universally adopted across high-income countries, and practices vary according to setting, target population, and clinical indication.

This rapid systematic review aimed to identify the indicators used to estimate or measure the appropriateness of antibiotic prescribing in high-income countries (HICs) (defined as Organization for Economic Co-operation and Development (OECD) member countries(10)) and to classify these by setting, population, and indication. The secondary objective was to describe levels of inappropriate prescribing in HICs. The findings aim to inform efforts to improve antibiotic prescribing practices and strengthen AMS interventions.

## Methods

This review followed the Preferred Reporting Items for Systematic Reviews and Meta-Analyses (PRISMA) guidelines (11) with adaptations for a rapid review due to a requirement for prompt information and resource limitations as recommended by Stevens et al(12), including restriction of the publication period, limiting dual screening and extraction to 10% of included articles, and exclusion of articles not accessible through institutional subscriptions of the project group. The PRISMA checklist is provided in the web-only Supplementary Table S1. The protocol was registered with PROSPERO (CRD42024628584).

### Search strategy

A systematic literature search was conducted in Embase, MEDLINE and the Cochrane Library databases in January 2025. Policy Commons was also searched for grey literature; however, due to the rapid review design these results are not included here and will be reported separately.

The search strategy was developed by the expert review team with support with advice from UK Health Security Agency Knowledge and Evidence specialists. It targeted studies describing, developing, or evaluating indicators or metrics used to estimate the appropriateness of systemic antibiotic prescribing (treatment or prophylaxis) in any healthcare setting in HICs. Eligibility criteria are summarised in Figure 1.

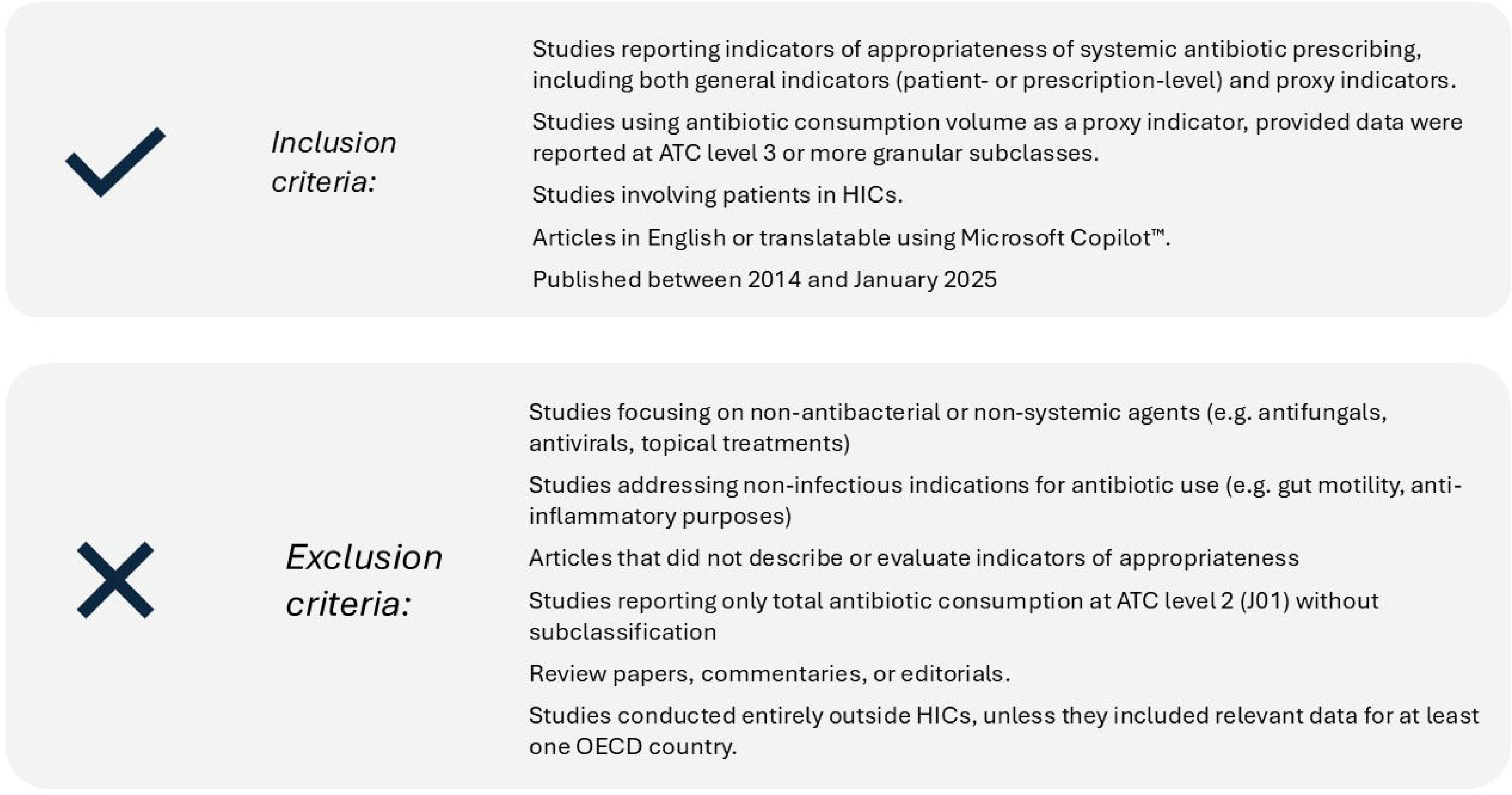

A summary of the search strategy is shown in Table 1 (for detailed search strategy see web-only Supplementary Table S2).

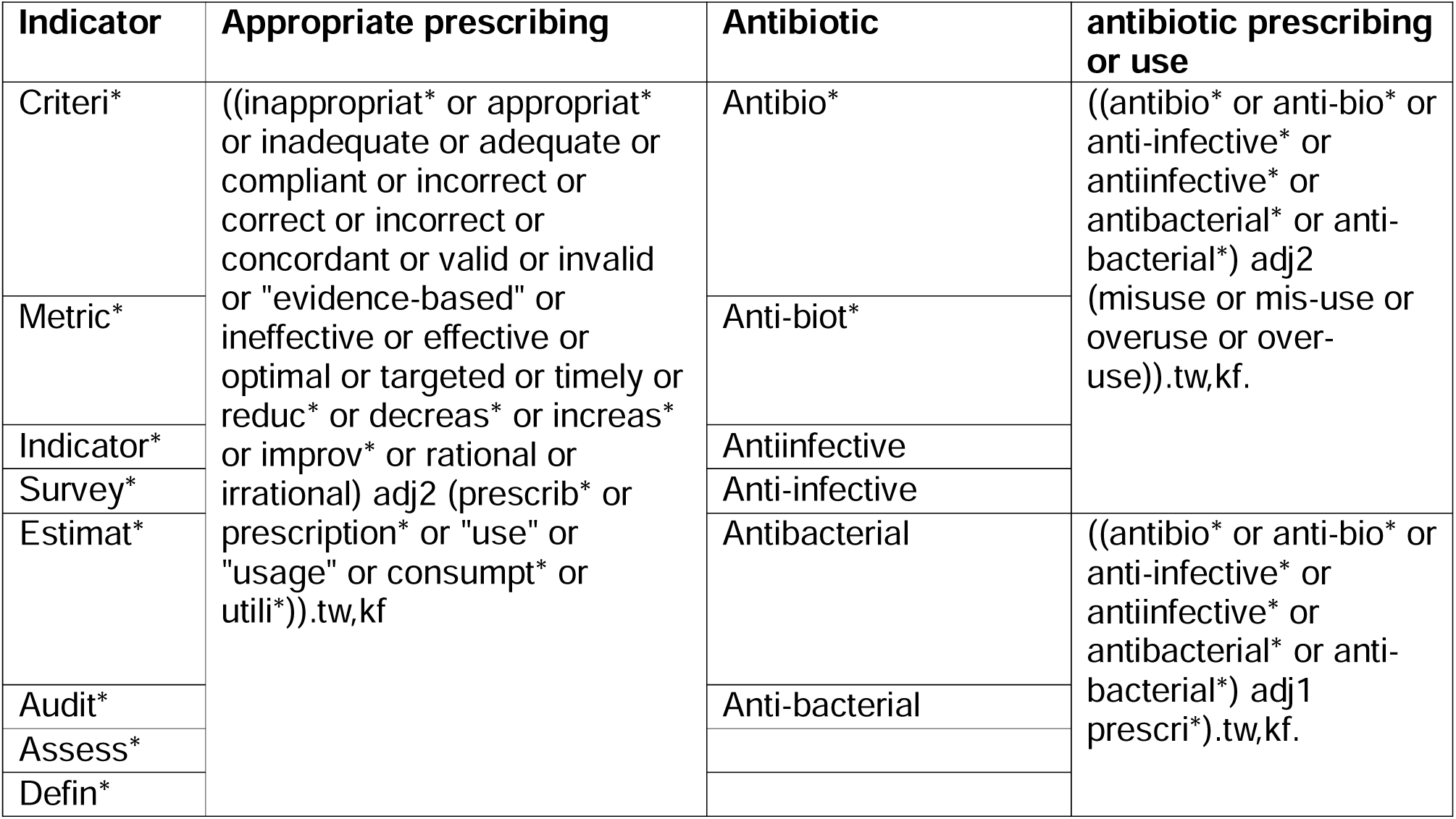

### Screening

All retrieved articles were imported into Rayyan™ and duplicates were removed. Initially, all reviewers jointly screened 10 records to refine eligibility criteria and ensure consistency. Thereafter, each article was screened by one reviewer, with 10% randomly reviewed by a second. Discrepancies were resolved through discussion or by a third reviewer, and by accessing full text if needed. Rayyan™ was used to track inclusion/exclusion decisions.

### Data extraction

A standardised data extraction tool was developed in Microsoft Excel™. Each article was extracted by one reviewer, with 10% verified in duplicate. Discrepancies were resolved by consensus or by a third reviewer.

Extracted data included:

- Study characteristics (country, setting, patient population, infection type, treatment or prophylaxis)
- Indicators used to define appropriateness of antibiotic prescribing
- Method of indicator identification and validation status
- Quantitative estimates of appropriateness (if available).

### Risk of bias assessment

Risk of bias was assessed using the Mixed Methods Appraisal Tool (MMAT)(13). Each article was assessed by one reviewer, with 10% reassessed by a second. Disagreements were resolved by a third reviewer.

### Data synthesis

Considering the heterogeneity in study designs, populations, settings and indicators, a narrative synthesis was conducted. Indicators were grouped thematically into general appropriateness indicators or proxy indicators, depending on their purpose and data source. Indicators were further categorised by setting (e.g. hospital, community), patient characteristics and infection type. For proxy indicators, required data sources were summarised into comparative tables. The thematic coding involved identifying recurring concepts or keywords from the original texts, consolidating conceptually similar indicators even if described differently. Proxy indicators were grouped by the outcome they measured (e.g., delayed prescribing, compliance). The thematic organisation, filtering, and mapping processes were conducted using Microsoft Excel™.

### Quantitative summary of inappropriateness

The secondary objective was to describe levels of inappropriate prescribing reported in HICs. Due to high variability in study design and outcome definitions, meta-analysis was not feasible. Instead, a descriptive synthesis was performed.

Studies reporting general indicators and quantitative outcomes for appropriateness were included. Definitions of appropriateness varied (e.g., “appropriate”, “inappropriate”, “unnecessary”, “unclassifiable”), so data were standardized to reflect the proportion of inappropriate prescribing. For example:

- When appropriateness was reported, all the remaining prescriptions were assumed to be inappropriate.
- For pre- and post-intervention studies, only baseline data were used.
- For studies reporting multiple infections, an average was taken. Subgroup analyses were performed by setting, as either community (including primary care, long term, and dental settings) and hospital care.

## Results

The searches returned 2,046 records. After removing duplicates, 1,495 unique papers were screened on title and abstract. Of these, 1,235 were excluded, leaving 260 for full-text review. Ten articles could not be retrieved, and a further 85 were excluded because of a wrong outcome or wrong population, leaving 165 eligible studies (see Figure 2).

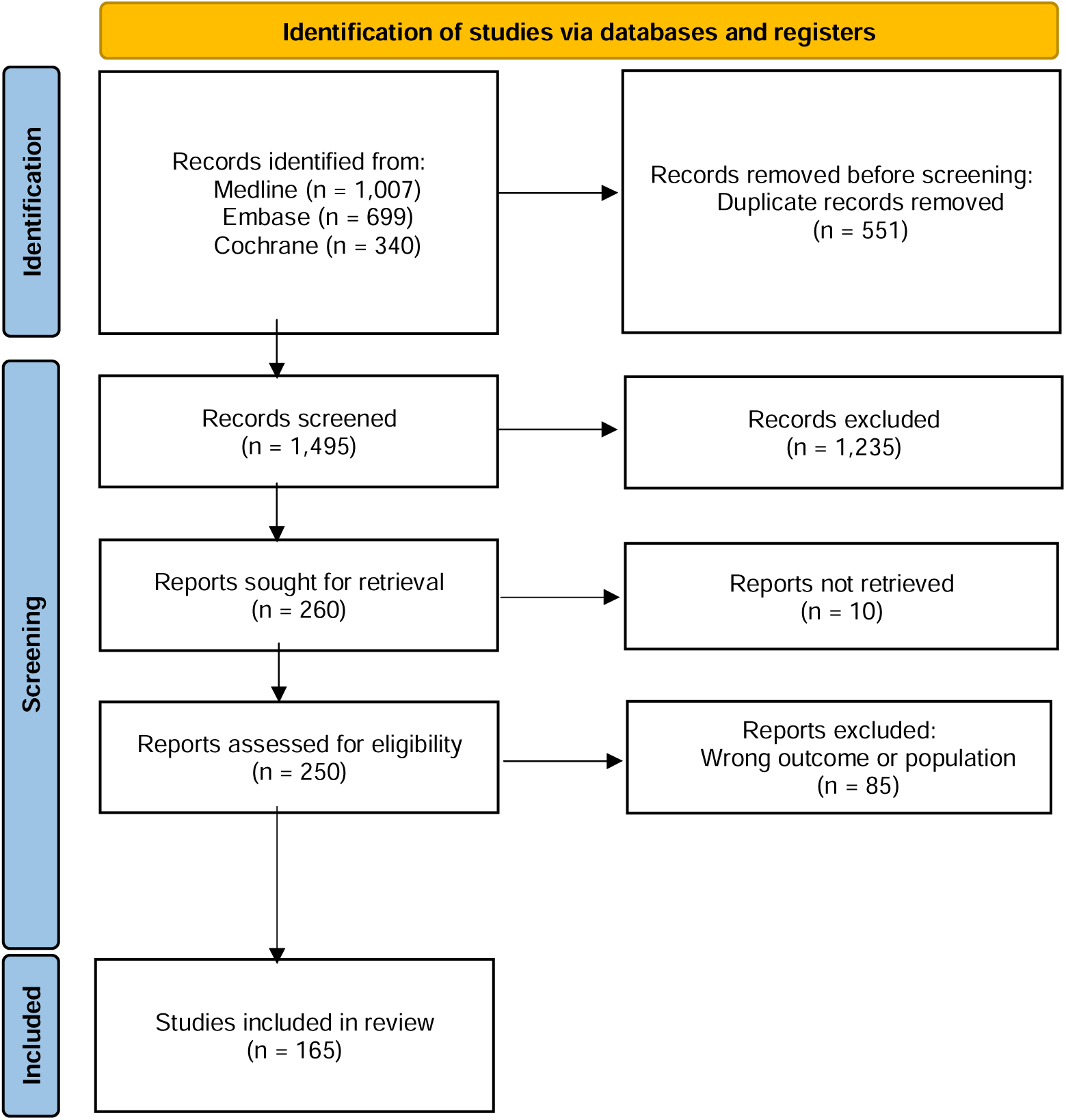

### Characteristics of the identified articles

A summary of article characteristics is provided in Table 2. For the full extraction table see web-only Supplementary Table S3.

| Characteristics | Number of articles (%)<br>Total = 165 | References |
| --- | --- | --- |
| **Country** |  |  |
| United States | 33 (20%) | (14-46) |
| United Kingdom | 17 (10%) | (9, 47-62) |
| France | 15 (9.1%) | (5, 63-76) |
| Australia | 14 (8.5%) | (77-90) |
| Netherlands | 13 (7.8%) | (91-103) |
| Spain | 11 (6.7%) | (6, 104-113) |
| Multi-country | 10 (6%) | (114-123) |
| Canada | 8 (4.8%) | (124-131) |
| Italy | 6 (3.6%) | (132-137) |
| Japan | 6 (3.6%) | (138-143) |
| Germany | 5 (3%) | (144-148) |
| Korea | 4 (2.4%) | (149-152) |
| Switzerland | 4 (2.4%) | (153-156) |
| Belgium | 3 (1.8%) | (157-159) |
| Greece | 3 (1.8%) | (160-162) |
| Ireland | 3 (1.8%) | (163-165) |
| Norway | 3 (1.8%) | (166-168) |
| Türkiye | 2 (1.2%) | (169, 170) |
| Austria | 1 (0.6%) | (171) |
| Denmark | 1 (0.6%) | (172) |
| Estonia | 1 (0.6%) | (173) |
| New Zealand | 1 (0.6%) | (174) |
| Sweden | 1 (0.6%) | (175) |
| **Healthcare Setting** |  |  |
| Hospital, including private settings | 103 (62.4%) | (6, 9, 14, 15, 18, 22, 24, 30, 31, 33-36, 40, 43, 45-59, 62, 64-68, 71, 72, 77-80, 82, 83, 85-99, 104-107, 111-113, 115, 117, 120, 122, 124-128, 130, 132-134, 136, 137, 139, 140, 142, 144, 145, 148, 153, 155, 157-162, 164, 166-169, 171, 174, 175) |
| Primary, community setting or ambulatory care | 44 (26.7%) | (5, 16, 17, 19, 20, 23, 26, 28, 29, 32, 37-39, 44, 60, 61, 69, 73, 76, 81, 84, 100-102, 108-110, 118, 119, 121, 123, 129, 131, 135, 138, 141, 147, 152, 154, 156, 165, 170, 172, 173) |
| Long term care or residential care setting | 6 (3.6%) | (21, 27, 63, 70, 75, 163) |
| Multiple settings (see detail) | 6 (3.6%) | (103, 114, 116, 149-151) |
| Dental | 5 (3%) | (25, 41, 42, 74, 146) |
| Community Pharmacy | 1 (0.6%) | (143) |
| **Population** |  |  |
| Not specified | 94 (56.9%) | (5, 6, 9, 14, 17-19, 22-24, 29, 31-33, 35-37, 39, 40, 45, 48-50, 52, 53, 55, 56, 58, 66-68, 71-73, 77-84, 88-93, 95, 96, 100-103, 106, 108, 110, 112, 114-116, 118, 121, 124-127, 134, 136, 139-149, 151, 153-155, 157, 158, 160, 161, 164, 166, 169, 171, 173, 175) |
| Adults | 41 (24.8%) | (16, 21, 25-27, 30, 41-44, 47, 54, 57, 59, 63, 70, 74, 75, 86, 94, 97-99, 104, 105, 107, 111, 117, 120, 122, 128-130, 133, 135, 152, 156, 159, 163, 170, 174) |
| Children | 16 (9.7%) | (15, 20, 34, 46, 51, 61, 62, 76, 87, 113, 119, 137, 138, 162, 167, 168) |
| Adults and children | 11 (6.7%) | (28, 38, 60, 69, 109, 123, 131, 132, 150, 165, 172) |
| Older adults >75 | 2 (1.2%) | (64, 65) |
| Neonates <28 days | 1 (0.6%) | (85) |
| **Treatment or prophylaxis** |  |  |
| Treatment | 87 (52.7%) | (5, 6, 9, 15-17, 19-24, 26, 28-30, 32, 34, 37-39, 43, 44, 49, 50, 52, 56, 57, 59-61, 63-66, 70-76, 81, 84, 85, 88, 93, 94, 96, 98-105, 107-112, 117, 121, 122, 125-131, 133, 137, 142, 143, 147, 149, 154, 156, 158, 159, 165, 166, 170, 172) |
| Both | 59 (35.8%) | (14, 25, 27, 33, 36, 40, 45-47, 51, 53, 58, 62, 67, 69, 77-80, 83, 86, 87, 90, 91, 95, 97, 106, 113-116, 120, 123, 124, 135, 136, 138-140, 144-146, 148, 150-152, 155, 157, 161-164, 167-169, 171, 173-175) |
| Prophylaxis | 10 (6%) | (31, 35, 41, 42, 48, 54, 55, 82, 132, 160) |
| Not specified | 9 (5.5%) | (18, 68, 89, 92, 118, 119, 134, 141, 153) |
| **Clinical Indication** |  |  |
| Not specified | 97 (58.8%) | (5, 6, 9, 14, 15, 18, 24, 30, 33, 34, 43, 45, 46, 49, 51, 52, 56-58, 63-69, 71-73, 75-81, 83, 86, 87, 91-95, 97, 99, 101, 103, 104, 106-108, 111-113, 115, 116, 118-120, 122, 124-128, 130, 134-141, 143-145, 148, 150-153, 155, 157-159, 161, 163, 164, 167-169, 171, 173-175) |
| Respiratory Tract Infections | 18 (10.9%) | (16, 20, 23, 29, 38, 39, 44, 61, 84, 110, 117, 121, 123, 133, 165, 166, 170, 172) |
| Various | 11 (6.7%) | (28, 32, 40, 53, 60, 100, 102, 109, 129, 131, 147) |
| Dental or oral infections or prophylaxis | 9 (5.5%) | (25, 36, 41, 42, 50, 62, 74, 90, 146) |
| Urinary tract infection | 8 (5%) | (22, 26, 27, 70, 96, 149, 156, 162) |
| Prophylaxis of surgical or procedural infection | 7 (4.2%) | (31, 35, 48, 54, 55, 82, 160) |
| Sepsis or bacteraemia | 5 (3%) | (85, 89, 98, 105, 142) |
| Urinary tract infections and Respiratory tract infections | 4 (2.4%) | (17, 21, 114, 154) |
| Urinary tract Infection and Skin and soft tissue infections | 1 (0.6%) | (19) |
| Healthcare Associated Infections | 1 (0.6%) | (132) |
| Acute Pancreatitis | 1 (0.6%) | (47) |
| Skin and soft tissue infections or post-surgery infections | 1 (0.6%) | (88) |
| Suspected or confirmed intra-abdominal infection | 1 (0.6%) | (59) |
| Chlamydia trachomatis infection | 1 (0.6%) | (37) |
| **Specific criteria** |  |  |
| Patients prescribed specific antibiotics | 18 (10.9%) | (24, 43, 49, 58, 59, 66-69, 86, 104, 107, 112, 125, 128, 130, 143, 159) |
| Patients in the emergency department | 5 (3%) | (45, 79, 93, 94, 111) |
| Patients on critical or intensive care | 4 (2.4%) | (80, 92, 120, 127) |
| Veterans | 4 (2.4%) | (16, 41-43) |
| Patients in the dental department | 3 (1.8%) | (36, 50, 62) |
| Medical Patients | 3 (1.8%) | (56, 57, 134) |
| Oncology or cancer patients | 2 (1.2%) | (22, 54) |
| Patients on Out-patient Parenteral Antibiotic Therapy (OPAT) | 2 (1.2%) | (81, 109) |
| Hospitalised older adults | 2 (1.2%) | (64, 65) |
| Patients prescribed intravenous (IV) antibiotics | 2 (1.2%) | (97, 169) |
| Patients prescribed oral antibiotics | 2 (1.2%) | (14, 15) |
| Haemodialysis patients | 1 (0.6%) | (30) |
| **Method used for indicator identification** |  |  |
| Literature review | 46 (27.9%) | (5, 6, 15, 22, 24, 27, 29, 32, 34, 35, 38-40, 45, 54, 58, 67-69, 71, 72, 75, 76, 78, 93, 95, 97, 100, 106, 108, 109, 113, 117, 120, 129, 139, 140, 142, 144, 149, 152, 153, 159, 162, 169, 172) |
| Using treatment or diagnostic guidelines | 35 (21.2%) | (16, 19-21, 23, 25, 26, 30, 31, 36, 37, 41, 42, 44, 48, 50, 55, 59, 62, 66, 74, 84, 99, 105, 107, 110, 121, 125, 128, |
|  |  | 133, 137, 155, 160, 166, 170) |
| Not specified | 30 (18.2%) | (14, 17, 18, 33, 47, 57, 87, 91, 103, 104, 111, 112, 114, 115, 119, 124, 130, 132, 134, 136, 138, 143, 145, 148, 161, 164, 167, 168, 171, 175) |
| Pre-existing Audit tool | 16 (9.7%) | (46, 52, 56, 77, 79-82, 85, 86, 88-90, 126, 158, 174) |
| Delphi consensus | 15 (9.1%) | (9, 63, 65, 73, 92, 94, 96, 98, 122, 127, 131, 146, 150, 157, 163) |
| Pre-existing indicator | 12 (7.3%) | (51, 61, 101, 118, 123, 135, 141, 147, 154, 156, 165, 173) |
| Expert group | 11 (6.7%) | (28, 43, 49, 53, 60, 64, 70, 83, 102, 116, 151) |
| **Validation status of the indicators** |  |  |
| Yes – in this study | 62 (37.6%) | (5, 9, 17, 20-24, 26, 29, 31, 32, 34, 35, 37, 39, 42-44, 47-50, 53, 56, 57, 63, 65-67, 70, 71, 73, 76, 78, 83, 93, 96, 98, 99, 101-104, 106, 108, 109, 116, 117, 122, 125, 128, 140, 144-146, 151, 152, 154, 160, 166, 171) |
| Yes – in previous study | 47 (28.5%) | (6, 27, 38, 40, 45, 46, 58, 61, 68, 69, 72-74, 77, 79, 80, 82, 85, 86, 88, 89, 95, 97, 110, 118, 120, 121, 123, 126, 129, 132, 133, 135, 136, 139, 147, 153, 155, 156, 158, 159, 161, 165, 169, 173-175) |
| No | 32 (19.4%) | (15, 18, 30, 33, 41, 51, 52, 54, 59, 60, 62, 90, 92, 94, 100, 105, 113, 114, 124, 127, 130, 131, 134, 137, 138, 143, 148, 150, 157, 162-164, 167) |
| Not specified | 24 (14.5%) | (14, 16, 19, 25, 28, 36, 55, 60, 64, 81, 84, 87, 91, 107, 111, 112, 115, 119, 141, 142, 149, 168, 170, 172) |
| **Indicator type described** |  |  |
| General indicators | 127 (77%) | (6, 9, 14-19, 21-27, 30, 31, 33, 35-37, 41-50, 52-59, 62, 64-72, 77-83, 85-99, 103-108, 110-117, 120-122, 124-128, 130, 132-134, 136-140, 142, 144, 145, 149-153, 155, 157-164, 166-171, 174, 175) |
| Proxy indicators | 37 (22.3%) | (5, 20, 28, 29, 32, 34, 38-40, 51, 60, 61, 63, 73-76, 84, 100-102, 118, 119, 123, 129, 135, 141, 143, 146-148, 154, 156, 165, 172, 173) |
| Both | 1 (0.7%) | (109) |

Among the 165 included articles, the most frequently represented countries were: the United States (33/165, 20%), the United Kingdom (17/165,10%), France (15/165, 9.1%), Australia (14/165, 8.5%), the Netherlands (13/165, 7.8%) and Spain (11/165, 6.7%). Ten articles were conducted across multiple countries (6%).

Most articles focused on the hospital or inpatient settings (103/165, 62.4%) and did not specify the age group of the patient population (94/165, 56.9%). The majority addressed treatment (87/165, 52.7%) rather than prophylaxis and 58.8% (97/165) did not specify a particular clinical indication.

Most articles described indicators identified utilising a literature review (46/165, 27.9%) or based on diagnostic or treatment guidelines (35/165, 21.2%). 62 articles (37.6%) validated indicators within the study itself, 47 articles (28.5%) referred to validation in a previous study. Around a third used indicators that were either not validated (32/165, 19.4%) or did not specify validation status (24/165, 14.5%).

Regarding indicator type, 127 articles (77%) described general indicators, 37 articles (22.3%) described proxy-indicators, and one article described both general and proxy indicators.

### Risk of bias assessment

The full risk of bias assessment is provided in the web-only Supplementary Table S4. A summary of the Risk of Bias assessment is shown in Figure 3.

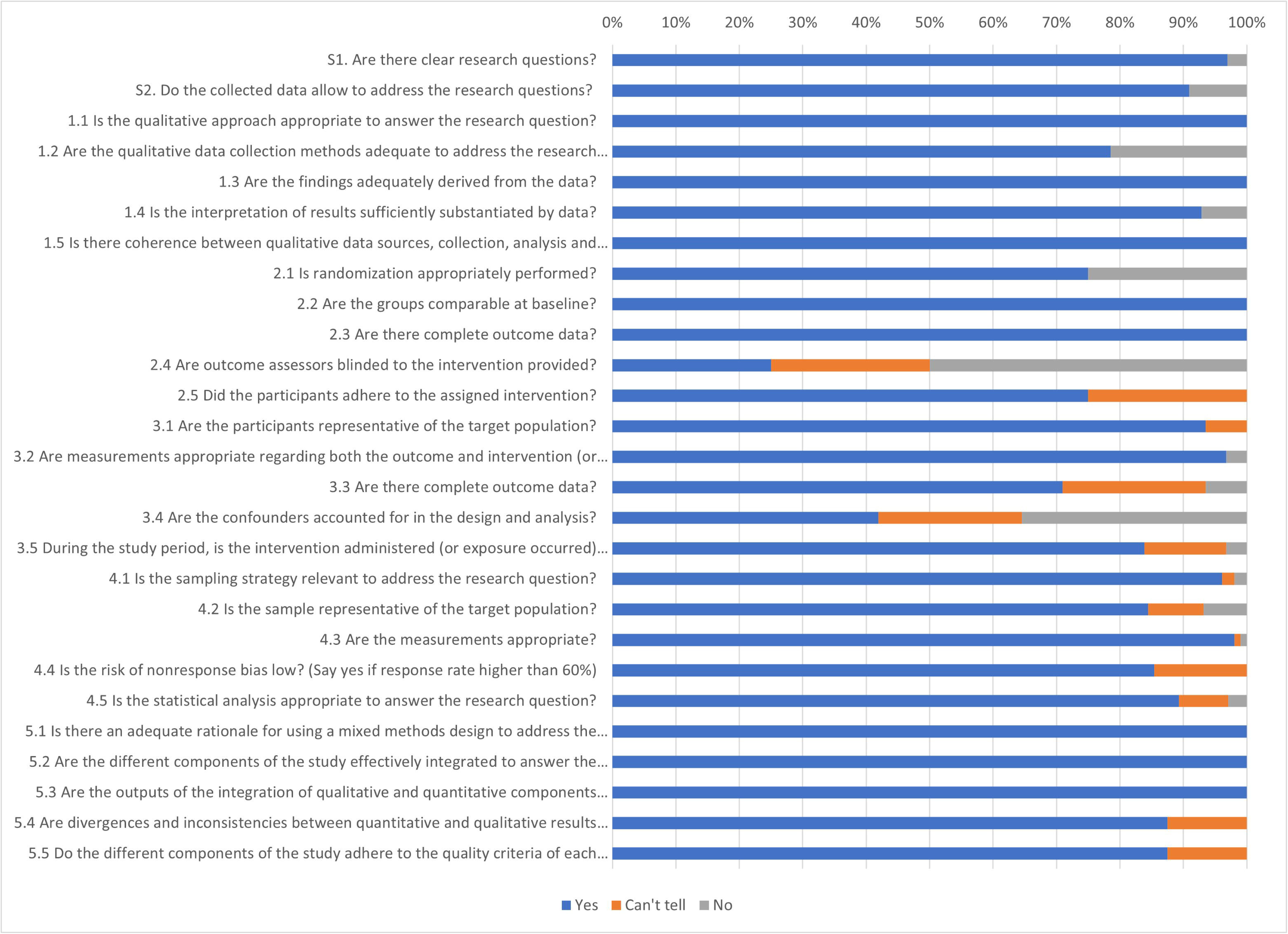

Overall, studies were considered to be of moderate to good quality as 103 of the included studies (62.4%) fulfilled 100% of the relevant quality criteria, 31 studies (18.8%) fulfilled 80%, and 13 fulfilled 60% (10.9%). Five studies were excluded from assessment as they did not report methods or results. Although 8 studies fulfilled less than 50%, this represented a small proportion of the total number of included studies (4.8%).

### Indicators

#### General Indicators

A total of 127 studies described general indicators of appropriateness (127/165, 77%), and one described both general and proxy indicators (1/165, 0.7%) (Table 2).

Among the 128 studies describing general indicators, the majority were from the United States (25/128, 19.5%), followed by the United Kingdom (14/128, 10.9%), and Australia (13/128, 10.1%). Most focused on the hospital or in-patient settings (99/128, 77.3%), and more than half did not specify patient age (76/128, 59.3%) or the infection type (82/128, 64%). General indicators were most often identified through literature review (34/128, 26.6%) or treatment/diagnostic guidelines (32/128, 25%). In terms of validation, 49/128 (38.2%) validated indicators within the study.

In total, 58 distinct general indicators were identified as measures of appropriate antibiotic prescribing in HICs. These indicators are presented in Table 3 with references provided in the web-only Supplementary Table S5.

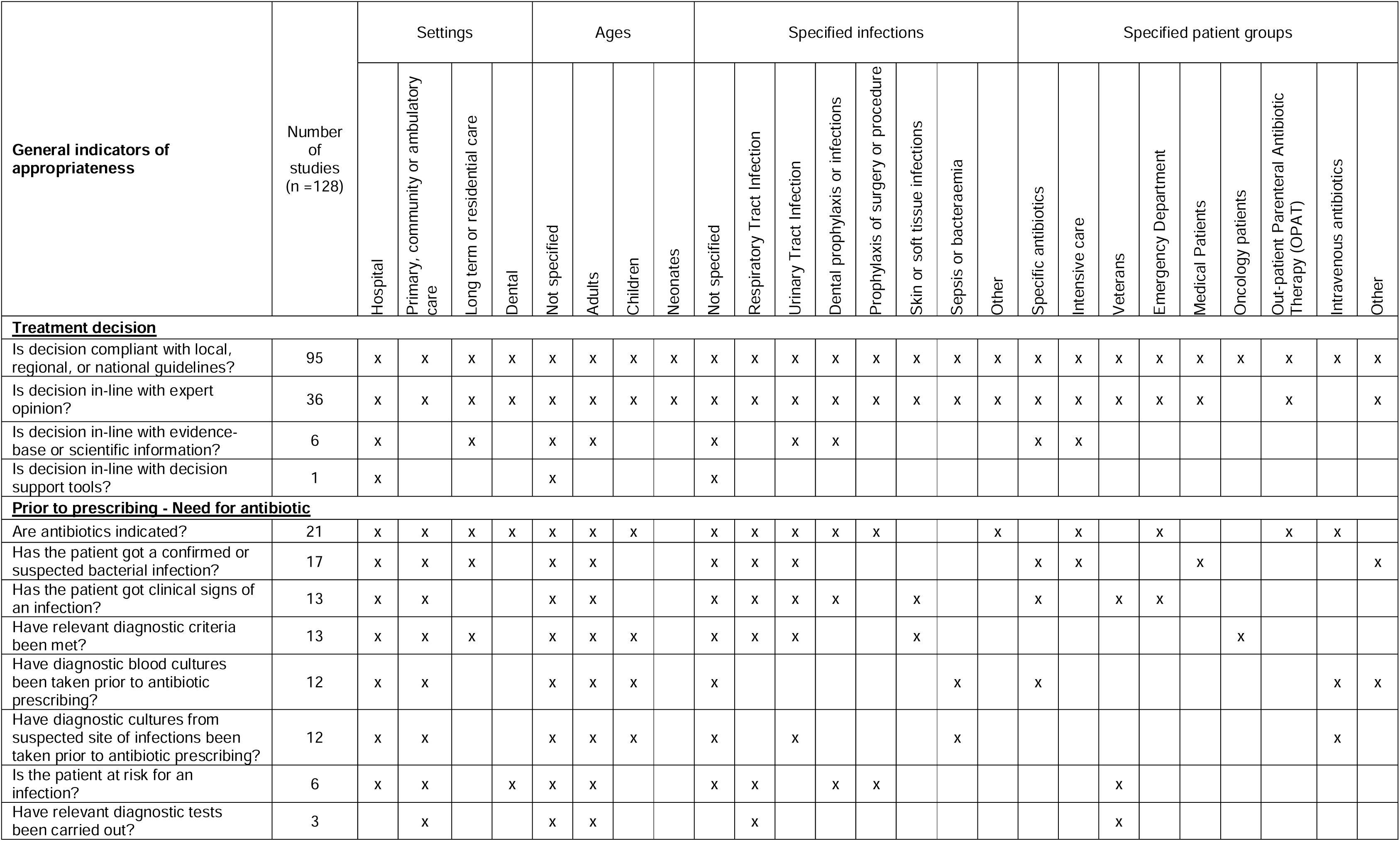
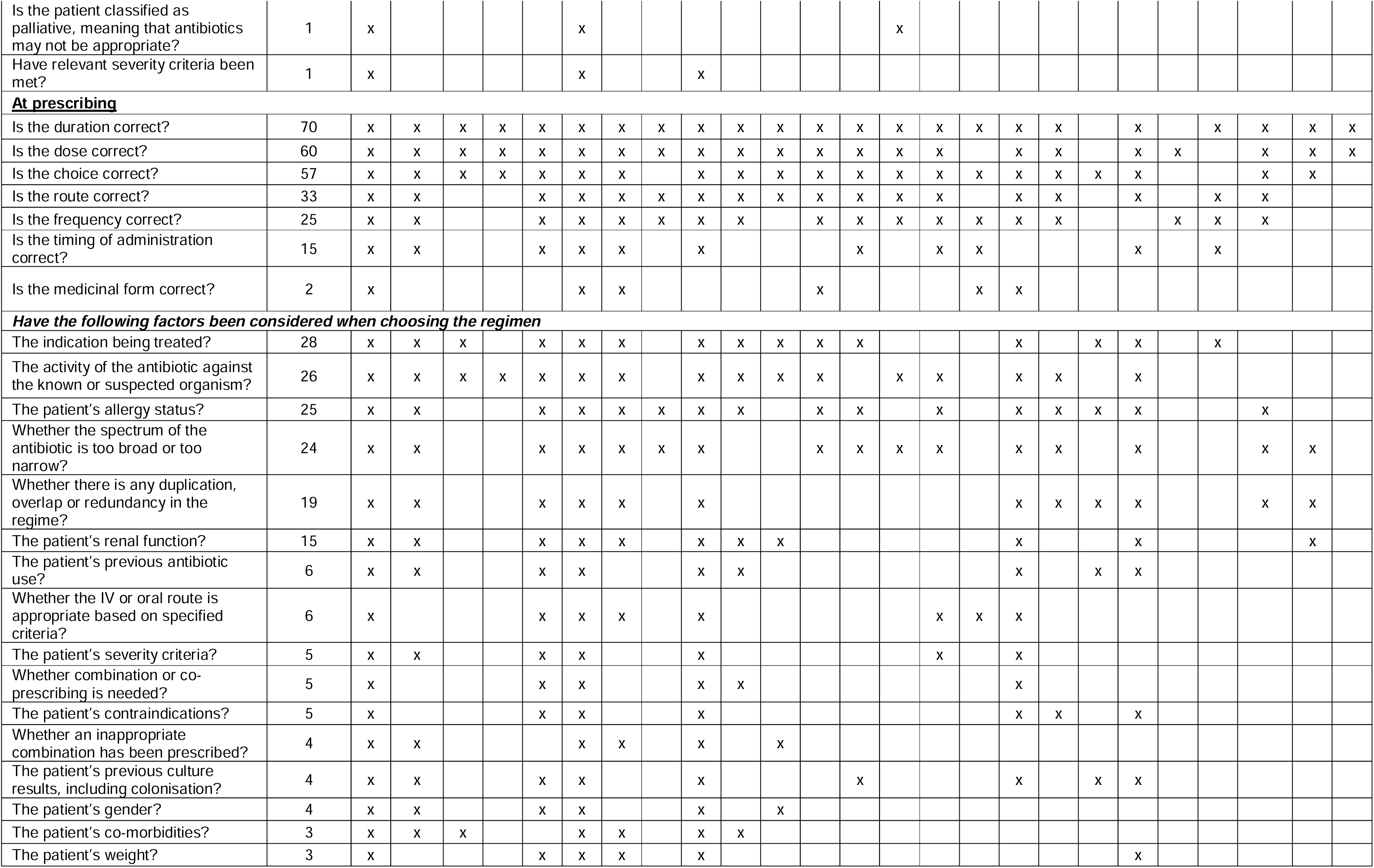
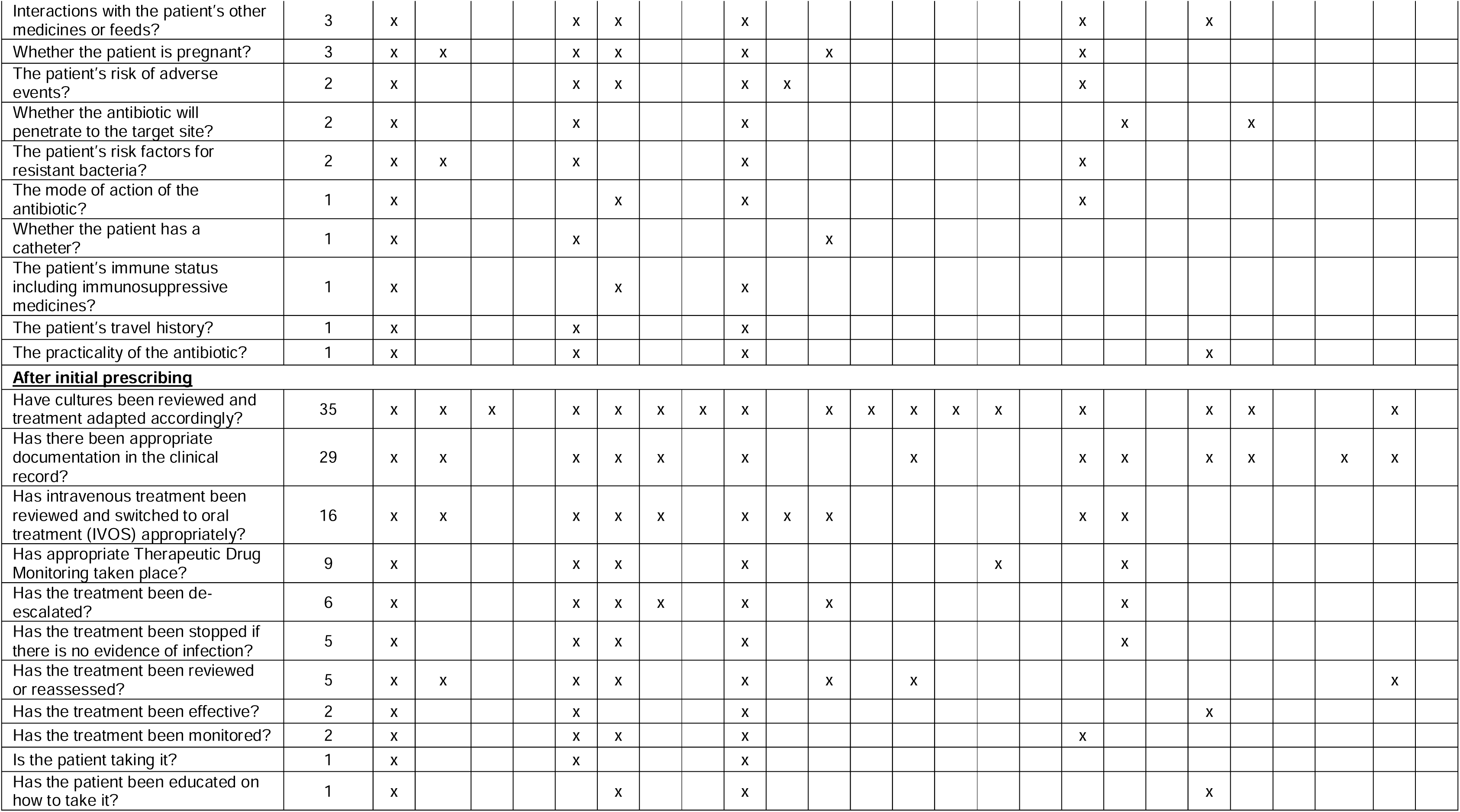

Most articles (92/128, 71.8%) employed composite definitions of appropriate prescribing, where multiple indicators were required to be met for a prescription to be classified as appropriate(6, 9, 14, 16–19, 24, 25, 30, 31, 36, 37, 41–47, 49, 50, 52–54, 56, 59, 62, 66–72, 77–83, 85–90, 93, 95, 103–107, 110–117, 120, 124–128, 130, 133, 134, 137, 139, 140, 142, 144, 149, 151–153, 155, 157, 159, 160, 163, 164, 166, 168, 169, 171, 174). These definitions incorporated between 2 and 16 individual indicators.

The most frequently reported general indicator of appropriate prescribing was compliance with local, national, or regional guidelines, reported in 90 of 128 articles (70.3%). This was followed by indicators related to appropriate duration of treatment (70/128, 54.7%) and appropriate dose (60/128, 46.9%). These indicators were applied consistently across all clinical settings, age groups, and infection types.

When analysed by setting, hospitals demonstrated the broadest application of general indicators, utilising 57 out of the 58 identified. In contrast, primary care, outpatient, and ambulatory care settings employed a more limited set, (34/58 general indicators), while in long-term or residential care settings 13 indicators were reported, and only 8 in dental settings.

By patient age group, the widest range of general indicators was reported in studies involving adults (50/58 indicators). The remaining eight indicators appeared only in studies that did not specify any age group, suggesting potential applicability to adult populations. In children, 27 indicators were described, and a narrower range was used in neonates, with only 9 indicators reported.

By infection type, fewer general indicators were reported . Urinary tract infections were associated with 23/58 indicators, respiratory tract infections with 22/58 indicators. Dental prophylaxis and treatment, prophylaxis of surgery or procedures, and sepsis or bacteraemia were each associated with 17 indicators; a narrower range was described for other identified infections.

#### Proxy-indicators

Thirty-seven articles described proxy indicators for assessing or estimating appropriateness (37/165, 22.3%) and one described both general and proxy indicators (1/165, 0.7%).

Of these 38 articles, most were from the United States (8/38, 21%) and France (6/38, 15.7%). Most studies focused on primary care, outpatient, or ambulatory settings (29/38, 76.3%) and did not specify patients age (15/38, 39.5%) or infection type (18/38, 47.3%), although some indicators themselves were infection-specific. Most indicators were identified through a literature review (13/38, 34.2%) or based on pre-existing proxy indicators (12/38, 31.6%), with 14/38 (36.8%) validated in the current study.

A total of 45 proxy-indicators were identified to assess appropriateness of antibiotic prescribing in HICs. These are listed in Table 4, with references included in web-only Supplementary Table S6.

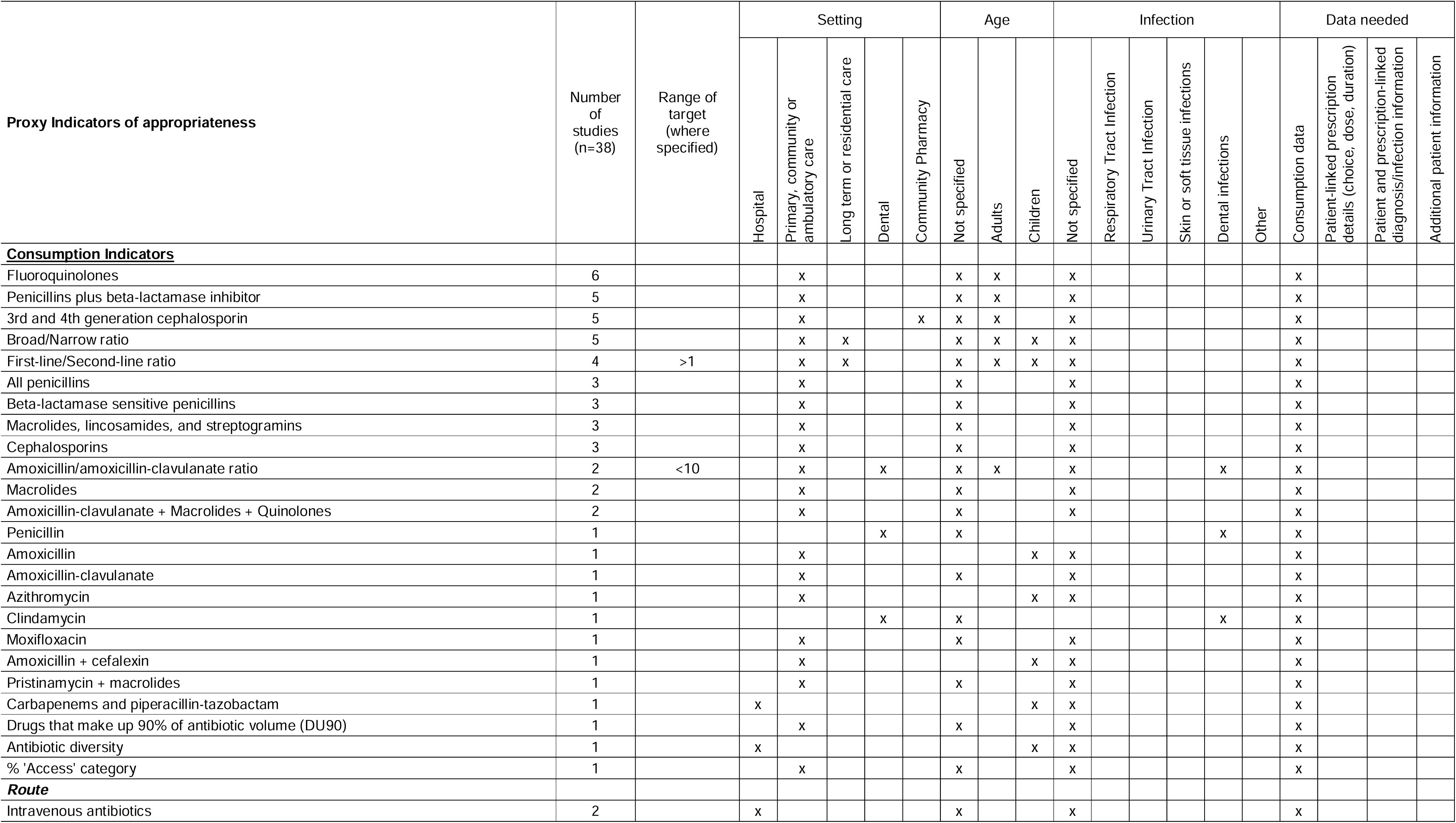
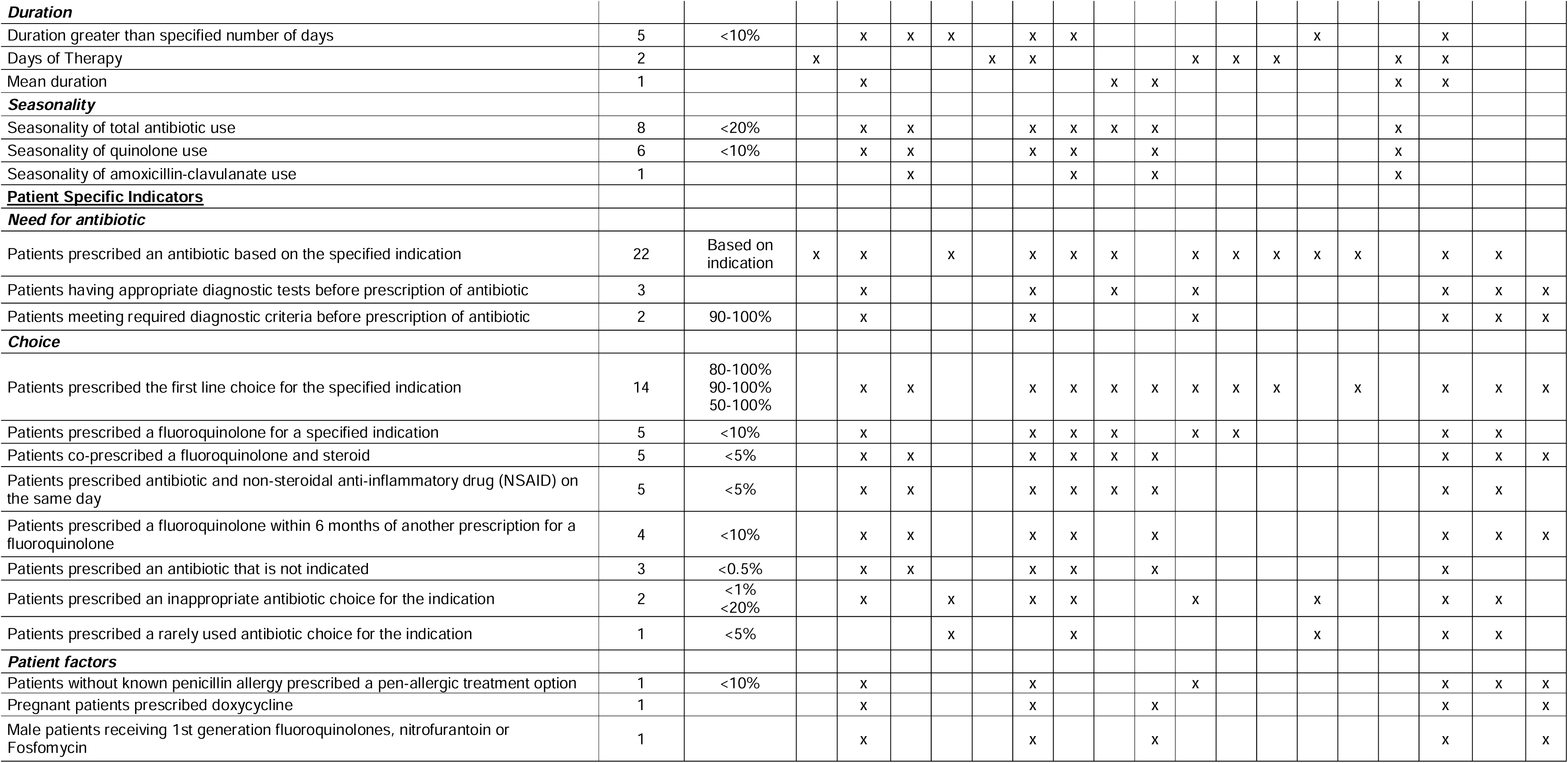

Overall, the most frequently reported proxy indicator of appropriate prescribing was the rate of prescribing by indication, described in 22 out of 38 articles (57.9%). The second most common was the proportion of patients receiving first-line treatment according to indication (14/38, 36.8%). Both were applied across most settings, age groups, and infection types.

When analysed by setting, primary care demonstrated the broadest application of proxy indicators, with 37 of the 45 identified. Long-term care settings employed a narrower subset of 11 proxy indicators. In contrast, hospital, dental and community pharmacy settings were associated with only a limited number of proxy indicators (5, 7 and 2 respectively).

By age group, the widest range of proxy indicators was reported in studies involving adults, with 19 of the 45 identified proxy indicators. However, 40 of the 45 were either reported for adults or not age-specified, suggesting broad applicability to adult populations. In children, 15 proxy indicators were described, six of which were unique to this group.

When classified by infection type, a narrower scope was observed. Respiratory tract infections were associated with eight proxy-indicators, dental infections with seven, urinary tract infections with four, skin and soft tissue infections with three and other infections with two.

### Levels of inappropriateness of antibiotic prescribing

A total of 103 articles describing general indicators provided quantifiable information on inappropriateness of prescribing (103/165, 62.4%)(6, 14–17, 19, 21–26, 30, 31, 35–37, 41–50, 52–59, 62, 66–72, 77–83, 85–90, 93, 95, 104–108, 110, 111, 113, 116, 117, 120, 121, 124–126, 128, 130, 132–134, 136–140, 144, 149, 151, 152, 155, 158–162, 164, 166–171, 174, 175).

Of these, most were conducted in hospital settings (79/103, 77.5%)(6, 14, 15, 22, 24, 30, 31, 35, 36, 43, 45–50, 52–59, 62, 66–68, 71, 72, 77–80, 82, 83, 85–90, 93, 95, 104–107, 111, 113, 117, 120, 124–126, 128, 130, 132–134, 136, 137, 139, 140, 144, 155, 158–162, 164, 166–169, 171, 174, 175), with a small proportion addressing multiple setting (3/103, 2.9%)(116, 149, 151), and the remainder focusing on community settings, including primary care, dental and long-term care (20/103, 19.6%)(16, 17, 19, 21, 23, 25, 26, 37, 41, 42, 44, 68, 70, 81, 108, 110, 121, 138, 152, 170).

Reported levels of inappropriate prescribing ranged from 2.2% to 88%. Hospital and multi-setting studies spanned this entire range, whereas community-based studies (primary care, dental and long-term care facilities) reported inappropriateness between 11% and 85%.

In hospitals or multiple settings, most studies reported inappropriate prescribing rates below 40% (53/82, 64.6%), although there were still articles that reported higher levels (Figure 4). In community settings a wider range can be seen, with the majority reporting between 10 and 70% inappropriateness (16/20 studies, 80%).

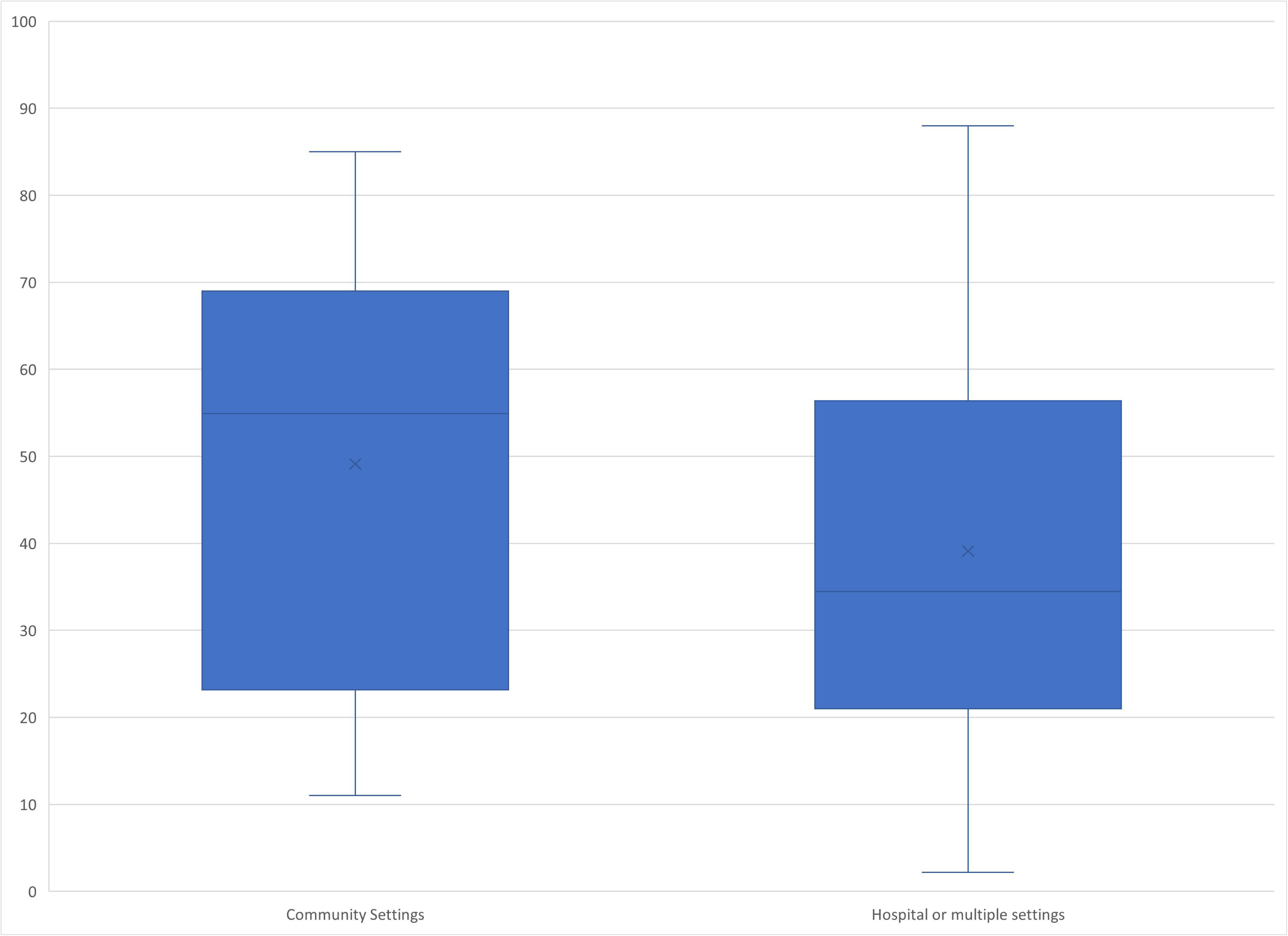

## Discussion

### Indicators

This review identified 103 unique indicators - 58 general and 45 proxy indicators - used to assess the appropriateness of systemic antibiotic prescribing across 22 HICs. These indicators reflect diverse approaches, data sources, and clinical contexts, with a predominant focus on hospital settings and treatment-related decisions.

The breadth of indicators underscores the complexity of defining appropriateness of antibiotic prescribing. Many were composite, combining agent choice, dose, route, duration, and clinical justification, highlighting that appropriateness is multidimensional and rarely captured by a single metric. This mirrors the complexity of stewardship practice, which must balance patient, pathogen, and clinical context- (176). However, the lack of standardisation in how indicators are defined and applied across studies limits comparability and benchmarking. National initiatives such as Australia’s national antimicrobial prescribing surveys (NAPS) demonstrate the value of defined, harmonised indicators for repeated measurement and feedback (8). Proxy indicators, though less granular, offer scalable metrics where patient-level data are not available.

#### Variation by setting, population and infection

General indicators were predominantly applied in hospitals where detailed clinical data and stewardship infrastructure are available(177). In contrast, proxy-indicators such as prescribing volume or adherence to guidelines were more common in primary care. This reflects the challenges in accessing patient-level clinical justification in these settings, where audits are less frequent and electronic prescribing systems less integrated with clinical records (73). The smaller number of published studies covering primary care might also explain this trend. These settings are often not included in national stewardship programmes such as the Point Prevalence Survey (PPS), may lack a dedicated antimicrobial stewardship team, and audits are typically conducted less frequently or at smaller scale. Additionally, data collection in primary care often relies on coded clinical notes and electronic searches, rather than patient-level audits more common in hospitals. With the expansion of digital health tools and electronic records in hospitals, there is an opportunity to improve access to patient-level prescribing data in all settings in high-income countries. This could enable more nuanced assessments of appropriateness, but further research is needed to explore feasibility and implementation.

Surprisingly, few studies specified patient age, restricting applicability to paediatrics or older adults, where prescribing recommendations differ. Similarly, infection-specific indicators were underused, despite their potential to provide more targeted feedback for high-impact conditions such as respiratory or urinary tract infections.

Despite heterogeneity, recurring themes emerged: indicators frequently assessed whether antibiotics were prescribed when indicated, whether the agent, dose, frequency, and duration were appropriate, and whether prescribing was consistent with clinical guidance. These recurring themes reflect core principles of antimicrobial stewardship and provide a foundation for harmonisation.

### Levels of inappropriateness of antibiotic prescribing

Reported rates of inappropriate prescribing varied widely, in some cases even exceeding 80%, but estimates were highly sensitive to how inappropriateness was defined and measured. Proxy indicators often inferred inappropriateness from broad-spectrum use or non-adherence to guidelines, while general indicators relied on detailed audits. This variability highlights the urgent need for transparent, standardised definitions and validated indicators.

### Limitations

This rapid review prioritised timeliness over comprehensiveness. Searches were limited to selected databases, terms in titles/abstracts, publication since 2014 and peer-reviewed literature; grey literature was excluded. While some indicators may have been missed, the review likely achieved broad coverage of those currently applied. Included studies were heterogeneous in design, populations, and outcome definitions, limiting comparability. Also, while we attempted to be comprehensive, the review excluded studies that reported only total antibiotic consumption at the ATC level 2 category (J01-Antimicrobials for systemic use) without further subclassification, as well as studies that did not specify indicators explicitly. As a result, some relevant examples of practical indicators may have been missed.

Moreover, we focused only on high-income countries and prescribing indicators, not diagnosis. This means that important pre-prescription determinants of appropriateness—such as diagnostic accuracy or indication validity—are not fully addressed. Also, due to variation in reporting, assumptions were occasionally made to classify indicators, which may have introduced subjectivity. Nonetheless, because the aim was to catalogue indicators rather than synthesise outcomes, these limitations are unlikely to undermine the main findings.

### Implications and future work

This review provides a structured resource for health authorities, AMS teams, and researchers aiming to monitor and improve antibiotic prescribing. It highlights the need for a core set of validated, standardised indicators, adaptable across settings and infection types. Proxy indicators can facilitate large-scale surveillance where detailed data are lacking, while general indicators remain essential for in-depth audits and quality improvement. Future research should explore how indicators perform across diverse populations, how they link to patient outcomes, and how they can be integrated into electronic health record systems to support real-time feedback and benchmarking.

## Conclusion

This review is the first to systematically catalogue and categorise both general and proxy indicators of antibiotic prescribing appropriateness across high income countries. Despite heterogeneity, recurring themes reflect shared stewardship principles: prescribing the right antibiotic, at the right dose and duration, in line with clinical guidance. Indicators varied in granularity and application, but together they provide a foundation for harmonisation.

Developing a core set of validated, standardised indicators, integrated within digital health systems, will be critical to enable benchmarking, enhance stewardship and ultimately improve antibiotic use across healthcare settings.

## Data Availability

All data produced in the present work are contained in the manuscript

## Declaration of Generative AI and AI-assisted technologies in the writing process

During the preparation of this work the authors used Microsoft Copilot™ in order to translate articles that were not in English, and to improve readability and language of the text. After using this tool/service, the authors reviewed and edited the content as needed and take full responsibility for the content of the publication

## Conflict of interest

None.

## Funding

The article processing fee was supported by the ESCMID Study Group for Antimicrobial Stewardship (ESGAP).

## Contribution

R.B. and D.A.O conceptualised the study. R.B., L.C., M.G.C., M.G.J.dB., E.M., F.M., R.M., N.P.F., and D.A.O. designed the study protocol including search terms and inclusion/exclusion criteria. R.B. conducted the searches. R.B., M.G.C, Y.K., E.M., F.M., N.P.F and D.A.O. screened the studies and extracted the data and R.B., M.G.C., Y.K., E.M., F.M. and B.Y.N conducted the risk of bias assessment. R.B. synthesised the results and wrote the original draft manuscript with input from L.C., and D.A.O. All authors edited and reviewed the manuscript.

## Acknowledgments

Initial findings from this work were previously presented at the ESCMID Global International Conference, Vienna, Austria, 11^th^-15^th^ April 2025.

We acknowledge members of the UKHSA AMR PROGRESS team for their support in the completion of this study. We also acknowledge the support and advice from members of the UKHSA Knowledge and Library Services.

## Notes

### Competing Interest Statement

The authors have declared no competing interest.

### Funding Statement

The article processing fee will be supported by the ESCMID Study Group for Antimicrobial Stewardship (ESGAP).

