## Supplementary Table for "Measuring appropriateness of antibiotic prescribing in high-income countries: a rapid systematic review of indicators"

### Supplementary information

### S1 - PRISMA checklist

| Section and Topic | Item # | Checklist item | Location where item is reported |
|---|---|---|---|
| **TITLE** | | | |
| Title | 1 | Identify the report as a systematic review. | Line 2 |
| **ABSTRACT** | | | |
| Abstract | 2 | See the PRISMA 2020 for Abstracts checklist. | Line 5 |
| **INTRODUCTION** | | | |
| Rationale | 3 | Describe the rationale for the review in the context of existing knowledge. | Line 66 |
| Objectives | 4 | Provide an explicit statement of the objective(s) or question(s) the review addresses. | Line 71 |
| **METHODS** | | | |
| Eligibility criteria | 5 | Specify the inclusion and exclusion criteria for the review and how studies were grouped for the syntheses. | Figure 1<br>Line 137 |
| Information sources | 6 | Specify all databases, registers, websites, organisations, reference lists and other sources searched or consulted to identify studies. Specify the date when each source was last searched or consulted. | Line 90 |
| Search strategy | 7 | Present the full search strategies for all databases, registers and websites, including any filters and limits used. | Table 1 and Supplementary information table S2 |
| Selection process | 8 | Specify the methods used to decide whether a study met the inclusion criteria of the review, including how many reviewers screened each record and each report retrieved, whether they worked independently, and if applicable, details of automation tools used in the process. | Line 113 |
| Data collection process | 9 | Specify the methods used to collect data from reports, including how many reviewers collected data from each report, whether they worked independently, any processes for obtaining or confirming data from study investigators, and if applicable, details of automation tools used in the process. | Line 120 |
| Data items | 10a | List and define all outcomes for which data were sought. Specify whether all results that were compatible with each outcome domain in each study were sought (e.g. for all measures, time points, analyses), and if not, the methods used to decide which results to collect. | Line 125 |
| | 10b | List and define all other variables for which data were sought (e.g. participant and intervention characteristics, funding sources). Describe any assumptions made about any missing or unclear information. | n/a |
| Study risk of bias assessment | 11 | Specify the methods used to assess risk of bias in the included studies, including details of the tool(s) used, how many reviewers assessed each study and whether they worked independently, and if applicable, details of automation tools used in the process. | Line 133 |
| Effect measures | 12 | Specify for each outcome the effect measure(s) (e.g. risk ratio, mean difference) used in the synthesis or presentation of results. | Line 158 |
| Synthesis | 13a | Describe the processes used to decide which studies were eligible for each synthesis (e.g. tabulating the study intervention characteristics | Line 139 |

| Section and Topic | Item # | Checklist item | Location where item is reported |
|---|---|---|---|
| methods | | and comparing against the planned groups for each synthesis (item #5)). | |
| | 13b | Describe any methods required to prepare the data for presentation or synthesis, such as handling of missing summary statistics, or data conversions. | Line 157 |
| | 13c | Describe any methods used to tabulate or visually display results of individual studies and syntheses. | Line 139 |
| | 13d | Describe any methods used to synthesize results and provide a rationale for the choice(s). If meta-analysis was performed, describe the model(s), method(s) to identify the presence and extent of statistical heterogeneity, and software package(s) used. | n/a |
| | 13e | Describe any methods used to explore possible causes of heterogeneity among study results (e.g. subgroup analysis, meta-regression). | Line 141 |
| | 13f | Describe any sensitivity analyses conducted to assess robustness of the synthesized results. | n/a |
| Reporting bias assessment | 14 | Describe any methods used to assess risk of bias due to missing results in a synthesis (arising from reporting biases). | Line 132 |
| Certainty assessment | 15 | Describe any methods used to assess certainty (or confidence) in the body of evidence for an outcome. | Line 132 |
| **RESULTS** | | | |
| Study selection | 16a | Describe the results of the search and selection process, from the number of records identified in the search to the number of studies included in the review, ideally using a flow diagram. | Figure 2 |
| | 16b | Cite studies that might appear to meet the inclusion criteria, but which were excluded, and explain why they were excluded. | Figure 2 |
| Study characteristics | 17 | Cite each included study and present its characteristics. | Table 2 and Supplementary information Table S3 |
| Risk of bias in studies | 18 | Present assessments of risk of bias for each included study. | Figure 3 and Supplementary information table S4. |
| Results of individual studies | 19 | For all outcomes, present, for each study: (a) summary statistics for each group (where appropriate) and (b) an effect estimate and its precision (e.g. confidence/credible interval), ideally using structured tables or plots. | Table 3 and Table 4. |
| Results of syntheses | 20a | For each synthesis, briefly summarise the characteristics and risk of bias among contributing studies. | Line 255<br>Line 308<br>Line 347 |
| | 20b | Present results of all statistical syntheses conducted. If meta-analysis was done, present for each the summary estimate and its precision (e.g. confidence/credible interval) and measures of statistical heterogeneity. If comparing groups, describe the direction of the effect. | n/a |
| | 20c | Present results of all investigations of possible causes of heterogeneity among study results. | n/a |

| Section and Topic | Item # | Checklist item | Location where item is reported |
|---|---|---|---|
| | 20d | Present results of all sensitivity analyses conducted to assess the robustness of the synthesized results. | n/a |
| Reporting biases | 21 | Present assessments of risk of bias due to missing results (arising from reporting biases) for each synthesis assessed. | Line 449 |
| Certainty of evidence | 22 | Present assessments of certainty (or confidence) in the body of evidence for each outcome assessed. | Line 449 |
| **DISCUSSION** | | | |
| Discussion | 23a | Provide a general interpretation of the results in the context of other evidence. | Line 380 |
| | 23b | Discuss any limitations of the evidence included in the review. | Line 434 |
| | 23c | Discuss any limitations of the review processes used. | Line 434 |
| | 23d | Discuss implications of the results for practice, policy, and future research. | Line 452 |
| **OTHER INFORMATION** | | | |
| Registration and protocol | 24a | Provide registration information for the review, including register name and registration number, or state that the review was not registered. | Line 87 |
| | 24b | Indicate where the review protocol can be accessed, or state that a protocol was not prepared. | n/a |
| | 24c | Describe and explain any amendments to information provided at registration or in the protocol. | Line 92 |
| Support | 25 | Describe sources of financial or non-financial support for the review, and the role of the funders or sponsors in the review. | Line 485 |
| Competing interests | 26 | Declare any competing interests of review authors. | Line 482 |
| Availability of data, code and other materials | 27 | Report which of the following are publicly available and where they can be found: template data collection forms; data extracted from included studies; data used for all analyses; analytic code; any other materials used in the review. | Supplementary information |

**S2 – Search strategies**

*a – Medline search strategy*

| | | |
|---|---|---|
| 1 | *Quality Indicators, Health Care/ | 10016 |
| 2 | *Benchmarking/ | 7885 |
| 3 | metric*.ti,kf. or metric*.ab. /freq=4 | 20479 |
| 4 | criteri*.ti,kf. or criteri*.ab. /freq=4 | 86243 |
| 5 | indicator*.ti,kf. or indicator*.ab. /freq=4 | 72575 |
| 6 | survey*.ti,kf. or survey*.ab. /freq=4 | 250815 |
| 7 | measur*.ti,kf. or measur*.ab. /freq=4 | 610614 |
| 8 | estimat*.ti,kf. or estimat*.ab. /freq=4 | 214309 |
| 9 | *Inappropriate Prescribing/ | 3071 |
| 10 | *Medication Errors/ae, mt, nu, pc, st, sn, td [Classification, Methods, Nursing, Prevention & Control, Standards, Statistics & Numerical Data, Trends] | 6027 |
| 11 | *Prescription Drug Overuse/ | 264 |
| 12 | ((inappropriat* or appropriat* or inadequate or adequate or compliant or incorrect or correct or incorrect or concordant or valid or invalid or "evidence-based" or ineffective or effective or optimal or targeted or timely or reduc* or decreas* or increas* or improv* or rational or irrational) adj2 (prescrib* or prescription* or "use" or "usage" or consumpt* or utili*)).tw,kf. | 309799 |
| 13 | 9 or 10 or 11 or 12 | 316573 |
| 14 | exp Anti-Bacterial Agents/ad, an, dt, st, sn, sd, tu, th, ur [Administration & Dosage, Analysis, Drug Therapy, Standards, Statistics & Numerical Data, Supply & Distribution, Therapeutic Use, Therapy, Urine] | 394204 |
| 15 | *Anti-Infective Agents/ad, st, sd, tu, ur [Administration & Dosage, Standards, Supply & Distribution, Therapeutic Use, Urine] | 12194 |
| 16 | *Antibiotic Prophylaxis/mt, nu, st, sn, td [Methods, Nursing, Standards, Statistics & Numerical Data, Trends] | 2833 |
| 17 | Audit*.ti,kf. or audit*.ab. /freq=4 | 79729 |
| 18 | assess*.ti,kf. or assess*.ab. /freq=4 | 681119 |
| 19 | defin*.ti,kf. or defin*.ab. /freq=4 | 109766 |
| 20 | ((antibiot* or anti-biot* or anti-infective* or antiinfective* or antibacterial* or anti-bacterial*) adj1 prescri*).tw,kf. | 11878 |
| 21 | 14 or 15 or 16 or 20 | 407842 |
| 22 | knowledge.tw,kf. | 1030439 |
| 23 | attitude*.tw,kf. | 215908 |
| 24 | belief*.tw,kf. | 116639 |
| 25 | 22 or 23 or 24 | 1257736 |
| 26 | 1 or 2 or 3 or 4 or 5 or 6 or 7 or 8 or 17 or 18 or 19 | 1993561 |
| 27 | 13 and 21 and 26 | 1621 |
| 28 | 27 not 25 | 1324 |
| 29 | limit 28 to yr="2014 -Current" | 1007 |

*b – Embase search strategy*

| | | |
|---|---|---|
| 1 | metric*.ti,kf. or metric*.ab. /freq=4 | 26109 |
| 2 | criteri*.ti,kf. or criteri*.ab. /freq=4 | 122886 |
| 3 | indicator*.ti,kf. or indicator*.ab. /freq=4 | 88021 |
| 4 | survey*.ti,kf. or survey*.ab. /freq=4 | 303299 |
| 5 | measur*.ti,kf. or measur*.ab. /freq=4 | 735053 |
| 6 | estimat*.ti,kf. or estimat*.ab. /freq=4 | 253716 |
| 7 | Audit*.ti,kf. or audit*.ab. /freq=4 | 111800 |
| 8 | assess*.ti,kf. or assess*.ab. /freq=4 | 940556 |
| 9 | defin*.ti,kf. or defin*.ab. /freq=4 | 146663 |
| 10 | *potentially inappropriate medication/ | 1814 |
| 11 | *prescribing error/ | 2132 |
| 12 | ((inappropriat* or appropriat* or inadequate or adequate or compliant or incorrect or correct or incorrect or concordant or valid or invalid or "evidence-based" or ineffective or effective or optimal or targeted or timely or reduc* or decreas* or increas* or improv* or rational or irrational) adj2 (prescrib* or prescription* or "use" or "usage" or consumpt* or utili*)).tw,kf. | 429047 |
| 13 | 10 or 11 or 12 | 430213 |
| 14 | *antibiotic agent/ad, dt, im, iv, po, pa [Drug Administration, Drug Therapy, Intramuscular Drug Administration, Intravenous Drug Administration, Oral Drug Administration, Parenteral Drug Administration] | 38433 |
| 15 | *antiinfective agent/ad, do, dt, im, iv, po, pa [Drug Administration, Drug Dose, Drug Therapy, Intramuscular Drug Administration, Intravenous Drug Administration, Oral Drug Administration, Parenteral Drug Administration] | 26452 |
| 16 | *antibiotic prophylaxis/ | 9913 |
| 17 | ((antibiot* or anti-biot* or anti-infective* or antiinfective* or antibacterial* or anti-bacterial*) adj1 prescri*).tw,kf. | 17600 |
| 18 | 14 or 15 or 16 or 17 | 85494 |
| 19 | 1 or 2 or 3 or 4 or 5 or 6 or 7 or 8 or 9 | 2533946 |
| 20 | 13 and 18 and 19 | 1538 |
| 21 | knowledge.tw,kf. | 1266865 |
| 22 | attitude*.tw,kf. | 261715 |
| 23 | belief*.tw,kf. | 139769 |
| 24 | 21 or 22 or 23 | 1535355 |
| 25 | 20 not 24 | 1277 |
| 26 | limit 25 to yr="2014 -Current" | 962 |
| 27 | limit 26 to conference abstracts | 263 |
| 28 | 26 not 27 | 699 |

*c – Cochrane library search strategy*

Search | Search manager | Medical terms (MeSH) | PICO search

Save this search | View/Share saved searches | ? Search help

View fewer lines | Print search history

| # | Search | | | Limits | Results |
|---|---|---|---|---|---|
| #1 | antibiot* | S | MeSH | Limits | 42252 |
| #2 | antibacterial* | | | Limits | 19057 |
| #3 | #1 or #2 | | | Limits | 50838 |
| #4 | Criteri* | | | Limits | 407506 |
| #5 | Metric* | | | Limits | 18422 |
| #6 | Indicator* | | | Limits | 30423 |
| #7 | Survey* | | | Limits | 91598 |
| #8 | estimat* | | | Limits | 129978 |
| #9 | audit* | | | Limits | 17714 |
| #10 | assess* | | | Limits | 754585 |
| #11 | defin* | | | Limits | 174112 |
| #12 | #4 OR #5 Or #6 OR #7 OR #8 OR #9 OR #10 OR #11 | | | Limits | 1153541 |
| #13 | (inappropriat* or appropriat* or inadequate or adequate or compliant or incorrect or correct or incorrect or concordant or valid or invalid or "evidence-based" or ineffective or effective or optimal or targeted or timely or reduc* or decreas* or increas* or improv* or rational or irrational) adj2 (prescrib* or prescription* or "use" or "usage" or consumpt* or utili*) | | | Limits | 2245 |
| #14 | #3 AND #12 AND #13 | | | Limits | 340 |
| #15 | Type a search term or use the S or MeSH buttons to compose | S | MeSH | Limits | N/A |

Clear all

Highlight orphan lines

### S3 – Detailed characteristics of included studies

NS – not specified
n/a – not applicable

| Article | Study design | Setting | Infection | Treatment type | Patient ages | Any specified patients? | Type | Indicators used | How were they chosen? | Were they validated? | Is quantitative data stated? | Percentage of inappropriate antibiotic prescribing |
|---|---|---|---|---|---|---|---|---|---|---|---|---|
| **Abbas et al., 2022 (77) Australia** | Point Prevalence Survey | Hospital, including private settings | NS | Both | NS | n/a | General | Is decision compliant with local, regional, or national guidelines<br>Is the choice correct?<br>Is the dose correct?<br>Is the route correct?<br>Is the duration correct?<br>Is decision in-line with expert opinion - e.g. Microbiologist, Infectious Diseases Consultant etc<br>Has the activity of the antibiotic against the known or suspected organism been considered?<br>Has whether the spectrum of the antibiotic is too broad or too narrow been considered? | Pre-existing Audit tool - NAPS | Yes - in previous study | Yes | 39.70 |
| **Aghdassi et al., 2019 (144) Germany** | Point Prevalence Survey | Hospital, including private settings | NS | Both | NS | n/a | General | Is the duration correct?<br>Has intravenous treatment been reviewed and switched to oral treatment (IVOS) appropriately?<br>Has the treatment been de-escalated?<br>Has the patient got clinical signs of an infection?<br>Has the patient got a confirmed or suspected bacterial infection? | Literature review | Yes - in this study | Yes | 16.70 |
| **Alba Fernandez et al., 2022 (104) Spain** | Retrospective study | Hospital, including private settings | NS | Treatment | Adults | Meropenem prescribing for any indication in non-pregnant patients | General | Has whether the patient is pregnant been considered?<br>Is decision in-line with expert opinion - e.g. Microbiologist, Infectious Diseases Consultant etc<br>Is decision compliant with local, regional, or national guidelines | NS | Yes - in this study | Yes | 30.20 |
| **Arcenillas et al., 2018 (6) Spain** | Observational single-center study | Hospital, including private settings | NS | Treatment | NS | n/a | General | Is decision compliant with local, regional, or national guidelines?<br>Have diagnostic blood cultures been taken prior to antibiotic prescribing?<br>Have diagnostic cultures from suspected site of infections been taken prior to antibiotic prescribing?<br>Have cultures been reviewed and treatment adapted accordingly?<br>Has the treatment been de-escalated?<br>Has the patient got a confirmed or suspected bacterial infection?<br>Has the patient got clinical signs of an infection?<br>Has the patient's renal function been considered?<br>Has appropriate Therapeutic Drug Monitoring taken place? | Literature review | Yes - in previous study | Yes | 12.20 |
| **Arnoldo et al., 2019(132) Italy** | Repeated point prevalence surveys | Hospital, including private settings | Healthcare Associated Infections | Prophylaxis | Adults and children | n/a | General | Is the duration correct? | NS | Yes - in previous study | Yes | 43.90 |

| | | | | | | | | | | | |
|---|---|---|---|---|---|---|---|---|---|---|---|
| **Asquier-Khati et al., 2023 (63)<br>France** | A four-step RAND-modified Delphi procedure | Long term care or residential care setting | NS | Treatment | Adults | n/a | Proxy | Patients prescribed the first line choice for the specified indication<br>Patients prescribed a fluoroquinolone within 6 months of another prescription for a fluoroquinolone<br>Seasonality of total antibiotic use<br>Seasonality of amoxicillin-clavulanate use<br>Broad/Narrow ratio<br>Duration greater than specified number of days<br>Patients prescribed antibiotic and non-steroidal anti-inflammatory drug (NSAID) on the same day<br>Intravenous antibiotics | Delphi consensus | Yes - in this study | n/a |
| **Baclet et al., 2022 (64)<br>France** | A qualitative, multicenter, focus-group-based study | Hospital, including private settings | NS | Treatment | Older adults >75 | Hospitalised older adults | General | Is the choice correct?<br>Has the patient got clinical signs of an infection?<br>Has the patient's gender been considered?<br>Is the route correct?<br>Has an inappropriate combination been prescribed?<br>Has the patient got a confirmed or suspected bacterial infection?<br>Have diagnostic cultures from suspected site of infections been taken prior to antibiotic prescribing?<br>Has the patient's severity criteria been considered?<br>Has the patient's previous antibiotic use been considered?<br>Has the patient's renal function been considered?<br>Has the patient's weight been considered?<br>Is the dose correct?<br>Is the duration correct?<br>Has whether there is any duplication, overlap or redundancy in the regimen been considered?<br>Has whether combination or co-prescribing is needed been considered?<br>Has appropriate Therapeutic Drug Monitoring taken place? | Expert group | NS | No |
| **Baclet et al., 2024 (65)<br>France** | Expert Consensus Study | Hospital, including private settings | NS | Treatment | Older adults >75 | Hospitalised older adults | General | Is the choice correct?<br>Has the patient got clinical signs of an infection?<br>Has the patient got a confirmed or suspected bacterial infection?<br>Have relevant severity criteria been met?<br>Is the patient at risk for an infection?<br>Is the patient classified as palliative, meaning that antibiotics may not be appropriate?<br>Has the patient's immune status including immunosuppressing medicines been considered?<br>Has the activity of the antibiotic against the known or suspected organism been considered?<br>Have diagnostic cultures from suspected site of infections been taken prior to antibiotic prescribing?<br>Is the timing of administration correct?<br>Are antibiotics indicated?<br>Has the patient's severity criteria been considered?<br>Has the patient's previous antibiotic use been considered?<br>Is the dose correct?<br>Has the patient's renal function been considered?<br>Is the duration correct?<br>Has an inappropriate combination been prescribed?<br>Has whether there is any duplication, overlap or redundancy in the regimen been considered?<br>Has appropriate Therapeutic Drug Monitoring taken place?<br>Has whether the IV or oral route is appropriate based on specified criteria been considered?<br>Is the route correct?<br>Has whether combination or co-prescribing is needed been | Delphi consensus | Yes - in this study | No |

| | | | | | | | | | | | | |
|---|---|---|---|---|---|---|---|---|---|---|---|---|
| | | | | | | | | considered?<br>Has the patient's gender been considered? | | | | |
| **Barnett, et al. 2020 (14) United States** | Quality Improvement study | Hospital, including private settings | NS | Both | NS | Patients prescribed oral antibiotics at discharge | General | Is the choice correct?<br>Is the dose correct?<br>Is the duration correct?<br>Is decision in-line with decision support tools<br>Is decision compliant with local, regional, or national guidelines?<br>Is decision in-line with expert opinion - e.g. Microbiologist, Infectious Diseases Consultant etc | NS | NS | Yes | 6.70 |
| **Barrie et al., 2018 (47) United Kingdom** | Peer review of cases | Hospital, including private settings | Acute Pancreatitis | Both | Adults | n/a | General | Are antibiotics indicated?<br>Is the timing of administration correct?<br>Is the duration correct?<br>Is decision in-line with expert opinion - e.g. Microbiologist, Infectious Diseases Consultant etc | NS | Yes - in this study | Yes | 19.30 |
| **Barstow et al., 2020 (15) United States** | Retrospective chart review | Hospital, including private settings | NS | Treatment | Children | Patients prescribed an oral antibiotic on discharge from the Emergency Department | General | Is the dose correct?<br>Has the indication being treated been considered?<br>Has the patient's weight been considered?<br>Is decision compliant with local, regional, or national guidelines | Literature review | No | Yes | 40.00 |
| **Berrevoets et al., 2017 (91) Netherlan ds** | Observational pilot study | Hospital, including private settings | NS | Both | NS | n/a | General | Have diagnostic blood cultures been taken prior to antibiotic prescribing?<br>Have diagnostic cultures from suspected site of infections been taken prior to antibiotic prescribing?<br>Is decision compliant with local, regional, or national guidelines?<br>Has the patient's renal function been considered?<br>Has there been appropriate documentation in the clinical record?<br>Have cultures been reviewed and treatment adapted accordingly?<br>Has intravenous treatment been reviewed and switched to oral treatment (IVOS) appropriately?<br>Has appropriate Therapeutic Drug Monitoring taken place?<br>Has the treatment been stopped if there is no evidence of infection?<br>Is the patient taking it? | NS | NS | No | |
| **Bjerrum et al., 2022 (114) France, Poland, Greece, Spain, and Lithuania** | Intervention implementation study protocol | General practice, out of hours services, nursing homes and community pharmacies | Respiratory tract infections and urinary tract infections | Both | NS | n/a | General | Has the patient got a confirmed or suspected bacterial infection?<br>Has the activity of the antibiotic against the known or suspected organism been considered?<br>Have relevant diagnostic criteria been met?<br>Is decision compliant with local, regional, or national guidelines | NS | No | No | |
| **Black et al., 2018(124) Canada** | Point prevalence survey | Hospital, including private settings | NS | Both | NS | n/a | General | Is decision compliant with local, regional, or national guidelines?<br>Is the choice correct?<br>Is the dose correct?<br>Is the duration correct?<br>Is decision in-line with expert opinion - e.g. Microbiologist, Infectious Diseases Consultant etc | NS | No | Yes | 24.20 |
| **Bohan et al., 2019 (16) United States** | A multicenter quality improvement evaluation | Primary, community setting or | Uncomplicate d Respiratory tract infections | Treatment | Adults | Veterans | General | Is decision compliant with local, regional, or national guidelines?<br>Have relevant diagnostic tests been carried out?<br>Has the patient's allergy status been considered? | Using treatment or | NS | Yes | 61.00 |

| | | | | | | | | | | | |
|---|---|---|---|---|---|---|---|---|---|---|---|
| | | ambulatory care | | | | | | Is the choice correct?<br>Have relevant diagnostic criteria been met?<br>Has the patient got clinical signs of an infection?<br>Has the indication being treated been considered?<br>Has there been appropriate documentation in the clinical record? | diagnostic guidelines | | | |
| **Burns et al., 2020 (17) United States** | Retrospective cohort study | Primary, community setting or ambulatory care | Upper respiratory tract infections and urinary tract infections | Treatment | NS | n/a | General | Has the indication being treated been considered?<br>Is the choice correct?<br>Is the duration correct?<br>Is decision compliant with local, regional, or national guidelines | NS | Yes - in this study | Yes | 68.4 |
| **Cameron et al., 2015 (48) United Kingdom** | Audit pre and post intervention | Hospital, including private settings | Prophylaxis of gastro surgical infection | Prophylaxis | NS | Gastro-surgical patients | General | Is decision compliant with local, regional, or national guidelines | Using treatment or diagnostic guidelines | Yes - in this study | Yes | 66.70 |
| **Canoui et al., 2018 (49) United Kingdom** | Letter to editor | Hospital, including private settings | NS | Treatment | NS | Carbapenem prescribing for any infection | General | Is decision compliant with local, regional, or national guidelines?<br>Has the patient's severity criteria been considered?<br>Have the patient's risk factors for resistant bacteria been considered? | Expert group | Yes - in this study | Yes | 37.50 |
| **Cao et al., 2016 (18) United States** | Prospective review | Hospital, including private settings | NS | NS | NS | n/a | General | Is the choice correct?<br>Is the dose correct?<br>Have interactions with the patient's other medicines or feeds been considered?<br>Has the treatment been monitored?<br>Has intravenous treatment been reviewed and switched to oral treatment (IVOS) appropriately?<br>Is decision in-line with expert opinion - e.g. Microbiologist, Infectious Diseases Consultant etc<br>Has whether there is any duplication, overlap or redundancy in the regimen been considered?<br>Has the activity of the antibiotic against the known or suspected organism been considered? | NS | No | No | |
| **Castel et al., 2016 (66) France** | Monocentric descriptive retrospective study | Hospital, including private settings | NS | Treatment | NS | Daptomycin treatment for any infection (more than 2 days) | General | Is decision compliant with local, regional, or national guidelines?<br>Is the route correct?<br>Has the patient's renal function been considered?<br>Has the patient's previous antibiotic use been considered?<br>Has the patient's allergy status been considered?<br>Is decision in-line with expert opinion - e.g. Microbiologist, Infectious Diseases Consultant etc<br>Is the dose correct?<br>Is the duration correct?<br>Has whether combination or co-prescribing is needed been considered? | Using treatment or diagnostic guidelines | Yes - in this study | Yes | 85.00 |
| **Catho et al., 2018 (153) Switzerland** | Study protocol for randomised multicentre trial | Hospital, including private settings | NS | NS | NS | n/a | General | Is decision compliant with local, regional, or national guidelines?<br>Has intravenous treatment been reviewed and switched to oral treatment (IVOS) appropriately?<br>Has the treatment been de-escalated?<br>Have relevant diagnostic criteria been met?<br>Is the choice correct?<br>Is the duration correct? | Literature review | Yes - in previous study | No | |
| **Cattani et al., 2020 (133) Italy** | Prospective multicentre study | Hospital, including | Community Acquired Pneumonia | Treatment | Adults | n/a | General | Is decision compliant with local, regional, or national guidelines?<br>Is the choice correct? | Using treatment or | Yes - in previous study | Yes | 77.55 |

| | | | | | | | | | | | | |
|---|---|---|---|---|---|---|---|---|---|---|---|---|
| | | private settings | and Hospital Acquired Pneumonia | | | | | Is the dose correct?<br>Is the duration correct? | diagnostic guidelines | | | |
| **Chen et al., 2023 (125) Canada** | Prevalence Audit | Hospital, including private settings | NS | Treatment | NS | Ciprofloxacin use | General | Is decision compliant with local, regional, or national guidelines?<br>Has the indication being treated been considered?<br>Has the activity of the antibiotic against the known or suspected organism been considered?<br>Is the route correct?<br>Is the dose correct?<br>Has the patient's allergy status been considered?<br>Has the patient got clinical signs of an infection?<br>Is the duration correct? | Using treatment or diagnostic guidelines | Yes - in this study | Yes | 69.00 |
| **Choi et al., 2021 (19) United States** | Retrospective, quasi-experimental study | Primary, community setting or ambulatory care | Urinary tract Infection and Skin and soft tissue infections | Treatment | NS | n/a | General | Is decision compliant with local, regional, or national guidelines?<br>Is the choice correct?<br>Is the dose correct?<br>Is the duration correct?<br>Have relevant diagnostic criteria been met?<br>Has the patient got clinical signs of an infection?<br>Has whether the patient is pregnant been considered? | Using treatment or diagnostic guidelines | NS | Yes | 72.50 |
| **Chopra et al., 2014 (50) United Kingdom** | Audit pre and post intervention - single centre | Hospital, including private settings | Dental infections | Treatment | NS | Patients in the dental department | General | Is decision compliant with local, regional, or national guidelines?<br>Has the patient got clinical signs of an infection?<br>Is the patient at risk for an infection?<br>Are antibiotics indicated?<br>Is the dose correct?<br>Is the duration correct?<br>Is the frequency correct? | Using treatment or diagnostic guidelines | Yes - in this study | Yes | 70.00 |
| **Chorafa et al., 2021 (160) Greece** | Audit pre and post intervention - multicentre | Hospital, including private settings | Prophylaxis of surgical infection | Prophylaxis | NS | Patient undergoing specified procedures | General | Is the choice correct?<br>Is the timing of administration correct?<br>Is the duration correct?<br>Is decision compliant with local, regional, or national guidelines? | Using treatment or diagnostic guidelines | Yes - in this study | Yes | 71.80 |
| **Clegg et al., 2021 (20) United States** | Cluster randomized trial | Primary, community setting or ambulatory care | Acute Respiratory Tract Infection | Treatment | Children | n/a | Proxy | Rate of patients prescribed an antibiotic based on the specified indication<br>Patients prescribed the first line choice for the specified indication | Using treatment or diagnostic guidelines | Yes - in this study | n/a | |
| **Colombo et al., 2025 (21) United States** | Retrospective analysis | Long term care or residential care setting | Urinary tract infections or Respiratory tract infections | Treatment | Adults | Nursing home residents | General | Have relevant diagnostic criteria been met? | Using treatment or diagnostic guidelines | Yes - in this study | Yes | 69.00 |
| **Cona et al., 2021 (134) Italy** | Point prevalence survey | Hospital, including private settings | NS | NS | NS | Patients within the medical department | General | Is decision in-line with expert opinion - e.g. Microbiologist, Infectious Diseases Consultant etc<br>Is the dose correct?<br>Is the duration correct?<br>Has whether the antibiotic will penetrate to the target site been considered?<br>Is the frequency correct?<br>Have cultures been reviewed and treatment adapted accordingly?<br>Is decision compliant with local, regional, or national guidelines? | NS | no | Yes | 52.00 |
| **Cotta et al., 2014 (78) Australia** | Point Prevalence Survey | Hospital, including private settings | NS | Both | NS | n/a | General | Is decision in-line with expert opinion - e.g. Microbiologist, Infectious Diseases Consultant etc<br>Is decision compliant with local, regional, or national guidelines? | Literature review | Yes - in this study | Yes | 27.00 |

| | | | | | | | | | | | | |
|---|---|---|---|---|---|---|---|---|---|---|---|---|
| | | | | | | | | Is the choice correct?<br>Is the dose correct?<br>Is the frequency correct?<br>Is the duration correct?<br>Has the patient's allergy status been considered?<br>Have the patient's contraindications been considered?<br>Have cultures been reviewed and treatment adapted accordingly? | | | | |
| **Datta et al., 2019 (22) United States** | Cohort Study | Hospital, including private settings | Urinary tract infection | Treatment | NS | Advanced cancer patients transitionin g to comfort measures | General | Have relevant diagnostic criteria been met? | Literature review | Yes - in this study | Yes | 14.00 |
| **Degnan et al., 2022 (23) United States** | Longitudinal descriptive study | Primary, community setting or ambulatory care | Respiratory Tract Infections | Treatment | NS | n/a | General | Have relevant diagnostic criteria been met?<br>Is decision compliant with local, regional, or national guidelines? | Using treatment or diagnostic guidelines | Yes - in this study | Yes | 69.00 |
| **Denny et al., 2019 (79) Australia** | Retrospective observational study | Hospital, including private settings | NS | Both | NS | All patients who presented to the Emergency Department and were prescribed at least one antibiotic. | General | Is decision compliant with local, regional, or national guidelines?<br>Is the choice correct?<br>Is the dose correct?<br>Is the route correct?<br>Is the duration correct?<br>Is decision in-line with expert opinion - e.g. Microbiologist, Infectious Diseases Consultant etc<br>Has the activity of the antibiotic against the known or suspected organism been considered?<br>Has whether the spectrum of the antibiotic is too broad or too narrow been considered?<br>Has whether there is any duplication, overlap or redundancy in the regimen been considered?<br>Has the patient's allergy status been considered?<br>Are antibiotics indicated? | Pre-existing Audit tool - NAPS | Yes - in previous study | Yes | 34.20 |
| **Dentan et al., 2017 (67) France** | Retrospective study | Hospital, including private settings | NS | Both | NS | All patients prescribed linezolid. | General | Is decision compliant with local, regional, or national guidelines?<br>Is decision compliant with local, regional, or national guidelines?<br>Is decision in-line with evidence-base or scientific information<br>Is decision in-line with expert opinion - e.g. Microbiologist, Infectious Diseases Consultant etc<br>Has the patient got clinical signs of an infection?<br>Has whether the spectrum of the antibiotic is too broad or too narrow been considered?<br>Has the activity of the antibiotic against the known or suspected organism been considered?<br>Has the mode of action of the antibiotic been considered?<br>Have the patient's contraindications been considered?<br>Is the dose correct?<br>Is the route correct?<br>Has intravenous treatment been reviewed and switched to oral treatment (IVOS) appropriately?<br>Has the treatment been monitored? | Literature review | Yes - in this study | Yes | 77.00 |
| **DePestel et al. 2014 (24) United States** | Observational, retrospective, cohort study | Hospital, including private settings | NS | Treatment | NS | Patients receiving selected | General | Is decision compliant with local, regional, or national guidelines?<br>Have cultures been reviewed and treatment adapted accordingly? | Literature review | Yes - in this study | Yes | 21.00 |

| | | | | | | | | | | | | |
|---|---|---|---|---|---|---|---|---|---|---|---|---|
| | | | | | | antimicrobials. | | Is decision in-line with evidence-base or scientific information<br>Is decision in-line with expert opinion - e.g. Microbiologist, Infectious Diseases Consultant etc<br>Has the patient got a confirmed or suspected bacterial infection?<br>Is the choice correct? | | | | |
| **Devchand et al., 2019 (80) Australia** | Point Prevalence Survey | Hospital, including private settings | NS | Both | NS | Patients in Intensive care | General | Is decision compliant with local, regional, or national guidelines?<br>Is the choice correct?<br>Is the dose correct?<br>Is the route correct?<br>Is the duration correct?<br>Is decision in-line with expert opinion - e.g. Microbiologist, Infectious Diseases Consultant etc<br>Has the activity of the antibiotic against the known or suspected organism been considered?<br>Has whether the spectrum of the antibiotic is too broad or too narrow been considered?<br>Has whether there is any duplication, overlap or redundancy in the regimen been considered?<br>Has the patient's allergy status been considered?<br>Are antibiotics indicated? | Pre-existing Audit tool - NAPS | Yes - in previous study | Yes | 53.30 |
| **Doyle et al., 2021 (126) Canada** | Pre and post intervention Point Prevalence Survey | Hospital, including private settings | NS | Treatment | NS | n/a | General | Is decision compliant with local, regional, or national guidelines?<br>Is the choice correct?<br>Is the dose correct?<br>Is the route correct?<br>Is the duration correct?<br>Is decision in-line with expert opinion - e.g. Microbiologist, Infectious Diseases Consultant etc<br>Has the activity of the antibiotic against the known or suspected organism been considered?<br>Has whether the spectrum of the antibiotic is too broad or too narrow been considered?<br>Has whether there is any duplication, overlap or redundancy in the regimen been considered?<br>Has the patient's allergy status been considered?<br>Are antibiotics indicated? | Pre-existing Audit tool - NAPS | Yes - in previous study | Yes | 44.90 |
| **Dresser et al., 2018 (127) Canada** | Delphi consensus | Hospital, including private settings | NS | treatment | NS | Patients in critical care | General | Has the patient got a confirmed or suspected bacterial infection?<br>Has whether the spectrum of the antibiotic is too broad or too narrow been considered?<br>Has the patient's allergy status been considered?<br>Have the patient's contraindications been considered?<br>Is the route correct?<br>Is the dose correct?<br>Is the frequency correct?<br>Has whether the antibiotic will penetrate to the target site been considered?<br>Is decision in-line with evidence-base or scientific information<br>Is the duration correct?<br>Has whether there is any duplication, overlap or redundancy in the regimen been considered?<br>Are antibiotics indicated?<br>Has the treatment been stopped if there is no evidence of infection? | Delphi consensus | No | No | |

| | | | | | | | | | | | | |
|---|---|---|---|---|---|---|---|---|---|---|---|---|
| **Dulku et al., 2022 (128) Canada** | Retrospective chart review | Hospital, including private settings | NS | Treatment | Adults | Patients on treatment with meds with high PO bioavailability | General | Have interactions with the patient's other medicines or feeds been considered?<br>Has whether the IV or oral route is appropriate based on specified criteria been considered?<br>Have diagnostic blood cultures been taken prior to antibiotic prescribing?<br>Has intravenous treatment been reviewed and switched to oral treatment (IVOS) appropriately? | Using treatment or diagnostic guidelines | Yes - in this study | Yes | 57.90 |
| **Durkin et al., 2018 (25) United States** | Longitudinal descriptive study | Dental | Dental infections | Both | Adults | n/a | General | Are antibiotics indicated?<br>Is decision compliant with local, regional, or national guidelines?<br>Is the duration correct?<br>Is decision in-line with expert opinion - e.g. Microbiologist, Infectious Diseases Consultant etc | Using treatment or diagnostic guidelines | NS | Yes | 13.70 |
| **Durkin et al., 2018 (26) United States** | Retrospective observational cohort study | Primary, community setting or ambulatory care | Urinary tract infection | Treatment | Adults | Non-pregnant women aged 18-45 | General | Has the patient's gender been considered?<br>Has whether the patient is pregnant been considered?<br>Is decision compliant with local, regional, or national guidelines?<br>Is the choice correct?<br>Has an inappropriate combination been prescribed?<br>Is the duration correct? | Using treatment or diagnostic guidelines | Yes - in this study | Yes | 49.78 |
| **Erturk Sengal et al., 2019 (169) Türkiye** | Prospective quasi-experimental study | Hospital, including private settings | NS | Both | NS | Patients on intravenous therapy for more than 72 hours | General | Is decision compliant with local, regional, or national guidelines?<br>Is the choice correct?<br>Is the dose correct?<br>Has whether there is any duplication, overlap or redundancy in the regimen been considered?<br>Has whether the spectrum of the antibiotic is too broad or too narrow been considered?<br>Are antibiotics indicated? | Literature review | Yes - in previous study | Yes | 33.00 |
| **Eure et al., 2017 (27) United States** | Point Prevalence Survey | Long term care or residential care setting | Urinary tract infection | Both | Adults | Nursing home residents | General | Have relevant diagnostic criteria been met? | Literature review | Yes - in previous study | No | |
| **Fleming et al., 2015 (115) United Kingdom and Ireland** | Comparison of surveys | Hospital, including private settings | NS | Both | NS | n/a | General | Has whether the spectrum of the antibiotic is too broad or too narrow been considered?<br>Have diagnostic cultures from suspected site of infections been taken prior to antibiotic prescribing?<br>Has the treatment been de-escalated?<br>Is decision compliant with local, regional, or national guidelines? | NS | NS | No | |
| **Fleming-Dutra et al., 2016 (28) United States** | Comparison of prescribing | Primary, community setting or ambulatory care | Various | Treatment | Adults and children | n/a | Proxy | Rate of patients prescribed an antibiotic based on the indication | Expert group | NS | n/a | |
| **Forst et al., 2024 (145) Germany** | Multi-centre interventional study | Hospital, including private settings | NS | Both | NS | n/a | General | Have diagnostic blood cultures been taken prior to antibiotic prescribing?<br>Have diagnostic cultures from suspected site of infections been taken prior to antibiotic prescribing?<br>Has the indication being treated been considered?<br>Has the patient got a confirmed or suspected bacterial infection?<br>Is the dose correct?<br>Has the patient's renal function been considered?<br>Is the choice correct?<br>Has whether the spectrum of the antibiotic is too broad or too narrow been considered?<br>Is the duration correct? | NS | Yes - in this study | No | |

| | | | | | | | | | | | | |
|---|---|---|---|---|---|---|---|---|---|---|---|---|
| | | | | | | | | Has intravenous treatment been reviewed and switched to oral treatment (IVOS) appropriately?<br>Has there been appropriate documentation in the clinical record? | | | | |
| **Franchi et al., 2021(135) Italy** | Logistic regression analyses of prescribing data | Primary, community setting or ambulatory care | NS | Both | Adults | n/a | Proxy | Penicillins plus beta-lactamase inhibitor<br>3rd and 4th generation cephalosporin<br>Fluoroquinolones<br>Patients co-prescribed a fluoroquinolone and steroid | Pre-existing indicator - ESAC | Yes - in previous study | n/a | |
| **Friedman et al., 2020 (81) Australia** | Pilot and evaluation | Primary, community setting or ambulatory care | NS | Treatment | NS | Patients on Outpatient Antimicrobial Therapy (OPAT) | General | Is decision compliant with local, regional, or national guidelines?<br>Is the choice correct?<br>Is the dose correct?<br>Is the route correct?<br>Is the duration correct?<br>Is decision in-line with expert opinion - e.g. Microbiologist, Infectious Diseases Consultant etc<br>Has the activity of the antibiotic against the known or suspected organism been considered?<br>Has whether the spectrum of the antibiotic is too broad or too narrow been considered?<br>Has whether there is any duplication, overlap or redundancy in the regimen been considered?<br>Has the patient's allergy status been considered?<br>Are antibiotics indicated? | Pre-existing Audit tool - NAPS | NS | Yes | 11.00 |
| **Garcia-Sangenis et al., 2024 (116) France, Greece, Lithuania, Poland, and Spain** | Before-and-after study | General practice, out-of-hours services, nursing homes, and community pharmacies | NS | Both | NS | n/a | General | Are antibiotics indicated?<br>Is the choice correct?<br>Has the indication being treated been considered?<br>Is decision compliant with local, regional, or national guidelines? | Expert group | Yes - in this study | Yes | 72.20 |
| **Gardiner et al., 2020 (174) New Zealand** | Point Prevalence Survey | Hospital, including private settings | NS | Both | Adults | n/a | General | Has there been appropriate documentation in the clinical record?<br>Is decision compliant with local, regional, or national guidelines?<br>Is the choice correct?<br>Is the dose correct?<br>Is the route correct?<br>Is the duration correct?<br>Is decision in-line with expert opinion - e.g. Microbiologist, Infectious Diseases Consultant etc<br>Has the activity of the antibiotic against the known or suspected organism been considered?<br>Has whether the spectrum of the antibiotic is too broad or too narrow been considered?<br>Has whether there is any duplication, overlap or redundancy in the regimen been considered?<br>Has the patient's allergy status been considered?<br>Are antibiotics indicated? | Pre-existing Audit tool - NAPS | Yes - in previous study | Yes | 16.90 |
| **Garlasco et al., 2024 (136) Italy** | Point Prevalence Survey | Hospital, including private settings | NS | both | NS | n/a | General | Has there been appropriate documentation in the clinical record? | NS | Yes - in previous study | Yes | 23.50 |
| **Gharbi et al., 2016 (51) United Kingdom** | Cross-sectional study | Hospital, including private settings | NS | Both | Children | n/a | Proxy | Rate of patients prescribed an antibiotic based on the indication<br>Carbapenems and piperacillin-tazobactam | Pre-existing indicator - | No | n/a | |

| | | | | | | | | | | | | |
|---|---|---|---|---|---|---|---|---|---|---|---|---|
| | | | | | | | | | National CQUIN | | | |
| **Gimenez-Perez et al., 2024 (105) Spain** | Prospective multicentre study | Hospital, including private settings | Urinary source E. coli bacteraemia | Treatment | Adults | n/a | General | Is decision compliant with local, regional, or national guidelines?<br>Have cultures been reviewed and treatment adapted accordingly? | Using treatment or diagnostic guidelines | No | Yes | 11.90 |
| **Glinz et al., 2017 (154) Switzerland** | Nationwide Audit survey | Primary, community setting or ambulatory care | Respiratory tract infections and urinary tract infections | Treatment | NS | n/a | Proxy | Rate of patients prescribed an antibiotic based on the indication<br>Patients prescribed the first line choice for the specified indication<br>Patients prescribed a fluoroquinolone for a specified indication | Pre-existing indicator - ESAC | Yes - in this study | n/a | |
| **Gurtler et al., 2019 (155) Switzerland** | Repeated point prevalence survey | Hospital, including private settings | NS | Both | NS | n/a | General | Has the patient got clinical signs of an infection?<br>Is the patient at risk for an infection?<br>Has the activity of the antibiotic against the known or suspected organism been considered?<br>Is the dose correct?<br>Has the patient's allergy status been considered?<br>Has intravenous treatment been reviewed and switched to oral treatment (IVOS) appropriately?<br>Is decision compliant with local, regional, or national guidelines?<br>Have cultures been reviewed and treatment adapted accordingly?<br>Has appropriate Therapeutic Drug Monitoring taken place?<br>Has there been appropriate documentation in the clinical record?<br>Has the patient's travel history been considered?<br>Is the frequency correct?<br>Is the duration correct?<br>Has the patient's renal function been considered?<br>Has the patient's weight been considered?<br>Has the indication being treated been considered? | Using treatment or diagnostic guidelines | Yes - in previous study | Yes | 33.20 |
| **Gutierrez-Urbon et al., 2024 (106) Spain** | Observational, multicenter, cross-sectional study | Hospital, including private settings | NS | both | NS | n/a | General | Has the indication being treated been considered?<br>Is the choice correct?<br>Is the timing of administration correct?<br>Is the dose correct?<br>Is the route correct?<br>Is the duration correct?<br>Has the patient's risk of adverse events been considered?<br>Has there been appropriate documentation in the clinical record?<br>Is the frequency correct?<br>Has the treatment been effective? | Literature review | Yes - in this study | Yes | 21.00 |
| **Guzik et al., 2019 (29) United States** | Observational study | Primary, community setting or ambulatory care | Acute Respiratory Infection | Treatment | NS | n/a | Proxy | Rate of patients prescribed an antibiotic based on the indication | Literature review | Yes - in this study | n/a | |
| **Hahn et al., 2019 (30) United States** | Longitudinal descriptive study | Hospital, including private settings | NS | Treatment | Adults | Intravenous starts in haemodialysis patients | General | Have diagnostic blood cultures been taken prior to antibiotic prescribing?<br>Has the patient got a confirmed or suspected bacterial infection? | Using treatment or diagnostic guidelines | No | Yes | 57.50 |
| **Hearsey et al., 2022 (52) United Kingdom** | Single Centre Retrospective Audit | Hospital, including private settings | NS | Treatment | NS | Surgical patients | General | Is decision in-line with expert opinion - e.g. Microbiologist, Infectious Diseases Consultant etc<br>Are antibiotics indicated? | Pre-existing Audit tool - UK | No | Yes | 16.50 |

| | | | | | | | | | | | | |
|---|---|---|---|---|---|---|---|---|---|---|---|---|
| | | | | | | | | Has the activity of the antibiotic against the known or suspected organism been considered?<br>Has the treatment been stopped if there is no evidence of infection?<br>Is the duration correct?<br>Has whether there is any duplication, overlap or redundancy in the regimen been considered? | national audit tool | | | |
| **Higgins et al., 2023 (53) United Kingdom** | point prevalence survey | Hospital, including private settings | Community-acquired infections and surgical antibiotic prophylaxis | Both | NS | n/a | General | Is the choice correct?<br>Is the duration correct?<br>Is decision compliant with local, regional, or national guidelines? | Expert group | Yes - in this study | Yes | 30.00 |
| **Hogli et al., 2016 (166) Norway** | Audit and feedback intervention study | Hospital, including private settings | Community Acquired Pneumonia and Infective Exacerbation of COPD | Treatment | NS | n/a | General | Is decision compliant with local, regional, or national guidelines?<br>Is the choice correct?<br>Is the dose correct?<br>Is the duration correct?<br>Has whether combination or co-prescribing is needed been considered? | Using treatment or diagnostic guidelines | Yes - in this study | Yes | 38.30 |
| **Hood et al., 2019 (9) United Kingdom** | RAND-modified Delphi process | Hospital, including private settings | NS | Treatment | NS | n/a | General | Is decision in-line with expert opinion - e.g. Microbiologist, Infectious Diseases Consultant etc<br>Are antibiotics indicated?<br>Has the activity of the antibiotic against the known or suspected organism been considered?<br>Has the treatment been stopped if there is no evidence of infection?<br>Is the duration correct?<br>Has whether there is any duplication, overlap or redundancy in the regimen been considered? | Delphi consensus | Yes - in this study | No | |
| **Hussein et al., 2017 (146) Germany** | Literature review and Delphi process | Dental | Dental infections | Both | NS | n/a | Proxy | Rate of patients prescribed an antibiotic based on the indication<br>Penicillin<br>Clindamycin | Delphi consensus | Yes - in this study | n/a | |
| **Ierano et al., 2020 (82) Australia** | Multicenter, national, quality improvement study | Hospital, including private settings | Prophylaxis of orthopaedic surgical infection | Prophylaxis | NS | Orthopaedic surgical patients | General | Is decision in-line with expert opinion - e.g. Microbiologist, Infectious Diseases Consultant etc<br>Has the patient's allergy status been considered?<br>Have cultures been reviewed and treatment adapted accordingly?<br>Is the dose correct?<br>Is the route correct?<br>Is the timing of administration correct?<br>Is the duration correct?<br>Has whether the spectrum of the antibiotic is too broad or too narrow been considered?<br>Are antibiotics indicated?<br>Is the frequency correct? | Pre-existing Audit tool - Surgical NAPS | Yes - in previous study | Yes | 35.30 |
| **James et al., 2015 (83) Australia** | Design and pilot of national audit tool | Hospital, including private settings | NS | Both | NS | n/a | General | Has there been appropriate documentation in the clinical record?<br>Have cultures been reviewed and treatment adapted accordingly?<br>Has the patient's allergy status been considered?<br>Is decision compliant with local, regional, or national guidelines?<br>Is the duration correct?<br>Is the route correct?<br>Has whether the spectrum of the antibiotic is too broad or too narrow been considered? | Expert group | Yes - in this study | Yes | 39.00 |

| | | | | | | | | | | | | |
|---|---|---|---|---|---|---|---|---|---|---|---|---|
| | | | | | | | | Is decision in-line with expert opinion - e.g. Microbiologist, Infectious Diseases Consultant etc<br>Is the dose correct?<br>Is the frequency correct? | | | | |
| **Jung et al., 2023 (149) Korea** | Cohort study | Inpatient, outpatient, and emergency departments. | Asymptomatic bacteriuria and urinary tract infection | Treatment | NS | n/a | General | Is decision compliant with local, regional, or national guidelines?<br>Have cultures been reviewed and treatment adapted accordingly?<br>Is the duration correct?<br>Is the dose correct?<br>Is the route correct?<br>Has the patient's renal function been considered?<br>Has intravenous treatment been reviewed and switched to oral treatment (IVOS) appropriately?<br>Has the treatment been reviewed or reassessed? | Literature review | NS | Yes | 22.00 |
| **Kallen et al., 2018 (92) Netherlands** | Delphi consensus | Hospital, including private settings | NS | NS | NS | Patients on Intensive care | General | Have diagnostic blood cultures been taken prior to antibiotic prescribing?<br>Has appropriate Therapeutic Drug Monitoring taken place? | Delphi consensus | No | No | |
| **Kern et al., 2024 (171) Austria** | Point prevalence survey | Hospital, including private settings | NS | Both | NS | n/a | General | Is decision compliant with local, regional, or national guidelines?<br>Has the activity of the antibiotic against the known or suspected organism been considered?<br>Is the dose correct?<br>Is the duration correct?<br>Have cultures been reviewed and treatment adapted accordingly?<br>Has there been appropriate documentation in the clinical record? | NS | Yes - in this study | Yes | 31.70 |
| **Khaw et al., 2018 (31) United States** | Cohort study | Hospital, including private settings | Prophylaxis of procedural infection | Prophylaxis | NS | Patients undergoing endoscopic urological procedures. | General | Is decision compliant with local, regional, or national guidelines?<br>Is the choice correct?<br>Have cultures been reviewed and treatment adapted accordingly?<br>Has the patient's allergy status been considered?<br>Is the duration correct?<br>Has there been appropriate documentation in the clinical record?<br>Has the indication being treated been considered?<br>Has the treatment been reviewed or reassessed? | Using treatment or diagnostic guidelines | Yes - in this study | Yes | 57.90 |
| **Kim et al., 2021 (150) Korea** | Delphi consensus | In patients, and outpatients | NS | Both | Adults and children | n/a | General | Is decision compliant with local, regional, or national guidelines?<br>Have cultures been reviewed and treatment adapted accordingly?<br>Have diagnostic cultures from suspected site of infections been taken prior to antibiotic prescribing?<br>Have diagnostic blood cultures been taken prior to antibiotic prescribing?<br>Has the patient's renal function been considered?<br>Has there been appropriate documentation in the clinical record?<br>Is decision compliant with local, regional, or national guidelines?<br>Is the timing of administration correct? | Delphi consensus | No | No | |
| **Kinoshita et al., 2020 (138) Japan** | Quasi-experimental study | Primary, community setting or ambulatory care | NS | Both | Children | n/a | General | Is decision in-line with expert opinion - e.g. Microbiologist, Infectious Diseases Consultant etc | NS | No | Yes | 78.60 |

| | | | | | | | | | | | | |
|---|---|---|---|---|---|---|---|---|---|---|---|---|
| **Komagamine et al., 2019 (139) Japan** | Repeated point prevalence survey | Hospital, including private settings | NS | both | NS | n/a | General | Has the indication being treated been considered?<br>Is the dose correct?<br>Is the duration correct?<br>Is the choice correct?<br>Has the activity of the antibiotic against the known or suspected organism been considered?<br>Is the timing of administration correct? | Literature review | Yes - in previous study | Yes | 58.80 |
| **Komagamine et al., 2019 (140) Japan** | Point Prevalence Survey | Hospital, including private settings | NS | Both | NS | n/a | General | Has the patient got a confirmed or suspected bacterial infection?<br>Is the dose correct?<br>Is the timing of administration correct?<br>Is the duration correct?<br>Has the activity of the antibiotic against the known or suspected organism been considered?<br>Has whether the spectrum of the antibiotic is too broad or too narrow been considered? | Literature review | Yes - in this study | Yes | 38.00 |
| **Kusama, 2024 (141) Japan** | Letter to editor | Primary, community setting or ambulatory care | NS | NS | NS | n/a | Proxy | % ‘Access’ category | Pre-existing indicator - International indicator | NS | n/a | |
| **Lass et al., 2020 (173) Estonia** | Retrospective observational study | Primary, community setting or ambulatory care | NS | both | NS | n/a | Proxy | All penicillins<br>Cephalosporins<br>Macrolides, lincosamides, and streptogramins<br>Fluoroquinolones<br>Beta-lactamase sensitive penicillins<br>Penicillins plus beta-lactamase inhibitor<br>3rd and 4th generation cephalosporin<br>Broad/Narrow ratio<br>Seasonality of total antibiotic use<br>Seasonality of quinolone use | Pre-existing indicator - ESAC | Yes - in previous study | n/a | |
| **Legros et al., 2024 (157) Belgium** | Delphi consensus | Hospital, including private settings | NS | Both | NS | n/a | General | Is the dose correct?<br>Is the route correct?<br>Is the duration correct?<br>Has there been appropriate documentation in the clinical record?<br>Is decision compliant with local, regional, or national guidelines?<br>Is decision compliant with local, regional, or national guidelines?<br>Is the choice correct?<br>Is the timing of administration correct? | Delphi consensus | No | No | |
| **Leslie et al., 2023 (129) Canada** | Cohort Study | Primary, community setting or ambulatory care | Various | Treatment | Adults | n/a | Proxy | Rate of patients prescribed an antibiotic based on the indication | Literature review | Yes - in previous study | n/a | |
| **Levine et al., 2016 (32) United States** | Temporal trend analysis | Primary, community setting or ambulatory care | Various | Treatment | NS | n/a | Proxy | Rate of patients prescribed an antibiotic based on the indication | Literature review | Yes - in this study | n/a | |
| **Liu et al., 2019 (33) United States** | Commentary | Hospital, including private settings | NS | Both | NS | n/a | General | Have cultures been reviewed and treatment adapted accordingly?<br>Has whether the spectrum of the antibiotic is too broad or too narrow been considered?<br>Is the duration correct?<br>Has the patient got a confirmed or suspected bacterial infection? | NS | No | No | |

| | | | | | | | | | | | | |
|---|---|---|---|---|---|---|---|---|---|---|---|---|
| | | | | | | | | Has the patient got clinical signs of an infection?<br>Is decision compliant with local, regional, or national guidelines? | | | | |
| **Lopez-Campos et al., 2015 (117) Austria, Belgium, Croatia, Greece, Ireland, Malta, Poland, Romania, Slovakia, Spain, Switzerland, Türkiye, and United Kingdom.** | Audit | Hospital, including private settings | Acute exacerbations of Chronic Obstructive Pulmonary Disease | Treatment | Adults | n/a | General | Have relevant diagnostic criteria been met?<br>Is decision compliant with local, regional, or national guidelines? | Literature review | Yes - in this study | Yes | 38.70 |
| **Malo et al., 2014 (118) Denmark, Spain.** | Point prevalence survey | Primary, community setting or ambulatory care | NS | NS | NS | n/a | Proxy | All penicillins<br>Drugs that make up 90% of antibiotic volume (DU90)<br>Cephalosporins<br>Macrolides, lincosamides, and streptogramins<br>Fluoroquinolones<br>Beta-lactamase sensitive penicillins<br>Penicillins plus beta-lactamase inhibitor<br>3rd and 4th generation cephalosporin<br>Broad/Narrow ratio<br>Seasonality of total antibiotic use<br>Seasonality of quinolone use | Pre-existing indicator - ESAC | Yes - in previous study | n/a | |
| **Manzano-Garcia et al., 2016 (107) Spain** | Retrospective observational study | Hospital, including private settings | NS | Treatment | Adults | Patients prescribed ertapenem | General | Is decision compliant with local, regional, or national guidelines? | Using treatment or diagnostic guidelines | NS | Yes | 17.30 |
| **March-Lopez et al., 2020 (109) Spain** | Feasibility study | Primary, community setting or ambulatory care | Various | Treatment | Adults and children | n/a | Both | Is decision compliant with local, regional, or national guidelines?<br>Has the indication being treated been considered?<br>Is the choice correct?<br>Is the duration correct?<br>Is the dose correct?<br>Is the timing of administration correct?<br>Rate of patients prescribed an antibiotic based on the indication<br>Patients prescribed the first line choice for the specified indication<br>Moxifloxacin<br>Male patients receiving 1st generation fluoroquinolones, nitrofurantoin or fosfomycin<br>Patients having appropriate diagnostic tests before prescription of antibiotic<br>Pregnant patients prescribed doxycycline | Literature review | Yes - in this study | No | |
| **March-Lopez et al., 2021 (108) Spain** | Feasibility study | Primary, community setting or ambulatory care | NS | treatment | NS | Patients receiving Outpatient Antimicrobial therapy (OPAT) | General | Has there been appropriate documentation in the clinical record?<br>Is the choice correct?<br>Is the dose correct?<br>Is the frequency correct?<br>Is the duration correct? | Literature review | Yes - in this study | Yes | 20.30 |

| | | | | | | | | | | | | |
|---|---|---|---|---|---|---|---|---|---|---|---|---|
| **Marek et al., 2020 (130) Canada** | A multicenter quasi-experimental study | Hospital, including private settings | NS | Treatment | Adults | Patients prescribed fluoroquinolones | General | Is decision compliant with local, regional, or national guidelines?<br>Have cultures been reviewed and treatment adapted accordingly?<br>Has the patient's risk of adverse events been considered? | NS | No | Yes | 17.00 |
| **Markham et al., 2025 (34) United States** | Cross-sectional study | Hospital, including private settings | NS | Treatment | Children | n/a | Proxy | Antibiotic diversity | Literature review | Yes - in this study | n/a | |
| **Marwat et al., 2024 (54) United Kingdom** | Audit and feedback intervention study | Hospital, including private settings | Prophylaxis of procedural infection | Prophylaxis | Adults | Oncology patients undergoing invasive procedures | General | Is decision compliant with local, regional, or national guidelines?<br>Has the indication being treated been considered?<br>Is the route correct?<br>Is the frequency correct?<br>Is the timing of administration correct?<br>Is the duration correct? | Literature review | No | Yes | 7.00 |
| **McCullough et al., 2017 (84) Australia** | Cross-sectional survey | Primary, community setting or ambulatory care | Acute Respiratory Infection | Treatment | NS | n/a | Proxy | Rate of patients prescribed an antibiotic based on the indication<br>Patients meeting required diagnostic criteria before prescription of antibiotic | Using treatment or diagnostic guidelines | NS | n/a | |
| **McMullan et al., 2020 (85) Australia** | NAPS data analysis | Hospital, including private settings | Neonatal sepsis | Treatment | Neonates <28 days | n/a | General | Is decision compliant with local, regional, or national guidelines?<br>Has whether the spectrum of the antibiotic is too broad or too narrow been considered?<br>Is the route correct?<br>Is the dose correct?<br>Is the frequency correct?<br>Is the duration correct?<br>Has the patient's allergy status been considered?<br>Have cultures been reviewed and treatment adapted accordingly?<br>Is decision in-line with expert opinion - e.g. Microbiologist, Infectious Diseases Consultant etc | Pre-existing Audit tool - NAPS | Yes - in previous study | Yes | 3.10 |
| **Mo et al., 2023 (86) Australia** | Longitudinal descriptive study | Hospital, including private settings | NS | Both | Adults | Patients prescribed metronidazole | General | Is decision compliant with local, regional, or national guidelines?<br>Has the activity of the antibiotic against the known or suspected organism been considered?<br>Has whether the spectrum of the antibiotic is too broad or too narrow been considered?<br>Is the route correct?<br>Is the dose correct?<br>Is the frequency correct?<br>Is the duration correct?<br>Has the patient's allergy status been considered?<br>Have cultures been reviewed and treatment adapted accordingly?<br>Is decision in-line with expert opinion - e.g. Microbiologist, Infectious Diseases Consultant etc<br>Is the choice correct? | Pre-existing Audit tool - NAPS | Yes - in previous study | Yes | 32.20 |
| **Mohamed et al., 2018 (55) United Kingdom** | Point Prevalence Survey | Hospital, including private settings | Prophylaxis of surgical infection | Prophylaxis | NS | Patients undergoing head and neck surgery | General | Is the duration correct? | Using treatment or diagnostic guidelines | NS | Yes | 88.00 |
| **Molero et al., 2020 (110) Spain** | Re-audit 6 years after intervention | Primary, community setting or | Acute pharyngitis | Treatment | NS | n/a | General | Have relevant diagnostic tests been carried out?<br>Is decision compliant with local, regional, or national guidelines? | Using treatment or | Yes - in previous study | Yes | 31.80 |

| | | | | | | | | | | | | |
|---|---|---|---|---|---|---|---|---|---|---|---|---|
| | | ambulatory care | | | | | | | diagnostic guidelines | | | |
| **Mull et al., 2020 (35) United States** | Comparison of manual vs electronic algorithm analysis of appropriateness | Hospital, including private settings | Prophylaxis for cardiac catheterisation | Prophylaxis | NS | Patients undergoing Cardiac catheterisation | General | Is the timing of administration correct? | Literature review | Yes - in this study | Yes | 2.20 |
| **Murphy et al., 2024 (36) United States** | Cross-sectional study | Hospital, including private settings | Dental infections and procedure | Both | NS | Patients undergoing dental procedures | General | Is decision compliant with local, regional, or national guidelines?<br>Is decision in-line with evidence-base or scientific information | Using treatment or diagnostic guidelines | NS | Yes | 35.70 |
| **Nagao et al., 2017 (142) Japan** | Retrospective cohort study | Hospital, including private settings | Staphylococcus aureus bacteraemia | Treatment | NS | n/a | General | Have diagnostic blood cultures been taken prior to antibiotic prescribing?<br>Is the timing of administration correct?<br>Is the duration correct?<br>Has the patient's allergy status been considered?<br>Has appropriate Therapeutic Drug Monitoring taken place?<br>Has the patient's severity criteria been considered? | Literature review | NS | No | |
| **Nakamura et al., 2024 (143) Japan** | Quasi-experimental study | Community Pharmacy | NS | Treatment | NS | Patients prescribed third generation cephalosporins | Proxy | 3rd and 4th generation cephalosporin<br>Days of therapy | NS | No | n/a | |
| **Nasso et al., 2022 (137) Italy** | Prospective audit | Hospital, including private settings | NS | treatment | Children | n/a | General | Is the choice correct?<br>Is decision compliant with local, regional, or national guidelines?<br>Is decision in-line with expert opinion - e.g. Microbiologist, Infectious Diseases Consultant etc<br>Has the indication being treated been considered? | Using treatment or diagnostic guidelines | No | Yes | 56.00 |
| **Nguyen et al., 2021 (163) Ireland** | Delphi consensus | Long term care or residential care setting | NS | Both | Adults | Care home residents | General | Is decision in-line with evidence-base or scientific information<br>Have the patient's co-morbidities been considered?<br>Has the indication being treated been considered? | Delphi consensus | No | No | |
| **Oltra Hostalet et al., 2018 (111) Spain** | Cross-sectional serial point-prevalence study | Hospital, including private settings | NS | Treatment | Adults | Patients in the emergency département | General | Is decision compliant with local, regional, or national guidelines?<br>Is decision in-line with expert opinion - e.g. Microbiologist, Infectious Diseases Consultant etc<br>Is the choice correct?<br>Has the indication being treated been considered?<br>Is the dose correct?<br>Is the route correct?<br>Is the duration correct?<br>Has the patient got clinical signs of an infection?<br>Has the activity of the antibiotic against the known or suspected organism been considered?<br>Have cultures been reviewed and treatment adapted accordingly? | NS | NS | Yes | 51.50 |
| **Oomen et al., 2022 (93) Netherlands** | Retrospective cross-sectional study | Hospital, including private settings | NS | Treatment | NS | Patients in the emergency department | General | Has there been appropriate documentation in the clinical record?<br>Is the choice correct?<br>Is the dose correct?<br>Is the route correct?<br>Is decision compliant with local, regional, or national guidelines?<br>Has the indication being treated been considered? | Literature review | Yes - in this study | Yes | 23.60 |
| **Osowicki et al., 2014 (87)** | Point Prevalence Survey | Hospital, including | NS | Both | Children | n/a | General | Is decision in-line with expert opinion - e.g. Microbiologist, Infectious Diseases Consultant etc | NS | NS | Yes | 18.00 |

| | | | | | | | | | | | | |
|---|---|---|---|---|---|---|---|---|---|---|---|---|
| **Australia** | | private settings | | | | | | Has the indication being treated been considered?<br>Have cultures been reviewed and treatment adapted accordingly?<br>Is decision compliant with local, regional, or national guidelines?<br>Are antibiotics indicated?<br>Has whether the spectrum of the antibiotic is too broad or too narrow been considered?<br>Is the dose correct?<br>Is the frequency correct?<br>Is the route correct?<br>Is the duration correct? | | | | |
| **Owens et al., 2024 (56) United Kingdom** | Retrospective study | Hospital, including private settings | NS | Treatment | NS | Patients discharged from medical specialities | General | Has the patient got a confirmed or suspected bacterial infection?<br>Has there been appropriate documentation in the clinical record?<br>Is the duration correct?<br>Is decision compliant with local, regional, or national guidelines? | Pre-existing Audit tool - UK National audit tool | Yes - in this study | Yes | 24.00 |
| **Palacios-Baena et al., 2020 (112) Spain** | Quasi-experimental intervention study protocol | Hospital, including private settings | NS | Treatment | NS | Patients on treatment with new antibiotics only | General | Is decision compliant with local, regional, or national guidelines?<br>Has the indication being treated been considered?<br>Is the route correct?<br>Is the dose correct?<br>Is the duration correct?<br>Has whether the spectrum of the antibiotic is too broad or too narrow been considered?<br>Is decision in-line with expert opinion - e.g. Microbiologist, Infectious Diseases Consultant etc | NS | NS | No | |
| **Park et al., 2024 (88) Australia** | Retrospective data analysis | Hospital, including private settings | Skin and soft tissue infections or post-surgery infections | Treatment | NS | n/a | General | Is decision compliant with local, regional, or national guidelines?<br>Has whether the spectrum of the antibiotic is too broad or too narrow been considered?<br>Is the route correct?<br>Is the dose correct?<br>Is the frequency correct?<br>Is the duration correct?<br>Has the patient's allergy status been considered?<br>Have cultures been reviewed and treatment adapted accordingly?<br>Is decision in-line with expert opinion - e.g. Microbiologist, Infectious Diseases Consultant etc<br>Is decision compliant with local, regional, or national guidelines?<br>Has the activity of the antibiotic against the known or suspected organism been considered?<br>Is decision in-line with expert opinion - e.g. Microbiologist, Infectious Diseases Consultant etc<br>Is the choice correct? | Pre-existing Audit tool - NAPS | Yes - in previous study | Yes | 22.9 |
| **Park et al., 2022 (151) Korea** | Point Prevalence Survey | In-patient and outpatient | NS | Both | NS | n/a | General | Is decision compliant with local, regional, or national guidelines?<br>Is decision compliant with local, regional, or national guidelines?<br>Is the choice correct?<br>Have cultures been reviewed and treatment adapted accordingly?<br>Has the patient got clinical signs of an infection?<br>Has whether there is any duplication, overlap or redundancy in the regimen been considered? | Expert group | Yes - in this study | Yes | 27.70 |

| | | | | | | | | | | | | |
|---|---|---|---|---|---|---|---|---|---|---|---|---|
| | | | | | | | | Is decision compliant with local, regional, or national guidelines?<br>Have the patient's previous culture results, including colonisation been considered?<br>Have the patient's risk factors for resistant bacteria been considered?<br>Has the patient's allergy status been considered?<br>Are antibiotics indicated?<br>Is decision compliant with local, regional, or national guidelines?<br>Has there been appropriate documentation in the clinical record? | | | | |
| **Pearson et al., 2015 (37) United States** | Comparative study | Primary, community setting or ambulatory care | Chlamydia trachomatis infection | Treatment | NS | Patients with chlamydia | General | Is decision compliant with local, regional, or national guidelines?<br>Is the choice correct? | Using treatment or diagnostic guidelines | Yes - in this study | Yes | 41.90 |
| **Pedeboscq et al., 2021 (68) France** | Quasi-experimental study | Hospital, including private settings | NS | NS | NS | Patients on fluoroquinol ones | General | Has the indication being treated been considered?<br>Is the choice correct?<br>Is the dose correct?<br>Is the duration correct?<br>Is the route correct?<br>Has whether combination or co-prescribing is needed been considered?<br>Is decision in-line with expert opinion - e.g. Microbiologist, Infectious Diseases Consultant etc | Literature review | Yes - in previous study | Yes | 81.00 |
| **Plate et al., 2020 (156) Switzerland** | Cross-sectional study | Primary, community setting or ambulatory care | Urinary Tract Infection | Treatment | Adults | n/a | Proxy | Rate of patients prescribed an antibiotic based on the indication<br>Patients prescribed the first line choice for the specified indication<br>Patients prescribed a fluoroquinolone for a specified indication | Pre-existing indicator - ESAC | Yes - in previous study | n/a | |
| **Poole et al., 2021 (119) United States and United Kingdom** | Report | Primary, community setting or ambulatory care | NS | NS | Children | n/a | Proxy | Rate of patients prescribed an antibiotic based on the indication<br>Broad/Narrow ratio<br>Amoxicillin<br>Amoxicillin and cefalexin<br>Azithromycin<br>Patients prescribed the first line choice for the specified indication<br>Mean duration | NS | NS | n/a | |
| **Poss-Doering et al., 2021 (147) Germany** | Cluster-randomized trial | Primary, community setting or ambulatory care | Various | Treatment | NS | n/a | Proxy | Rate of patients prescribed an antibiotic based on the indication | Pre-existing indicator - ESAC | Yes - in previous study | n/a | |
| **Quiros et al., 2022 (120) Argentina, Ecuador, Colombia, Uruguay, Brazil, Chile, Peru, Panama, and Bolivia** | Longitudinal descriptive study | Hospital, including private settings | NS | Both | Adults | Patients on Intensive care | General | Is the duration correct?<br>Is decision in-line with expert opinion - e.g. Microbiologist, Infectious Diseases Consultant etc<br>Has there been appropriate documentation in the clinical record?<br>Is decision compliant with local, regional, or national guidelines?<br>Is decision in-line with expert opinion - e.g. Microbiologist, Infectious Diseases Consultant etc<br>Has appropriate Therapeutic Drug Monitoring taken place?<br>Has whether there is any duplication, overlap or redundancy in the regimen been considered?<br>Has the treatment been de-escalated? | Literature review | Yes - in previous study | Yes | 71.70 |

| | | | | | | | | | | | | |
|---|---|---|---|---|---|---|---|---|---|---|---|---|
| | | | | | | | | Has intravenous treatment been reviewed and switched to oral treatment (IVOS) appropriately? | | | | |
| **Roberts et al., 2015 (57) United Kingdom** | Longitudinal descriptive study | Hospital, including private settings | NS | Treatment | Adults | Patients admitted to Acute Medical Admissions ward | General | Has there been appropriate documentation in the clinical record?<br>Is decision compliant with local, regional, or national guidelines? | NS | Yes - in this study | Yes | 21.10 |
| **Roberts et al., 2016 (38) United States** | Time analysis | Primary, community setting or ambulatory care | Respiratory Tract Infections | Treatment | Adults and children | n/a | Proxy | Patients having appropriate diagnostic tests before prescription of antibiotic<br>Rate of patients prescribed an antibiotic based on the indication | Literature review | Yes - in previous study | n/a | |
| **Robson et al., 2018 (58) United Kingdom** | Point Prevalence Survey | Hospital, including private settings | NS | Both | NS | Patients prescribed carbapenems or piperacillin/tazobactam | General | Has there been appropriate documentation in the clinical record?<br>Is decision compliant with local, regional, or national guidelines? | Literature review | Yes - in previous study | Yes | 21.00 |
| **Roche et al., 2016 (69) France** | Prospective audit | Primary, community setting or ambulatory care | NS | Both | Adults and children | Patients prescribed hospital only outpatient drugs | General | Is the dose correct?<br>Is the frequency correct?<br>Is the choice correct?<br>Is decision compliant with local, regional, or national guidelines? | Literature review | Yes - in previous study | Yes | 60.00 |
| **Rochefolle et al., 2017 (70) France** | Prospective observational study | Long term care or residential care setting | Urinary Tract Infection | Treatment | Adults | Patients in specialist neurology rehabilitation unit | General | Is decision compliant with local, regional, or national guidelines?<br>Is the dose correct?<br>Have cultures been reviewed and treatment adapted accordingly?<br>Is the duration correct?<br>Is decision in-line with evidence-base or scientific information | Expert group | Yes - in this study | Yes | 15.00 |
| **Roger et al., 2024 (71) France** | Retrospective multicenter audit | Hospital, including private settings | NS | Treatment | NS | n/a | General | Has there been appropriate documentation in the clinical record?<br>Has the treatment been reviewed or reassessed?<br>Is the choice correct?<br>Has the patient got a confirmed or suspected bacterial infection?<br>Is decision compliant with local, regional, or national guidelines?<br>Is the dose correct?<br>Has whether the IV or oral route is appropriate based on specified criteria been considered?<br>Have cultures been reviewed and treatment adapted accordingly? Have relevant diagnostic criteria been met? | Literature review | Yes - in this study | Yes | 79.00 |
| **Roger et al., 2022 (164) Ireland** | Intervention implementation study | Hospital, including private settings | NS | both | NS | n/a | General | Has the patient got a confirmed or suspected bacterial infection?<br>Is the choice correct?<br>Is decision compliant with local, regional, or national guidelines?<br>Has the treatment been reviewed or reassessed? | NS | No | Yes | 83.00 |
| **Roger et al., 2019 (72) France** | Prospective, multicenter study | Hospital, including private settings | NS | Treatment | NS | n/a | General | Has the patient got a confirmed or suspected bacterial infection?<br>Has whether there is any duplication, overlap or redundancy in the regimen been considered?<br>Have cultures been reviewed and treatment adapted accordingly? | Literature review | Yes - in previous study | Yes | 71.00 |

| | | | | | | | | | | | | |
|---|---|---|---|---|---|---|---|---|---|---|---|---|
| | | | | | | | | Is decision compliant with local, regional, or national guidelines?<br>Is the choice correct?<br>Is the dose correct? | | | | |
| **Run Sigurardottir et al., 2015 (121) Iceland and Denmark** | Cross-sectional study | Primary, community setting or ambulatory care | Upper respiratory infections | Treatment | NS | n/a | General | Have relevant diagnostic tests been carried out?<br>Have relevant diagnostic criteria been met? | Using treatment or diagnostic guidelines | Yes - in previous study | Yes | 11.50 |
| **Sathe et al., 2024 (39) United States** | Quality improvement project | Primary, community setting or ambulatory care | Upper respiratory infections | Treatment | NS | n/a | Proxy | Rate of patients prescribed an antibiotic based on the indication | Literature review | Yes - in this study | n/a | |
| **Saust et al., 2018 (172) Denmark** | Retrospective audit | Primary, community setting or ambulatory care | Acute Respiratory Infection | treatment | Adults and children | n/a | Proxy | Patients having appropriate diagnostic tests before prescription of antibiotic<br>Patients meeting required diagnostic criteria before prescription of antibiotic<br>Patients prescribed a fluoroquinolone for a specified indication<br>Patients prescribed an inappropriate antibiotic choice for the indication<br>Patients prescribed the first line choice for the specified indication<br>Patients without known penicillin allergy prescribed a pen-allergic treatment option | Literature review | NS | n/a | |
| **Schoffelen et al., 2021 (94) Netherlands** | Delphi process | Hospital, including private settings | NS | Treatment | Adults | Patients in the emergency department | General | Is decision compliant with local, regional, or national guidelines?<br>Is the dose correct?<br>Is the frequency correct?<br>Have the patient's previous culture results, including colonisation been considered?<br>Is the timing of administration correct?<br>Has there been appropriate documentation in the clinical record?<br>Has the patient been educated on how to take it?<br>Is the route correct?<br>Has the patient's previous antibiotic use been considered?<br>Has the patient's allergy status been considered?<br>Have the patient's contraindications been considered? | Delphi consensus | No | No | |
| **Sgro et al., 2021 (59) United Kingdom** | Audit | Hospital, including private settings | Suspected or confirmed Intra-abdominal infection | treatment | Adults | Patients prescribed intravenous metronidazole for suspected or confirmed intra-abdominal infection | General | Is the medicinal form correct?<br>Is decision compliant with local, regional, or national guidelines?<br>Has whether the IV or oral route is appropriate based on specified criteria been considered? | Using treatment or diagnostic guidelines | No | Yes | 81.40 |
| **Shah et al., 2023 (165) Ireland** | Point prevalence survey | Primary, community setting or ambulatory care | Respiratory Tract Infections | Treatment | Adults and children | n/a | Proxy | Rate of patients prescribed an antibiotic based on the indication<br>Patients prescribed the first line choice for the specified indication<br>Patients prescribed a fluoroquinolone for a specified indication | Pre-existing indicator - ESAC | Yes - in previous study | n/a | |

| | | | | | | | | | | | | |
|---|---|---|---|---|---|---|---|---|---|---|---|---|
| **Shin et al., 2024 (152) Korea** | Audit | Primary, community setting or ambulatory care | NS | Both | Adults | n/a | General | Has there been appropriate documentation in the clinical record?<br>Is decision compliant with local, regional, or national guidelines?<br>Has the indication being treated been considered?<br>Is decision in-line with expert opinion - e.g. Microbiologist, Infectious Diseases Consultant etc<br>Have cultures been reviewed and treatment adapted accordingly?<br>Has the patient's severity criteria been considered?<br>Is the duration correct?<br>Is the dose correct?<br>Is the frequency correct?<br>Has the patient's renal function been considered? | Literature review | Yes - in this study | Yes | 41.80 |
| **Sikkens et al., 2017 (95) Netherlands** | Pre and post intervention audit | Hospital, including private settings | NS | Both | NS | n/a | General | Has the indication being treated been considered?<br>Is the choice correct?<br>Is the dose correct?<br>Is the route correct?<br>Is the duration correct?<br>Is decision compliant with local, regional, or national guidelines?<br>Is decision in-line with expert opinion - e.g. Microbiologist, Infectious Diseases Consultant etc<br>Has the activity of the antibiotic against the known or suspected organism been considered?<br>Has whether there is any duplication, overlap or redundancy in the regimen been considered?<br>Has whether the spectrum of the antibiotic is too broad or too narrow been considered?<br>Has the patient's allergy status been considered?<br>Has intravenous treatment been reviewed and switched to oral treatment (IVOS) appropriately?<br>Have the patient's previous culture results, including colonisation been considered? | Literature review | Yes - in previous study | Yes | 33.90 |
| **Simon et al., 2021 (75) France** | Cross-sectional Observational Study | Long term care or residential care setting | NS | Treatment | Adults | Nursing home residents | Proxy | Patients prescribed the first line choice for the specified indication<br>Patients prescribed a fluoroquinolone within 6 months of another prescription for a fluoroquinolone<br>Seasonality of total antibiotic use<br>Seasonality of quinolone use<br>First line/Second line ratio<br>Patients prescribed an antibiotic that is not indicated<br>Duration greater than specified number of days<br>Patients prescribed antibiotic and non-steroidal anti-inflammatory drug (NSAID) on the same day<br>Patients co-prescribed a fluoroquinolone and steroid | Literature review | Yes - in previous study | n/a | |
| **Simon et al., 2024 (73) France** | Delphi consensus | Primary, community setting or ambulatory care | NS | Treatment | NS | n/a | Proxy | Patients prescribed the first line choice for the specified indication<br>Patients prescribed a fluoroquinolone within 6 months of another prescription for a fluoroquinolone<br>Seasonality of total antibiotic use<br>Seasonality of quinolone use<br>First line/Second line ratio<br>Patients prescribed an antibiotic that is not indicated<br>Duration greater than specified number of days<br>Patients prescribed antibiotic and non-steroidal anti-inflammatory drug (NSAID) on the same day<br>Patients co-prescribed a fluoroquinolone and steroid<br>Pristinamycin + macrolides | Delphi consensus | Yes - in this study | n/a | |

| | | | | | | | | | | | | |
|---|---|---|---|---|---|---|---|---|---|---|---|---|
| | | | | | | | | Amoxicillin/amoxicillin-clavulanate ratio | | | | |
| **Simon et al., 2023 (74) France** | Cross-sectional study | Dental | Dental infections | Treatment | Adults | n/a | Proxy | Amoxicillin/amoxicillin-clavulanate ratio<br>Duration greater than specified number of days<br>Patients prescribed an inappropriate antibiotic choice for the indication<br>Patients prescribed a rarely used antibiotic choice for the indication | Using treatment or diagnostic guidelines | Yes - in previous study | n/a | |
| **Singh et al., 2024 (89) Australia** | Retrospective audit | Hospital, including private settings | Neutropenic fever | NS | NS | n/a | General | Is decision compliant with local, regional, or national guidelines?<br>Has the activity of the antibiotic against the known or suspected organism been considered?<br>Has whether the spectrum of the antibiotic is too broad or too narrow been considered?<br>Is the route correct?<br>Is the dose correct?<br>Is the frequency correct?<br>Is the duration correct?<br>Has the patient's allergy status been considered?<br>Have cultures been reviewed and treatment adapted accordingly?<br>Is decision in-line with expert opinion - e.g. Microbiologist, Infectious Diseases Consultant etc<br>Is the choice correct? | Pre-existing Audit tool - NAPS | Yes - in previous study | Yes | 22.60 |
| **Skoog et al., 2016 (175) Sweden** | Point prevalence surveys | Hospital, including private settings | NS | Both | NS | n/a | General | Has there been appropriate documentation in the clinical record? | NS | Yes - in previous study | Yes | 16.80 |
| **Smith et al., 2014 (40) United States** | Intervention study | Hospital, including private settings | Pneumonia, cellulitis, urinary tract infection | Both | NS | n/a | Proxy | Days of therapy | Literature review | Yes - in previous study | n/a | |
| **Smith et al., 2018 (60) United Kingdom** | Point prevalence survey | Primary, community setting or ambulatory care | Various | Treatment | Adults and children | n/a | Proxy | Rate of patients prescribed an antibiotic based on the indication | Expert group | NS | n/a | |
| **Pernaselli's et al., 2022 (161) Greece** | Pre and post intervention audit | Hospital, including private settings | NS | Both | NS | n/a | General | Has there been appropriate documentation in the clinical record?<br>Is decision compliant with local, regional, or national guidelines? | NS | Yes - in previous study | Yes | 38.20 |
| **Spoorenberg et al., 2015 (96) Netherlands** | Cluster randomized controlled trial | Hospital, including private settings | Complicated urinary tract infection | Treatment | NS | n/a | General | Have diagnostic cultures from suspected site of infections been taken prior to antibiotic prescribing?<br>Is decision compliant with local, regional, or national guidelines?<br>Has intravenous treatment been reviewed and switched to oral treatment (IVOS) appropriately?<br>Have cultures been reviewed and treatment adapted accordingly?<br>Is the choice correct?<br>Is the duration correct?<br>Has the patient's gender been considered?<br>Has whether the patient has a catheter been considered?<br>Has the patient's renal function been considered? | Delphi consensus | Yes - in this study | No | |
| **Suda et al., 2022 (42) United States** | Cross-sectional study | Dental | Prophylaxis of infective endocarditis | Prophylaxis | Adults | Veterans | General | Are antibiotics indicated?<br>Is the patient at risk for an infection?<br>Is the choice correct?<br>Is decision compliant with local, regional, or national guidelines? | Using treatment or diagnostic guidelines | Yes - in this study | Yes | 85.00 |

| | | | | | | | | | | | | |
|---|---|---|---|---|---|---|---|---|---|---|---|---|
| **Suda et al., 2019 (41) United States** | Retrospective cohort study | dental | Prophylaxis of infective endocarditis | Prophylaxis | Adults | Veterans | General | Are antibiotics indicated?<br>Is the patient at risk for an infection?<br>Is decision compliant with local, regional, or national guidelines? | Using treatment or diagnostic guidelines | No | Yes | 80.90 |
| **Suzuki et al., 2021 (43) United States** | Retrospective cohort study | Hospital, including private settings | NS | Treatment | Adults | Veterans prescribed carbapenems | General | Is decision in-line with expert opinion - e.g. Microbiologist, Infectious Diseases Consultant etc<br>Has the indication being treated been considered?<br>Have cultures been reviewed and treatment adapted accordingly?<br>Have the patient’s previous culture results, including colonisation been considered?<br>Has the patient’s previous antibiotic use been considered?<br>Has the patient’s allergy status been considered?<br>Has whether there is any duplication, overlap or redundancy in the regimen been considered? | Expert group | Yes - in this study | Yes | 49.40 |
| **Teoh et al., 2024 (90) Australia** | Retrospective cohort study | Hospital, including private settings | Oral and dental infections | Both | NS | n/a | General | Is decision compliant with local, regional, or national guidelines?<br>Has the activity of the antibiotic against the known or suspected organism been considered?<br>Has whether the spectrum of the antibiotic is too broad or too narrow been considered?<br>Is the route correct?<br>Is the dose correct?<br>Is the frequency correct?<br>Is the duration correct?<br>Has the patient’s allergy status been considered?<br>Have cultures been reviewed and treatment adapted accordingly?<br>Is decision in-line with expert opinion - e.g. Microbiologist, Infectious Diseases Consultant etc<br>Is the choice correct? | Pre-existing Audit tool - NAPS | No | Yes | 34.70 |
| **Thaulow et al., 2019 (167) Norway** | Prospective observational study | Hospital, including private settings | NS | both | Children | n/a | General | Is decision compliant with local, regional, or national guidelines?<br>Is the choice correct?<br>Has the indication being treated been considered? | NS | No | Yes | 52.00 |
| **Thaulow et al., 2019 (168) Norway** | Prospective observational study | Hospital, including private settings | NS | both | Children | n/a | General | Is decision compliant with local, regional, or national guidelines?<br>Is the choice correct?<br>Is the dose correct?<br>Have cultures been reviewed and treatment adapted accordingly? | NS | NS | Yes | 28.00 |
| **Thiessen et al., 2017 (44) United States** | Retrospective cohort study | Primary, community setting or ambulatory care | Community Acquired Pneumonia and Hospital Acquired Pneumonia | Treatment | Adults | n/a | General | Is decision compliant with local, regional, or national guidelines?<br>Have the patient’s co-morbidities been considered?<br>Is the patient at risk for an infection?<br>Has the patient’s previous antibiotic use been considered?<br>Is the choice correct?<br>Is the dose correct?<br>Is the frequency correct?<br>Is the duration correct?<br>Is the route correct? | Using treatment or diagnostic guidelines | Yes - in this study | Yes | 65.50 |
| **Thilly et al., 2020 (76) France** | Cross-sectional observational study | Primary, community setting or ambulatory care | NS | Treatment | Children | n/a | Proxy | Seasonality of total antibiotic use<br>First line/Second line ratio<br>Patients prescribed antibiotic and non-steroidal anti-inflammatory drug (NSAID) on the same day<br>Patients co-prescribed a fluoroquinolone and steroid | Literature review | Yes - in this study | n/a | |
| **Thilly et al., 2020 (5)** | Cross-sectional study | Primary, community | NS | Treatment | NS | n/a | Proxy | Patients prescribed the first line choice for the specified indication | Literature review | Yes - in this study | n/a | |

| | | | | | | | | | | | | |
|---|---|---|---|---|---|---|---|---|---|---|---|---|
| **France** | | setting or ambulatory care | | | | | | Patients prescribed a fluoroquinolone within 6 months of another prescription for a fluoroquinolone<br>Seasonality of total antibiotic use<br>Seasonality of quinolone use<br>First line/Second line ratio<br>Patients prescribed an antibiotic that is not indicated<br>Duration greater than specified number of days<br>Patients prescribed antibiotic and non-steroidal anti-inflammatory drug (NSAID) on the same day<br>Patients co-prescribed a fluoroquinolone and steroid | | | | |
| **Timbrook et al., 2017 (45) United States** | Period prevalence survey | Hospital, including private settings | NS | Both | NS | Patients in the emergency department | General | Has the indication being treated been considered?<br>Has the treatment been effective?<br>Is the dose correct?<br>Has the patient's renal function been considered?<br>Is the frequency correct?<br>Has the practicality of the antibiotic been considered?<br>Have interactions with the patient's other medicines or feeds been considered?<br>Have the patient's contraindications been considered?<br>Has whether there is any duplication, overlap or redundancy in the regimen been considered?<br>Is the duration correct?<br>Is decision compliant with local, regional, or national guidelines? | Literature review | Yes - in previous study | yes | 39.00 |
| **Tribble et al., 2020 (46) United States** | Point prevalence survey | Hospital, including private settings | NS | Both | Children | n/a | General | Is decision in-line with expert opinion - e.g. Microbiologist, Infectious Diseases Consultant etc<br>Have cultures been reviewed and treatment adapted accordingly?<br>Has whether there is any duplication, overlap or redundancy in the regimen been considered?<br>Has whether the IV or oral route is appropriate based on specified criteria been considered?<br>Is the duration correct? | Pre-existing Audit tool - National audit tool | Yes - in previous study | Yes | 21.00 |
| **van Daalen et al., 2017 (97) Netherlands** | Multicentre stepped wedge cluster randomized trial, | Hospital, including private settings | NS | Both | Adults | Patients on intravenous antibiotics | General | Have diagnostic blood cultures been taken prior to antibiotic prescribing?<br>Have diagnostic cultures from suspected site of infections been taken prior to antibiotic prescribing?<br>Is decision compliant with local, regional, or national guidelines?<br>Has the patient's renal function been considered?<br>Is the dose correct?<br>Has there been appropriate documentation in the clinical record?<br>Has the treatment been reviewed or reassessed?<br>Have cultures been reviewed and treatment adapted accordingly?<br>Has intravenous treatment been reviewed and switched to oral treatment (IVOS) appropriately?<br>Is the frequency correct? | Literature review | Yes - in previous study | No | |
| **van den Bosch et al., 2016 (99) Netherlands** | Observational multicenter study | Hospital, including private settings | NS | Treatment | Adults | n/a | General | Is decision compliant with local, regional, or national guidelines?<br>Have diagnostic blood cultures been taken prior to antibiotic prescribing?<br>Have diagnostic cultures from suspected site of infections been taken prior to antibiotic prescribing?<br>Has there been appropriate documentation in the clinical record?<br>Has intravenous treatment been reviewed and switched to oral treatment (IVOS) appropriately? | Using treatment or diagnostic guidelines | Yes - in this study | No | |

| | | | | | | | | | | | |
|---|---|---|---|---|---|---|---|---|---|---|---|
| | | | | | | | | Have cultures been reviewed and treatment adapted accordingly? | | | |
| **van den Bosch et al., 2015 (122) Netherlands, Spain, Belgium, Scotland, Croatia, and Sweden** | RAND-modified Delphi process | Hospital, including private settings | NS | Treatment | Adults | n/a | General | Is decision compliant with local, regional, or national guidelines?<br>Have diagnostic blood cultures been taken prior to antibiotic prescribing?<br>Have diagnostic cultures from suspected site of infections been taken prior to antibiotic prescribing?<br>Have cultures been reviewed and treatment adapted accordingly?<br>Has the patient's renal function been considered?<br>Has intravenous treatment been reviewed and switched to oral treatment (IVOS) appropriately?<br>Has there been appropriate documentation in the clinical record?<br>Has appropriate Therapeutic Drug Monitoring taken place?<br>Has the treatment been stopped if there is no evidence of infection?<br>Is the duration correct? | Delphi consensus | Yes - in this study | No |
| **van den Bosch et al., 2014 (98) Netherlands** | A RAND-modified, five step Delphi procedure | Hospital, including private settings | Sepsis | treatment | Adults | n/a | General | Has whether the IV or oral route is appropriate based on specified criteria been considered?<br>Is the timing of administration correct?<br>Have diagnostic blood cultures been taken prior to antibiotic prescribing?<br>Have diagnostic cultures from suspected site of infections been taken prior to antibiotic prescribing?<br>Have cultures been reviewed and treatment adapted accordingly?<br>Is decision compliant with local, regional, or national guidelines?<br>Is the choice correct? | Delphi consensus | Yes - in this study | No |
| **van den Eijnde et al., 2024 (100) Netherlands** | Retrospective study | Primary, community setting or ambulatory care | Various | treatment | NS | n/a | Proxy | Amoxicillin-clavulanate<br>Macrolides<br>Fluoroquinolones<br>Amoxicillin/clavulanic acid + Macrolides + Quinolones<br>Rate of patients prescribed an antibiotic based on the indication<br>Patients prescribed the first line choice for the specified indication | Literature review | No | n/a |
| **van der Velden et al., 2016 (101) Netherlands** | Case study | Primary, community setting or ambulatory care | NS | Treatment | NS | n/a | Proxy | All penicillins<br>Cephalosporins<br>Macrolides, lincosamides, and streptogramins<br>Fluoroquinolones<br>Beta-lactamase sensitive penicillins<br>Penicillins plus beta-lactamase inhibitor<br>3rd and 4th generation cephalosporin<br>Broad/Narrow ratio<br>Seasonality of total antibiotic use<br>Seasonality of quinolone use | Pre-existing indicator - ESAC | Yes - in this study | n/a |
| **van der Velden et al., 2020 (102) Netherlands** | Consensus and pilot study | Primary, community setting or ambulatory care | various | Treatment | NS | n/a | Proxy | Penicillins plus beta-lactamase inhibitor<br>Macrolides<br>Fluoroquinolones<br>Amoxicillin/clavulanic acid + Macrolides + Quinolones<br>Rate of patients prescribed an antibiotic based on the indication<br>Patients prescribed the first line choice for the specified indication | Expert group | Yes - in this study | n/a |

| | | | | | | | | | | | | |
|---|---|---|---|---|---|---|---|---|---|---|---|---|
| **Vandael et al., 2020 (158) Belgium** | Point Prevalence Survey | Hospital, including private settings | NS | Treatment | NS | n/a | General | Is the choice correct?<br>Has there been appropriate documentation in the clinical record?<br>Is the route correct?<br>Is decision compliant with local, regional, or national guidelines?<br>Has the indication being treated been considered? | Pre-existing Audit tool - global and ECDC audit tool | yes - in previous study | yes | 23.40 |
| **Varol et al., 2020 (170) Türkiye** | Observational cross-sectional study | Primary, community setting or ambulatory care | Acute exacerbations of Chronic Obstructive Pulmonary Disease | Treatment | Adults | n/a | General | Is decision compliant with local, regional, or national guidelines??<br>Have relevant diagnostic criteria been met? | Using treatment or diagnostic guidelines | NS | yes | 36.00 |
| **Vazouras et al., 2023 (162) Greece** | Retrospective observational cohort study | Hospital, including private settings | Urinary tract infection | Both | Children | n/a | General | Is the choice correct?<br>Is the duration correct?<br>Has the activity of the antibiotic against the known or suspected organism been considered?<br>Has an inappropriate combination been prescribed?<br>Has intravenous treatment been reviewed and switched to oral treatment (IVOS) appropriately?<br>Is the dose correct?<br>Has the patient's renal function been considered?<br>Have relevant diagnostic criteria been met?<br>Has the treatment been de-escalated?<br>Are antibiotics indicated? | Literature review | No | Yes | 18.50 |
| **Velasco-Arnaiz et al., 2020 (113) Spain** | Pre-post study | Hospital, including private settings | NS | Both | Children | n/a | General | Has the indication being treated been considered?<br>Has the activity of the antibiotic against the known or suspected organism been considered?<br>Is decision compliant with local, regional, or national guidelines??<br>Has the patient's allergy status been considered?<br>Have the patient's co-morbidities been considered?<br>Is the route correct?<br>Is the dose correct?<br>Is the frequency correct?<br>Is the duration correct? | Literature review | No | Yes | 14.40 |
| **Vellinga et al., 2023 (123) Belgium, Croatia, Denmark, Georgia, Germany, Greece, Ireland, Moldova, the Netherlands, Poland, Romania, Spain, and the United Kingdom** | Point Prevalence Audit Surveys | Primary, community setting or ambulatory care | Respiratory Tract Infections | Both | Adults and children | n/a | Proxy | Rate of patients prescribed an antibiotic based on the indication | Pre-existing indicator - ESAC | Yes - in previous study | n/a | |
| **Wild et al., 2022 (148) Germany** | Quasi-experimental before–after study | Hospital, including private settings | NS | Both | NS | n/a | Proxy | Intravenous antibiotics | NS | no | n/a | |
| **Willems et al., 2023 (159) Belgium** | Prospective audit and feedback | Hospital, including | NS | Treatment | Adults | Patients on piperacillin-tazobactam | General | Is the choice correct?<br>Has the patient got a confirmed or suspected bacterial infection? | Literature review | Yes - in previous study | Yes | 39.70 |

| | | | | | | | | | | | | |
|---|---|---|---|---|---|---|---|---|---|---|---|---|
| | | private settings | | | | , meropenem and vancomycin i.v. | | Is decision compliant with local, regional, or national guidelines?<br>Has whether the spectrum of the antibiotic is too broad or too narrow been considered?<br>Is the dose correct?<br>Is the frequency correct?<br>Is the duration correct?<br>Has there been appropriate documentation in the clinical record? | | | | |
| **Willemsen et al., 2018 (103) Netherlands** | Audits and risk assessment | Hospital, several nursing homes, and a rehabilitation clinic | NS | Treatment | NS | n/a | General | Has the indication being treated been considered?<br>Is the choice correct?<br>Is decision compliant with local, regional, or national guidelines?<br>Are antibiotics indicated? | NS | Yes - in this study | no | |
| **Williams et al., 2018 (61) United Kingdom** | Observational study | Primary, community setting or ambulatory care | Acute upper respiratory tract infections (URTIs) | Treatment | Children | n/a | Proxy | Rate of patients prescribed an antibiotic based on the indication<br>Patients prescribed the first line choice for the specified indication<br>Patients prescribed a fluoroquinolone for a specified indication | Pre-existing indicator - ESAC | Yes - in previous study | n/a | |
| **Wu et al., 2020 (131) Canada** | Delphi consensus | Primary, community setting or ambulatory care | Various | Treatment | Adults and children | n/a | Proxy | Rate of patients prescribed an antibiotic based on the indication | Delphi consensus | No | n/a | |
| **Yesudian et al., 2015 (62) United Kingdom** | Multi cycle audit | Hospital, including private settings | Dental infections | Both | Children | Patients in the dental department | General | Is decision compliant with local, regional, or national guidelines?<br>Has the indication being treated been considered?<br>Is the dose correct?<br>Is the frequency correct?<br>Is the duration correct?<br>Is the medicinal form correct? | Using treatment or diagnostic guidelines | no | yes | 72.00 |

### S4 – Risk of Bias Assessment (Using Mixed Methods Appraisal Tool(13))

| | 1. QUALITATIVE STUDIES | | | | | 2. RANDOMIZED CONTROLLED TRIALS | | | | | 3. NON-RANDOMIZED STUDIES | | | | | 4. QUANTITATIVE DESCRIPTIVE STUDIES | | | | | 5. MIXED METHODS STUDIES | | | | | Percentage of relevant criteria met |
|---|---|---|---|---|---|---|---|---|---|---|---|---|---|---|---|---|---|---|---|---|---|---|---|---|---|---|
| Study (Author, Year) | 1.1 | 1.2 | 1.3 | 1.4 | 1.5 | 2.1 | 2.2 | 2.3 | 2.4 | 2.5 | 3.1 | 3.2 | 3.3 | 3.4 | 3.5 | 4.1 | 4.2 | 4.3 | 4.4 | 4.5 | 5.1 | 5.2 | 5.3 | 5.4 | 5.5 | |
| Abbas et al., 2022 (77) | | | | | | | | | | | Y | Y | Y | Y | Y | | | | | | | | | | | 100 |
| Aghdassi et al., 2019 (144) | | | | | | | | | | | | | | | | Y | Y | Y | Y | Y | | | | | | 100 |
| Alba Fernandez et al., 2022 (104) | | | | | | | | | | | | | | | | CT | Y | Y | CT | N | | | | | | 40 |
| Arcenillas et al., 2018 (6) | | | | | | | | | | | | | | | | Y | Y | Y | Y | Y | | | | | | 100 |
| Arnoldo et al., 2019 (132) | | | | | | | | | | | | | | | | Y | Y | Y | Y | Y | | | | | | 100 |
| Asquier-Khati et al., 2023 (63) | Y | Y | Y | Y | Y | | | | | | | | | | | | | | | | | | | | | 100 |
| Baclet et al., 2022 (64) | Y | N | Y | Y | Y | | | | | | | | | | | | | | | | | | | | | 80 |
| Baclet et al., 2024 (65) | Y | Y | Y | Y | Y | | | | | | | | | | | | | | | | | | | | | 100 |
| Barnett, et al. 2020 (14) | | | | | | | | | | | | | | | | Y | Y | Y | Y | Y | | | | | | 100 |
| Barrie et al., 2018 (47) | | | | | | | | | | | | | | | | Y | Y | Y | Y | Y | | | | | | 100 |
| Barstow et al., 2020 (15) | | | | | | | | | | | | | | | | Y | CT | Y | CT | Y | | | | | | 60 |
| Berrevoets et al., 2017 (91) | | | | | | | | | | | | | | | | Y | Y | Y | Y | Y | | | | | | 100 |
| Bjerrum et al., 2022 (114) | | | | | | | | | | | | | | | | | | | | | | | | | | n/a |
| Black et al., 2018(124) | | | | | | | | | | | | | | | | Y | Y | Y | Y | Y | | | | | | 100 |
| Bohan et al., 2019 (16) | | | | | | | | | | | | | | | | Y | Y | Y | Y | Y | | | | | | 100 |
| Burns et al., 2020 (17) | | | | | | | | | | | | | | | | Y | Y | Y | Y | Y | | | | | | 100 |
| Cameron et al., 2015 (48) | | | | | | | | | | | | | | | | | | | | | Y | Y | Y | Y | Y | 100 |
| Canoui et al., 2018 (49) | | | | | | | | | | | | | | | | Y | Y | Y | Y | Y | | | | | | 100 |
| Cao et al., 2016 (18) | | | | | | | | | | | CT | Y | CT | Y | CT | | | | | | | | | | | 40 |
| Castel et al., 2016 (66) | | | | | | | | | | | | | | | | Y | Y | Y | Y | CT | | | | | | 80 |
| Catho et al., 2018 (153) | | | | | | | | | | | | | | | | | | | | | | | | | | n/a |
| Cattani et al., 2020 (133) | | | | | | | | | | | Y | Y | Y | Y | N | | | | | | | | | | | 80 |
| Chen et al., 2023 (125) | Y | N | Y | Y | Y | | | | | | | | | | | | | | | | | | | | | 80 |
| Choi et al., 2021 (19) | | | | | | | | | | | Y | Y | Y | N | Y | | | | | | | | | | | 80 |
| Chopra et al., 2014 (50) | | | | | | | | | | | | | | | | CT | CT | Y | CT | CT | | | | | | 20 |
| Chorafa et al., 2021 (160) | | | | | | | | | | | | | | | | Y | Y | Y | Y | Y | | | | | | 100 |
| Clegg et al., 2021 (20) | | | | | | Y | Y | Y | Y | Y | | | | | | | | | | | | | | | | 100 |
| Colombo et al., 2025 (21) | | | | | | | | | | | | | | | | Y | Y | Y | Y | Y | | | | | | 100 |
| Cona et al., 2021 (134) | | | | | | | | | | | | | | | | Y | Y | Y | Y | Y | | | | | | 100 |

| | | | | | | | | | | | | | | | | | | | | | | | | | | |
|---|---|---|---|---|---|---|---|---|---|---|---|---|---|---|---|---|---|---|---|---|---|---|---|---|---|---|
| Cotta et al., 2014 (78) | | | | | | | | | | | | | | | | Y | CT | Y | Y | CT | | | | | | 60 |
| Datta et al., 2019 (22) | | | | | | | | | | | | | | | | Y | N | Y | Y | Y | | | | | | 80 |
| Degnan et al., 2022 (23) | | | | | | | | | | | Y | Y | Y | Y | Y | | | | | | | | | | | 100 |
| Denny et al., 2019 (79) | | | | | | | | | | | | | | | | Y | Y | Y | Y | Y | | | | | | 100 |
| Dentan et al., 2017 (67) | | | | | | | | | | | | | | | | Y | Y | Y | Y | Y | | | | | | 100 |
| DePestel et al. 2014 (24) | | | | | | | | | | | | | | | | Y | Y | Y | Y | Y | | | | | | 100 |
| Devchand et al., 2019 (80) | | | | | | | | | | | | | | | | Y | Y | Y | Y | Y | | | | | | 100 |
| Doyle et al., 2021 (126) | | | | | | | | | | | Y | Y | CT | CT | Y | | | | | | | | | | | 60 |
| Dresser et al., 2018 (127) | Y | Y | Y | Y | Y | | | | | | | | | | | | | | | | | | | | | 100 |
| Dulku et al., 2022 (128) | | | | | | | | | | | | | | | | Y | Y | Y | Y | Y | | | | | | 100 |
| Durkin et al., 2018 (25) | | | | | | | | | | | Y | Y | Y | N | Y | | | | | | | | | | | 80 |
| Durkin et al., 2018 (26) | | | | | | | | | | | Y | Y | Y | CT | Y | | | | | | | | | | | 80 |
| Erturk Sengal et al., 2019 (169) | | | | | | | | | | | Y | Y | CT | N | Y | | | | | | | | | | | 60 |
| Eure et al., 2017 (27) | | | | | | | | | | | | | | | | Y | N | Y | Y | Y | | | | | | 80 |
| Fleming et al., 2015 (115) | Y | Y | Y | Y | Y | | | | | | | | | | | | | | | | | | | | | 100 |
| Fleming-Dutra et al., 2016 (28) | | | | | | | | | | | | | | | | Y | Y | Y | Y | Y | | | | | | 100 |
| Forst et al., 2024 (145) | | | | | | | | | | | | | | | | Y | Y | Y | Y | Y | | | | | | 100 |
| Franchi et al., 2021 (135) | | | | | | | | | | | | | | | | Y | Y | Y | Y | Y | | | | | | 100 |
| Friedman et al., 2020 (81) | | | | | | | | | | | | | | | | Y | Y | Y | Y | Y | | | | | | 100 |
| Garcia-Sangenis et al., 2024 (116) | | | | | | | | | | | | | | | | Y | Y | Y | Y | Y | | | | | | 100 |
| Gardiner et al., 2020 (174) | | | | | | | | | | | | | | | | Y | Y | Y | Y | Y | | | | | | 100 |
| Garlasco et al., 2024 (136) | | | | | | | | | | | | | | | | Y | Y | Y | Y | Y | | | | | | 100 |
| Gharbi et al., 2016 (51) | | | | | | | | | | | | | | | | Y | Y | Y | Y | Y | | | | | | 100 |
| Gimenez-Perez et al., 2024 (105) | | | | | | | | | | | Y | Y | Y | N | Y | | | | | | | | | | | 80 |
| Glinz et al., 2017 (154) | | | | | | | | | | | | | | | | Y | N | Y | Y | Y | | | | | | 80 |
| Gurtler et al., 2019 (155) | | | | | | | | | | | | | | | | Y | Y | Y | Y | Y | | | | | | 100 |
| Gutierrez-Urbon et al., 2024 (106) | | | | | | | | | | | | | | | | Y | Y | Y | Y | Y | | | | | | 100 |
| Guzik et al., 2019 (29) | | | | | | | | | | | | | | | | Y | Y | Y | Y | Y | | | | | | 100 |
| Hahn et al., 2019 (30) | | | | | | | | | | | Y | Y | Y | N | Y | | | | | | | | | | | 80 |
| Hearsey et al., 2022 (52) | | | | | | | | | | | | | | | | Y | N | Y | Y | CT | | | | | | 60 |
| Higgins et al., 2023 (53) | | | | | | | | | | | | | | | | Y | Y | Y | Y | Y | | | | | | 100 |
| Hogli et al., 2016 (166) | | | | | | | | | | | Y | Y | Y | Y | Y | | | | | | | | | | | 100 |
| Hood et al., 2019 (9) | Y | Y | Y | Y | Y | | | | | | | | | | | | | | | | | | | | | 100 |

| | | | | | | | | | | | | | | | | | | | | | | | | | | |
|---|---|---|---|---|---|---|---|---|---|---|---|---|---|---|---|---|---|---|---|---|---|---|---|---|---|---|
| Hussein et al., 2017 (146) | Y | Y | Y | Y | Y | | | | | | | | | | | | | | | | | | | | | 100 |
| Ierano et al., 2020 (82) | | | | | | | | | | | | | | | | Y | Y | Y | Y | Y | | | | | | 100 |
| James et al., 2015 (83) | | | | | | | | | | | | | | | | Y | CT | Y | CT | CT | | | | | | 40 |
| Jung et al., 2023 (149) | | | | | | | | | | | | | | | | Y | Y | Y | Y | Y | | | | | | 100 |
| Kallen et al., 2018 (92) | | | | | | | | | | | Y | Y | Y | N | Y | | | | | | | | | | | 80 |
| Kern et al., 2024 (171) | | | | | | | | | | | | | | | | Y | CT | N | CT | Y | | | | | | 40 |
| Khaw et al., 2018 (31) | | | | | | | | | | | Y | Y | Y | N | Y | | | | | | | | | | | 80 |
| Kim et al., 2021 (150) | Y | Y | Y | Y | Y | | | | | | | | | | | | | | | | | | | | | 100 |
| Kinoshita et al., 2020 (138) | | | | | | | | | | | Y | Y | Y | Y | Y | | | | | | | | | | | 100 |
| Komagamine et al., 2019 (139) | | | | | | | | | | | | | | | | Y | Y | Y | Y | Y | | | | | | 100 |
| Komagamine et al., 2019 (140) | | | | | | | | | | | | | | | | Y | Y | Y | Y | Y | | | | | | 100 |
| Kusama, 2024 (141) | | | | | | | | | | | | | | | | | | | | | | | | | | n/a |
| Lass et al., 2020 (173) | | | | | | | | | | | | | | | | Y | Y | Y | Y | Y | | | | | | 100 |
| Legros et al., 2024 (157) | Y | Y | Y | Y | Y | | | | | | | | | | | | | | | | | | | | | 100 |
| Leslie et al., 2023 (129) | | | | | | | | | | | | | | | | Y | Y | Y | Y | Y | | | | | | 100 |
| Levine et al., 2016 (32) | | | | | | | | | | | | | | | | Y | Y | Y | Y | Y | | | | | | 100 |
| Liu et al., 2019 (33) | | | | | | | | | | | | | | | | | | | | | | | | | | n/a |
| Lopez-Campos et al., 2015 (117) | | | | | | | | | | | Y | Y | Y | Y | Y | | | | | | | | | | | 100 |
| Malo et al., 2014 (118) | | | | | | | | | | | | | | | | Y | Y | Y | Y | Y | | | | | | 100 |
| Manzano-Garcia et al., 2016 (107) | | | | | | | | | | | | | | | | Y | Y | Y | Y | Y | | | | | | 100 |
| March-Lopez et al., 2020 (109) | | | | | | | | | | | | | | | | Y | Y | Y | Y | Y | | | | | | 100 |
| March-Lopez et al., 2021 (108) | | | | | | | | | | | | | | | | Y | Y | Y | Y | Y | | | | | | 100 |
| Marek et al., 2020 (130) | | | | | | | | | | | Y | Y | Y | N | Y | | | | | | | | | | | 80 |
| Markham et al., 2025 (34) | | | | | | | | | | | | | | | | Y | Y | Y | Y | Y | | | | | | 100 |
| Marwat et al., 2024 (54) | | | | | | | | | | | Y | Y | Y | N | Y | | | | | | | | | | | 80 |
| McCullough et al., 2017 (84) | | | | | | | | | | | | | | | | Y | Y | Y | Y | Y | | | | | | 100 |
| McMullan et al., 2020 (85) | | | | | | | | | | | | | | | | Y | Y | Y | Y | Y | | | | | | 100 |
| Mo et al., 2023 (86) | | | | | | | | | | | | | | | | Y | N | Y | Y | Y | | | | | | 80 |
| Mohamed et al., 2018 (55) | | | | | | | | | | | Y | Y | N | CT | CT | | | | | | | | | | | 40 |
| Molero et al., 2020 (110) | | | | | | | | | | | Y | Y | Y | Y | Y | | | | | | | | | | | 100 |
| Mull et al., 2020 (35) | | | | | | | | | | | Y | Y | N | Y | Y | | | | | | | | | | | 80 |
| Murphy et al., 2024 (36) | | | | | | | | | | | | | | | | Y | Y | Y | Y | Y | | | | | | 100 |
| Nagao et al., 2017 (142) | | | | | | | | | | | | | | | | Y | Y | Y | Y | Y | | | | | | 100 |

| | | | | | | | | | | | | | | | | | | | | | | | | | | |
|---|---|---|---|---|---|---|---|---|---|---|---|---|---|---|---|---|---|---|---|---|---|---|---|---|---|---|
| Nakamura et al., 2024 (143) | | | | | | | | | | | CT | Y | CT | N | CT | | | | | | | | | | | 20 |
| Nasso et al., 2022 (137) | | | | | | | | | | | | | | | | Y | Y | Y | Y | Y | | | | | | 100 |
| Nguyen et al., 2021 (163) | Y | Y | Y | Y | Y | | | | | | | | | | | | | | | | | | | | | 100 |
| Oltra Hostalet et al., 2018 (111) | | | | | | | | | | | | | | | | Y | Y | Y | Y | Y | | | | | | 100 |
| Oomen et al., 2022 (93) | | | | | | | | | | | | | | | | Y | Y | Y | Y | Y | | | | | | 100 |
| Osowicki et al., 2014 (87) | | | | | | | | | | | | | | | | Y | Y | Y | Y | Y | | | | | | 100 |
| Owens et al., 2024 (56) | | | | | | | | | | | | | | | | N | N | CT | Y | N | | | | | | 20 |
| Palacios-Baena et al., 2020 (112) | | | | | | | | | | | | | | | | Y | Y | Y | Y | Y | | | | | | 100 |
| Park et al., 2024 (88) | | | | | | | | | | | | | | | | Y | Y | Y | Y | Y | | | | | | 100 |
| Park et al., 2022 (151) | | | | | | | | | | | | | | | | Y | Y | Y | Y | Y | | | | | | 100 |
| Pearson et al., 2015 (37) | | | | | | | | | | | | | | | | Y | Y | Y | Y | Y | | | | | | 100 |
| Pedeboscq et al., 2021 (68) | | | | | | | | | | | | | | | | Y | Y | Y | Y | Y | | | | | | 100 |
| Plate et al., 2020 (156) | | | | | | | | | | | | | | | | Y | Y | Y | CT | Y | | | | | | 80 |
| Poole et al., 2021 (119) | | | | | | | | | | | | | | | | | | | | | | | | | | n/a |
| Poss-Doering et al., 2021 (147) | | | | | | Y | Y | Y | N | CT | | | | | | | | | | | | | | | | 60 |
| Quiros et al., 2022 (120) | | | | | | | | | | | Y | Y | Y | Y | Y | | | | | | | | | | | 100 |
| Roberts et al., 2015 (57) | | | | | | | | | | | Y | N | Y | N | Y | | | | | | | | | | | 60 |
| Roberts et al., 2016 (38) | | | | | | | | | | | | | | | | Y | Y | Y | CT | Y | | | | | | 80 |
| Robson et al., 2018 (58) | Y | Y | Y | Y | Y | | | | | | | | | | | | | | | | | | | | | 100 |
| Roche et al., 2016 (69) | | | | | | | | | | | | | | | | Y | Y | Y | Y | Y | | | | | | 100 |
| Rochefolle et al., 2017 (70) | | | | | | | | | | | | | | | | Y | Y | Y | Y | Y | | | | | | 100 |
| Roger et al., 2024 (71) | | | | | | | | | | | Y | Y | Y | Y | Y | | | | | | | | | | | 100 |
| Roger et al., 2022 (164) | | | | | | | | | | | Y | Y | Y | Y | Y | | | | | | | | | | | 100 |
| Roger et al., 2019 (72) | | | | | | | | | | | | | | | | Y | Y | Y | Y | Y | | | | | | 100 |
| Run Sigurardottir et al., 2015 (121) | | | | | | | | | | | | | | | | Y | Y | Y | Y | Y | | | | | | 100 |
| Sathe et al., 2024 (39) | | | | | | | | | | | Y | Y | CT | Y | CT | | | | | | | | | | | 60 |
| Saust et al., 2018 (172) | | | | | | | | | | | | | | | | Y | Y | Y | CT | Y | | | | | | 80 |
| Schoffelen et al., 2021 (94) | | | | | | | | | | | | | | | | | | | | | Y | Y | Y | CT | Y | 80 |
| Sgro et al., 2021 (59) | | | | | | | | | | | Y | Y | Y | CT | Y | | | | | | | | | | | 80 |
| Shah et al., 2023 (165) | | | | | | | | | | | | | | | | Y | Y | Y | Y | Y | | | | | | 100 |
| Shin et al., 2024 (152) | | | | | | | | | | | | | | | | Y | Y | Y | Y | Y | | | | | | 100 |
| Sikkens et al., 2017 (95) | | | | | | | | | | | | | | | | | | | | | Y | Y | Y | Y | Y | 100 |
| Simon et al., 2021 (75) | | | | | | | | | | | | | | | | Y | Y | Y | Y | Y | | | | | | 100 |

| | | | | | | | | | | | | | | | | | | | | | | | | | | |
|---|---|---|---|---|---|---|---|---|---|---|---|---|---|---|---|---|---|---|---|---|---|---|---|---|---|---|
| Simon et al., 2024 (73) | | | | | | | | | | | | | | | | | | | | | Y | Y | Y | Y | Y | 100 |
| Simon et al., 2023 (74) | | | | | | | | | | | | | | | | Y | Y | Y | Y | Y | | | | | | 100 |
| Singh et al., 2024 (89) | | | | | | | | | | | | | | | | Y | Y | Y | Y | CT | | | | | | 80 |
| Skoog et al., 2016 (175) | | | | | | | | | | | | | | | | Y | Y | Y | CT | N | | | | | | 60 |
| Smith et al., 2014 (40) | | | | | | | | | | | | | | | | Y | Y | Y | Y | Y | | | | | | 100 |
| Smith et al., 2018 (60) | | | | | | | | | | | | | | | | Y | Y | Y | CT | Y | | | | | | 80 |
| Spernovasilis et al., 2022 (161) | | | | | | | | | | | Y | Y | CT | CT | Y | | | | | | | | | | | 60 |
| Spoorenberg et al., 2015 (96) | | | | | | N | Y | Y | CT | Y | | | | | | | | | | | | | | | | 60 |
| Suda et al., 2022 (42) | | | | | | | | | | | | | | | | Y | Y | Y | Y | Y | | | | | | 100 |
| Suda et al., 2019 (41) | | | | | | | | | | | | | | | | Y | Y | Y | Y | Y | | | | | | 100 |
| Suzuki et al., 2021 (43) | | | | | | | | | | | | | | | | Y | Y | Y | Y | Y | | | | | | 100 |
| Teoh et al., 2024 (90) | | | | | | | | | | | | | | | | Y | Y | Y | CT | Y | | | | | | 80 |
| Thaulow et al., 2019 (167) | | | | | | | | | | | | | | | | Y | Y | Y | CT | Y | | | | | | 80 |
| Thaulow et al., 2019 (168) | | | | | | | | | | | | | | | | Y | CT | Y | CT | Y | | | | | | 60 |
| Thiessen et al., 2017 (44) | | | | | | | | | | | | | | | | Y | Y | Y | Y | Y | | | | | | 100 |
| Thilly et al., 2020 (76) | | | | | | | | | | | | | | | | Y | Y | Y | Y | Y | | | | | | 100 |
| Thilly et al., 2020 (5) | | | | | | | | | | | | | | | | Y | Y | Y | Y | Y | | | | | | 100 |
| Timbrook et al., 2017 (45) | | | | | | | | | | | | | | | | N | Y | Y | Y | Y | | | | | | 80 |
| Tribble et al., 2020 (46) | | | | | | | | | | | | | | | | Y | Y | Y | Y | Y | | | | | | 100 |
| van Daalen et al., 2017 (97) | | | | | | Y | Y | Y | N | Y | | | | | | | | | | | | | | | | 80 |
| van den Bosch et al., 2016 (99) | | | | | | | | | | | | | | | | | | | | | Y | Y | Y | Y | Y | 100 |
| van den Bosch et al., 2015 (122) | Y | Y | Y | Y | Y | | | | | | | | | | | | | | | | | | | | | 100 |
| van den Bosch et al., 2014 (98) | | | | | | | | | | | | | | | | | | | | | Y | Y | Y | Y | Y | 100 |
| van den Eijnde et al., 2024 (100) | | | | | | | | | | | | | | | | Y | Y | Y | Y | Y | | | | | | 100 |
| van der Velden et al., 2016 (101) | | | | | | | | | | | | | | | | Y | Y | Y | Y | Y | | | | | | 100 |
| van der Velden et al., 2020 (102) | | | | | | | | | | | | | | | | | | | | | Y | Y | Y | Y | CT | 80 |
| Vandael et al., 2020 (158) | | | | | | | | | | | | | | | | Y | Y | Y | Y | Y | | | | | | 100 |
| Varol et al., 2020 (170) | | | | | | | | | | | | | | | | Y | CT | Y | CT | Y | | | | | | 60 |
| Vazouras et al., 2023 (162) | | | | | | | | | | | | | | | | Y | N | Y | Y | CT | | | | | | 60 |
| Velasco-Arnaiz et al., 2020 (113) | | | | | | | | | | | | | | | | Y | CT | Y | Y | CT | | | | | | 60 |
| Vellinga et al., 2023 (123) | | | | | | | | | | | | | | | | Y | CT | Y | CT | Y | | | | | | 60 |
| Wild et al., 2022 (148) | | | | | | | | | | | Y | Y | Y | CT | Y | | | | | | | | | | | 80 |
| Willems et al., 2023 (159) | | | | | | | | | | | | | | | | Y | Y | Y | Y | Y | | | | | | 100 |

| | | | | | | | | | | | | | | | | | | | | | | | | | | |
|---|---|---|---|---|---|---|---|---|---|---|---|---|---|---|---|---|---|---|---|---|---|---|---|---|---|---|
| Willemsen et al., 2018 (103) | | | | | | | | | | | Y | Y | CT | CT | Y | | | | | | | | | | | 60 |
| Williams et al., 2018 (61) | | | | | | | | | | | | | | | | Y | Y | Y | Y | Y | | | | | | 100 |
| Wu et al., 2020 (131) | | | | | | | | | | | | | | | | | | | | | Y | Y | Y | Y | Y | 100 |
| Yesudian et al., 2015 (62) | Y | N | Y | N | Y | | | | | | | | | | | | | | | | | | | | | 60 |

Y= yes; N = No; CT = Can’t tell.

1.1 Is the qualitative approach appropriate to answer the research question?

1.2 Are the qualitative data collection methods adequate to address the research question?

1.3 Are the findings adequately derived from the data?

1.4 Is the interpretation of results sufficiently substantiated by data?

1.5 Is there coherence between qualitative data sources, collection, analysis and interpretation?

2.1 Is randomization appropriately performed?

2.2 Are the groups comparable at baseline?

2.3 Are there complete outcome data?

2.4 Are outcome assessors blinded to the intervention provided?

2.5 Did the participants adhere to the assigned intervention?

3.1 Are the participants representative of the target population?

3.2 Are measurements appropriate regarding both the outcome and intervention (or exposure)?

3.3 Are there complete outcome data?

3.4 Are the confounders accounted for in the design and analysis?

3.5 During the study period, is the intervention administered (or exposure occurred) as intended?

4.1 Is the sampling strategy relevant to address the research question?

4.2 Is the sample representative of the target population?

4.3 Are the measurements appropriate?

4.4 Is the risk of nonresponse bias low? (Say yes if response rate higher than 60%)

4.5 Is the statistical analysis appropriate to answer the research question?

5.1 Is there an adequate rationale for using a mixed methods design to address the research question?

5.2 Are the different components of the study effectively integrated to answer the research question?

5.3 Are the outputs of the integration of qualitative and quantitative components adequately interpreted?

5.4 Are divergences and inconsistencies between quantitative and qualitative results adequately addressed?

5.5 Do the different components of the study adhere to the quality criteria of each tradition of the methods involved?

### S5 – References for general indicators

| General indicators of appropriateness | References |
|---|---|
| **Treatment decision** | |
| Is decision compliant with local, regional, or national guidelines? | 95 (6, 14-17, 19, 23-26, 31, 33, 36, 37, 41, 42, 44, 45, 48-50, 53, 54, 56-59, 62, 66, 67, 69-72, 77-81, 83, 85-91, 93-99, 103-105, 107, 109-117, 120, 122, 124-126, 130, 133, 134, 137, 149-153, 155, 157-161, 164, 166-171, 174) |
| Is decision in-line with expert opinion - e.g. Microbiologist, Infectious Diseases Consultant etc. | 36 (9, 14, 18, 25, 43, 46, 47, 52, 66-68, 77-83, 85-90, 95, 104, 111, 112, 120, 124, 126, 134, 137, 138, 152, 174) |
| Is decision in-line with evidence-base or scientific information | 6 (24, 36, 67, 70, 127, 163) |
| Is decision in-line with decision support tools | 1 (14) |
| **<u>Prior to prescribing - Need for antibiotic</u>** | |
| Are antibiotics indicated? | 21 (9, 25, 41, 42, 47, 50, 52, 65, 79-82, 87, 103, 116, 126, 127, 151, 162, 169, 174) |
| Has the patient got a confirmed or suspected bacterial infection? | 17 (6, 24, 30, 33, 56, 64, 65, 71, 72, 109, 114, 127, 140, 144, 145, 159, 164) |
| Has the patient got clinical signs of an infection? | 13 (6, 16, 19, 33, 50, 64, 65, 67, 111, 125, 144, 151, 155) |
| Have relevant diagnostic criteria been met? | 13 (16, 19, 21-23, 27, 71, 114, 117, 121, 153, 162, 170) |
| Have diagnostic blood cultures been taken prior to antibiotic prescribing? | 12 (6, 30, 91, 92, 97-99, 122, 128, 142, 145, 150) |
| Have diagnostic cultures from suspected site of infection been taken prior to antibiotic prescribing? | 12 (6, 64, 65, 91, 96-99, 115, 122, 145, 150) |
| Has the patient got a risk of infection? | 6 (41, 42, 44, 50, 65, 155) |
| Have relevant diagnostic tests been carried out? | 3 (16, 110, 121) |
| Is the patient classified as palliative, meaning that antibiotics may not be appropriate? | 1 (65) |
| Have relevant severity criteria been met? | 1 (65) |
| **<u>At prescribing</u>** | |
| Is the duration correct? | 70 (9, 14, 17, 19, 25, 26, 31, 33, 44-47, 50, 52-56, 62, 64-66, 68, 70, 77-83, 85-90, 95, 96, 106, 108, 109, 111-113, 120, 122, 124-127, 132-134, 139, 140, 142, 144, 145, 149, 152, 153, 155, 157, 159, 160, 162, 166, 171, 174) |
| Is the dose correct? | 60 (14, 15, 18, 19, 44, 45, 50, 62, 64-72, 77-83, 85-90, 93-95, 97, 106, 108, 109, 111-113, 124-127, 133, 134, 139, 140, 145, 149, 152, 155, 157, 159, 162, 166, 168, 169, 171, 174) |
| Is the choice correct? | 57 (14, 16-19, 24, 26, 31, 37, 42, 44, 53, 64, 65, 68, 69, 71, 72, 77-81, 86, 88-90, 93, 95, 96, 98, 103, 106, 108, 109, 111, 116, 124, 126, 133, 137, 139, 145, 151, 153, 156-160, 162, 164, 166-169, 174) |
| Is the route correct? | 33 (44, 54, 64-68, 77, 79-83, 85-90, 93-95, 106, 111-113, 125-127, 149, 157, 158, 174) |
| Is the frequency correct? | 25 (44, 45, 50, 54, 62, 69, 78, 82, 83, 85-90, 94, 97, 106, 108, 113, 127, 134, 152, 155, 159) |
| Is the timing of administration correct? | 15 (35, 47, 54, 64, 82, 94, 98, 106, 109, 139, 140, 142, 150, 157, 160) |
| Is the medicinal form correct? | 2 (59, 62) |
| ***Have the following factors been considered when choosing the regimen?*** | |
| The indication being treated? | 28 (15-17, 31, 43, 45, 54, 62, 68, 87, 93, 95, 103, 106, 109, 111-113, 116, 125, 137, 139, 145, 152, 155, 158, 163, 167) |
| The activity of the antibiotic against the known or suspected organism? | 26 (9, 18, 52, 64, 65, 67, 77, 79-81, 86, 88-90, 95, 111, 113, 114, 125, 126, 139, 140, 155, 162, 171, 174) |
| The patient's allergy status? | 25 (16, 31, 43, 66, 78-83, 85, 86, 88-90, 94, 95, 113, 125-127, 142, 151, 155, 174) |
| Whether the spectrum of the antibiotic is too broad or too narrow? | 23 (33, 67, 77, 79-83, 85-90, 95, 112, 115, 126, 127, 140, 145, 159, 169, 174) |
| Whether there is any duplication, overlap, or redundancy in the regime? | 19 (9, 18, 43, 45, 46, 52, 64, 65, 72, 79-81, 95, 120, 126, 127, 151, 169, 174) |
| The patient's renal function? | 15 (6, 45, 64-66, 91, 96, 97, 122, 145, 149, 150, 152, 155, 162) |
| The patient's previous antibiotic use? | 6 (43, 44, 64-66, 94) |

| | |
|---|---|
| Whether the IV or oral route is appropriate based on specified criteria? | 6 (46, 59, 65, 98, 128, 155) |
| The patient’s severity criteria? | 5 (49, 64, 65, 142, 152) |
| Whether combination or co-prescribing is needed? | 5 (64-66, 68, 166) |
| The patient’s contraindications? | 5 (45, 67, 78, 94, 127) |
| Whether an inappropriate combination has been prescribed? | 4 (26, 64, 65, 162) |
| The patient’s previous culture results, including colonisation? | 4 (43, 94, 95, 151) |
| The patient’s gender? | 4 (64, 65, 96) (26) |
| The patient’s co-morbidities? | 3 (44, 113, 163) |
| The patient’s weight? | 3 (15, 64, 155) |
| Interactions with the patient’s other medicines or feeds? | 3 (18, 45, 128) |
| Whether the patient is pregnant | 2 (19, 26, 104) |
| The patient’s risk of adverse events? | 2 (106, 130) |
| Whether it will penetrate to the target site? | 2 (127, 134) |
| The patient’s risk factors for resistant bacteria? | 2 (49, 151) |
| The mode of action of the antibiotic? | 1 (67) |
| Whether the patient has a catheter? | 1 (96) |
| The patient’s immune status including immunosuppressive medicines? | 1 (65) |
| The patient’s travel history? | 1 (155) |
| The practicality of the antibiotic? | 1 (45) |
| **After initial prescribing** | |
| Have cultures been reviewed and treatment adapted accordingly? | 35 (6, 24, 31, 33, 43, 46, 70-72, 78, 82, 83, 85-91, 96-99, 105, 111, 122, 130, 134, 149-152, 155, 168, 171) |
| Has there been appropriate documentation in the clinical record? | 29 (16, 31, 56-58, 71, 83, 91, 93, 94, 97, 99, 106, 108, 120, 122, 136, 145, 150-152, 155, 157-159, 161, 171, 174, 175) |
| Has intravenous treatment been reviewed and switched to oral treatment (IVOS) appropriately? | 16 (18, 67, 91, 95-97, 99, 120, 122, 128, 144, 145, 149, 153, 155, 162) |
| Has appropriate Therapeutic Drug Monitoring taken place? | 9 (6, 64, 65, 91, 92, 120, 122, 142, 155) |
| Has the treatment been de-escalated? | 6 (6, 115, 120, 144, 153, 162) |
| Has the treatment been stopped if there is no evidence of infection? | 5 (9, 91, 122, 127) (52) |
| Has the treatment been reviewed or reassessed? | 5 (31, 71, 97, 149, 164) |
| Has the treatment been effective? | 2 (45, 106) |
| Has the treatment been monitored? | 1 (18, 67) |
| Is the patient taking it? | 1 (91) |
| Has the patient been educated on how to take it? | 1 (94) |

### S6 – References for proxy indicators

| **Proxy Indicators of appropriateness** | **References** |
|---|---|
| **Consumption Indicators** | |
| Fluoroquinolones | 6 (100-102, 118, 135, 173) |
| Penicillins plus beta-lactamase inhibitor | 5 (101, 102, 118, 135, 173) |
| 3rd and 4th generation cephalosporin | 5 (101, 118, 135, 143, 173) |
| Broad/Narrow ratio | 5 (63, 101, 118, 119, 173) |
| 1st line/2nd line ratio | 4 (5, 73, 75, 76) |
| All penicillins | 3 (101, 118, 173) |
| Beta-lactamase sensitive penicillins | 3 (101, 118, 173) |
| Macrolides, lincosamides, and streptogramins | 3 (101, 118, 173) |
| Cephalosporins | 3 (101, 118, 173) |
| Amoxicillin/amoxicillin-clavulanate ratio | 2 (73, 74) |
| Macrolides | 2 (100, 102) |
| Amoxicillin-clavulanate + Macrolides + Quinolones | 2 (100, 102) |
| Penicillin | 1 (146) |
| Amoxicillin | 1 (119) |
| Amoxicillin-clavulanate | 1 (100) |
| Azithromycin | 1 (119) |
| Clindamycin | 1 (146) |
| Moxifloxacin | 1 (109) |
| Amoxicillin and cefalexin | 1 (119) |
| Pristinamycin + macrolides | 1 (73) |
| Carbapenems and piperacillin-tazobactam | 1 (51) |
| Drugs that make up 90% of antibiotic volume (DU90) | 1 (118) |
| Antibiotic diversity | 1 (34) |
| % 'Access' category | 1 (141) |
| ***Route*** | |
| Intravenous antibiotics | 2 (148) |
| ***Duration*** | |
| Duration greater than specified days | 5 (5, 63, 73-75) |
| Days of therapy | 2 (40, 143) |
| Mean duration | 1 (119) |
| ***Seasonality*** | |
| Seasonality of total antibiotic use | 8 (5, 63, 73, 75, 76, 101, 118, 173) |
| Seasonality of quinolone use | 6 (5, 73, 75, 101, 118, 173) |
| Seasonality of amoxicillin-clavulanate use | 1 (63) |
| **Patient Specific Indicators** | |
| ***Need for antibiotic*** | |
| Rate of patients prescribed an antibiotic based on the specified indication | 22 (20, 28, 29, 32, 38, 39, 51, 60, 61, 84, 100, 102, 109, 119, 123, 129, 131, 146, 147, 154, 156, 165) |
| Patients having appropriate diagnostic tests before prescription of antibiotic | 3 (38, 109, 172) |
| Patients meeting required diagnostic criteria before prescription of antibiotic | 2 (84, 172) |
| ***Choice*** | |
| Patients prescribed the first line choice for the specified indication | 14 (5, 20, 61, 63, 73, 75, 100, 102, 109, 119, 154, 156, 165, 172) |
| Patients prescribed a fluoroquinolone for a specified indication | 5 (61, 154, 156, 165, 172) |
| Patients co-prescribed a fluoroquinolone and steroid | 5 (5, 73, 75, 76, 135) |
| Patients prescribed antibiotic and non-steroidal anti-inflammatory drug (NSAID) on the same day | 5 (5, 63, 73, 75, 76) |
| Patients prescribed a fluoroquinolone within 6 months of another prescription for a fluoroquinolone | 4 (5, 63, 73, 75) |
| Patients prescribed an antibiotic that is not indicated | 3 (5, 73, 75) |
| Patients prescribed an inappropriate antibiotic choice for the indication | 2 (74, 172) |
| Patients prescribed a rarely used antibiotic choice for the indication | 1 (74) |
| ***Patient factors*** | |
| Patients without known penicillin allergy prescribed a pen-allergic treatment option | 1 (172) |
| Pregnant patients prescribed doxycycline | 1 (109) |
| Male patients receiving 1st generation fluoroquinolones, nitrofurantoin or fosfomycin | 1 (109) |
